# Protein-based genomic analysis for the identification of risk loci associated with acute respiratory distress syndrome

**DOI:** 10.64898/2026.01.14.26344107

**Authors:** Eva Suarez-Pajes, Luis A. Rubio-Rodríguez, Eva Tosco-Herrera, Melody Ramirez-Falcon, Silvia Gonzalez-Barbuzano, David Jáspez, Adrián Muñoz-Barrera, Tamara Hernandez-Beeftink, Almudena Corrales, Elena Espinosa, David Domínguez, Rafaela González-Montelongo, Jose M. Lorenzo-Salazar, M. Isabel García-Laorden, Jesús Villar, GEN-SEP study, Beatriz Guillen-Guio, Carlos Flores

## Abstract

**Background:** Acute respiratory distress syndrome (ARDS) is a life-threatening lung condition that requires admission to an intensive care unit (ICU). Sepsis is one of the leading causes of ARDS and understanding protein regulation during sepsis could reveal key mechanisms that predispose patients to ARDS. We performed genome-wide association studies (GWAS) on ARDS biomarkers levels to identify protein quantitative trait loci (pQTLs) and genes which could be associated with ARDS risk.

**Methods:** GWAS were performed in 209 patients with sepsis from the GEN-SEP cohort to determine the association of imputed genotypes with 10 serum biomarker levels relevant to ARDS. Measurements were obtained by ELISA within the first 24 hours (T1), 48-72 hours (T2), and 7 days (T7) after the diagnosis of sepsis. We conducted a multi-trait analysis to aggregate the GWAS results for each biomarker at three time points. We prioritized genes in the significant loci (*p*<5×10^-8^) and evaluated the association between rare variants and ARDS in whole-exome sequencing data from 272 patients with sepsis-associated ARDS and 550 sepsis controls from GEN-SEP. We analyzed the aggregated association of pQTLs with ICU mortality, multiple organ failure, and ARDS risk using polygenic scores (PGS) in independent patients (n=621) from GEN-SEP.

**Results:** We identified 27 significant independent loci and prioritized 56 genes. Seven of these were previously associated with respiratory infections and diseases (*LINGO2*, *MC4R*, *MCTP1*, *NUAK1*, *PIEZO2*, *PTPRD*, and *TMEM65*). Defects in another prioritized gene, *FOXN1*, cause an inborn error of immunity. Rare variants in *PTPRD*, which was previously involved in COVID-19 severity and pulmonary hypertension, were significantly associated with ARDS (*p*=3.11×10^-4^). PGS of PAI-1 levels was significantly associated with ICU mortality.

**Conclusions:** We prioritized genes of interest governing ARDS biomarker levels and identified *PTPRD* as a novel gene associated with ARDS risk. In addition, we demonstrate the value of biomarker PGS for predicting sepsis mortality.

Graphical Abstract

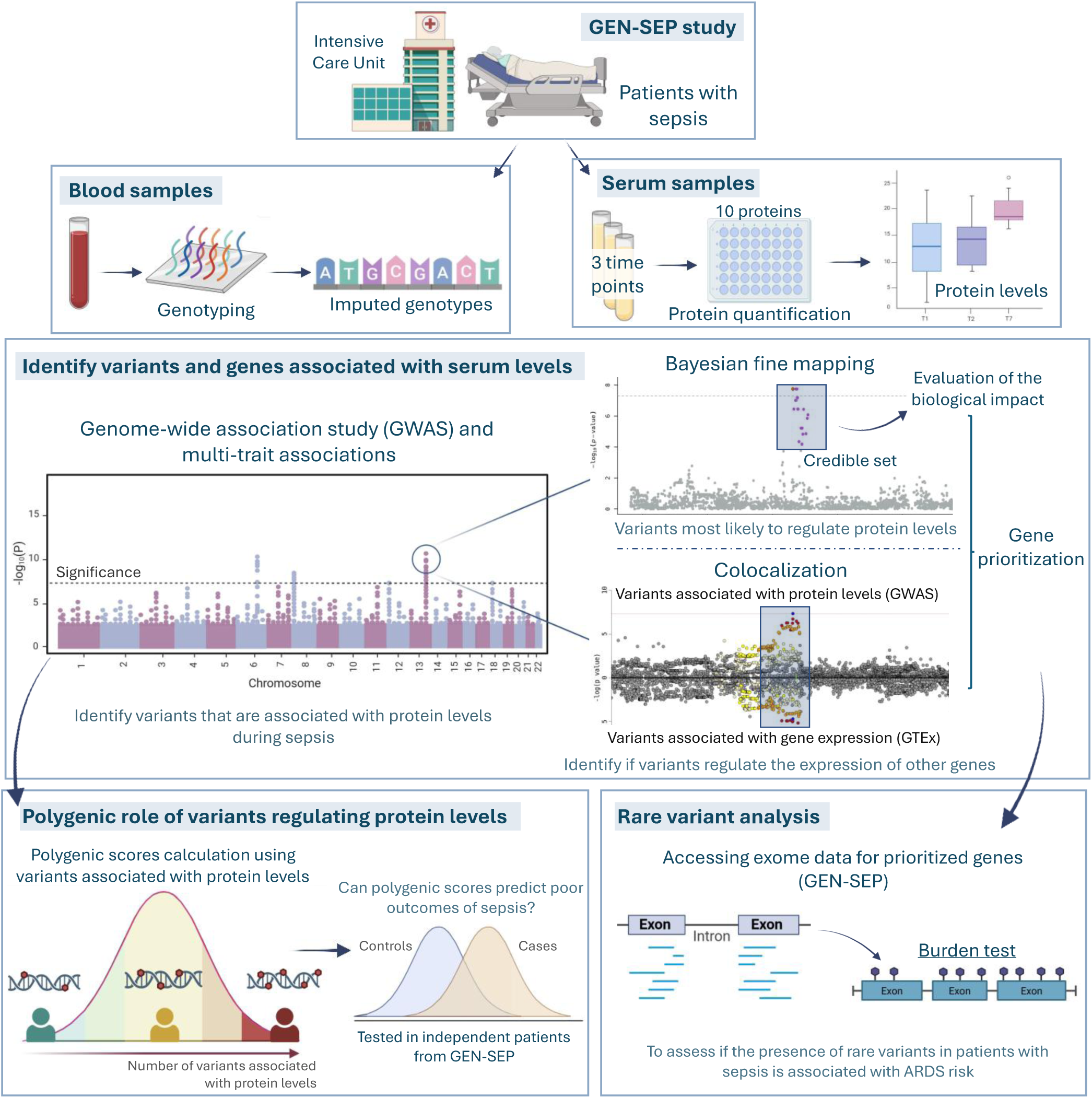

## Background

Acute respiratory distress syndrome (ARDS) is a severe life-threatening lung condition that requires admission to the intensive care unit (ICU) and the use of mechanical ventilation. Sepsis is one of its leading causes. However, ARDS can also result from trauma, aspiration, or severe viral or bacterial infections, among others (1).

ARDS is characterized clinically by the development of a non-cardiogenic pulmonary edema, bilateral pulmonary infiltrates, and severe hypoxemia. Diagnosis is based on chest imaging findings and the severity of hypoxemia (2,3). The mortality rate can exceed 40% in the most severe forms, and the current therapeutic options consist of life-support interventions to improve oxygenation and minimize the inflammatory response (4)

The alveolar-capillary barrier is impaired during ARDS, and the immune and inflammatory response is also dysregulated. Several proteins involved in key pathophysiological processes have been proposed as ARDS biomarkers in the literature. These include proteins linked to endothelial injury (Ang-2, ICAM-1, and vWF), coagulation or fibrinolysis (PAI-1 and thrombomodulin), epithelial injury (RAGE and SP-D), and inflammation (interleukins, TNF-α, and D-dimer) (5,6). Many studies have aimed to identify and assess the utility of biomarkers for the diagnosis and prognosis of ARDS (7–9). However, there is no consensus on their use in clinical practice, highlighting the need for further validation.

Proteogenomic studies provide insights into the functional effects of genetic variation governing protein regulation, which are essential for understanding disease susceptibility and progression. Several studies have used proteogenomic approaches in large population-scale datasets. However, gene regulation and quantitative trait loci (QTLs) can be influenced by the biological context (10,11). Understanding the genetic regulation of protein levels during sepsis may reveal key mechanisms that predispose patients to ARDS. One example is vascular endothelial growth factor receptor 1 (VEGFR1), a protein encoded by the *FLT1* gene that plays a key role in angiogenesis and vascular permeability. Variants of *FLT1* have been associated with susceptibility to sepsis-associated ARDS (12) and protein QTLs (pQTLs) that regulate VEGFR1 during sepsis were also associated with ICU mortality and ARDS risk (13). In addition, pQTL studies supported *TCF20* as a novel ARDS risk gene (13).

Here, we studied the genetic loci associated with levels of 10 serum proteins with strong literature support as biomarkers of key pathophysiological processes involved in ARDS. We first identified pQTLs associated with protein levels at three time points after sepsis diagnosis. Then, we evaluated the association of the polygenic component regulating biomarkers with ARDS risk, multiple organ failure (MOF), and ICU mortality. These approaches allowed us to prioritize genes of potential interest governing biomarker levels and to test the association of rare variants in these genes with susceptibility to sepsis-associated ARDS.

## Material and Methods

### Study design and quantification of protein biomarkers

We obtained samples from 221 ICU patients diagnosed with sepsis from the GEN-SEP study (14,15) (**Figure 1**). Samples were classified into different intervals according to the collection time. T1: within the first 24 hours after diagnosis, T2: between 48 and 72 hours after diagnosis, and T7: 7 days after diagnosis.

**Figure 1.**
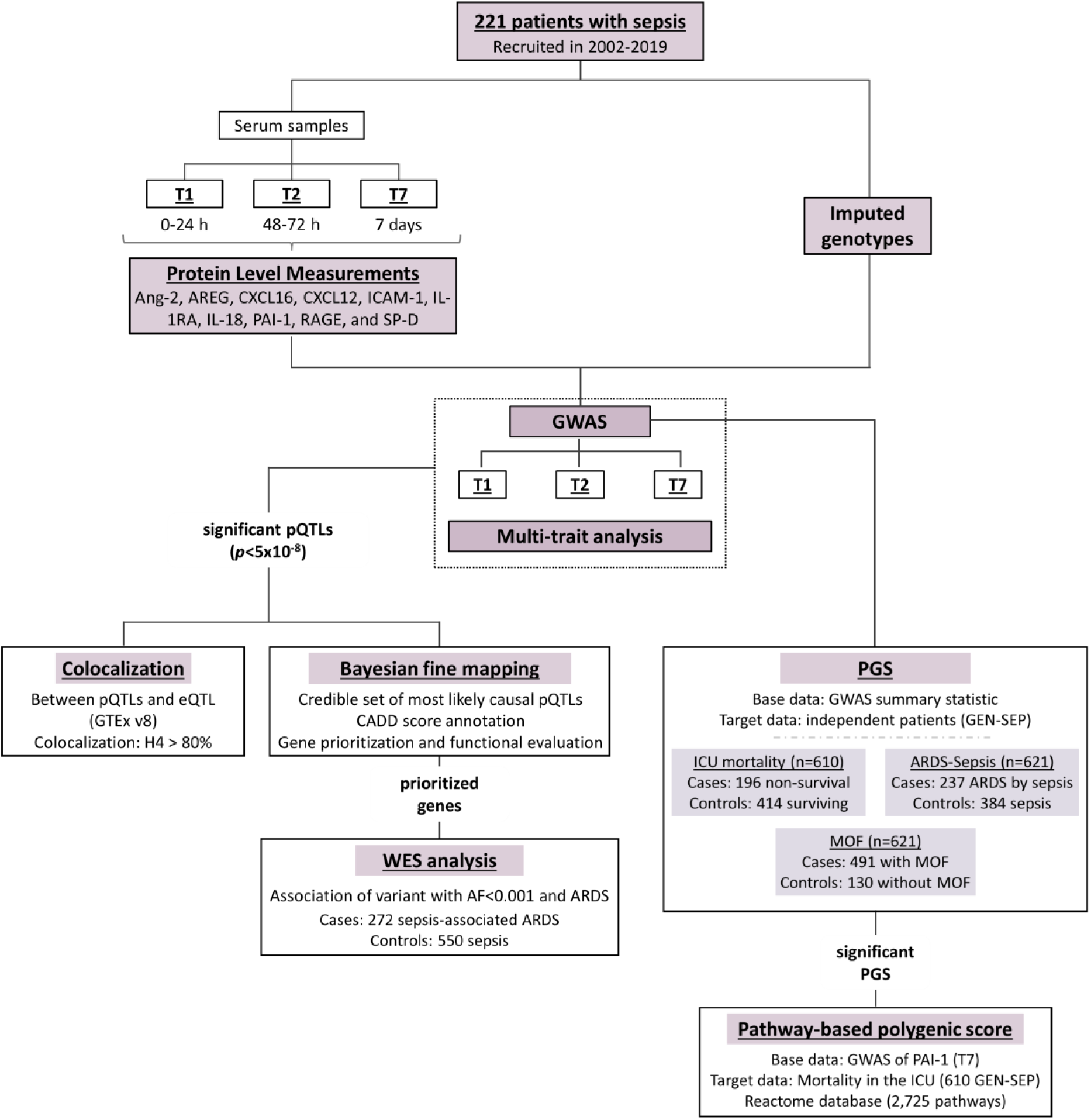
Desing study flowchart. CADD: combined annotation dependent depletion score; WES: whole-exome sequence data; AF: allele frequency; eQTL: expression quantitative trait locus; GWAS: genome-wide association study; ICU: intensive care unit; MOF: multiple organ failure, PGS: polygenic score; pQTLs: protein quantitative trait locus.

For the three collection points, we quantified serum protein levels of 10 literature-supported ARDS biomarkers: Ang-2 (angiopoietin-2), AREG (amphiregulin), CXCL16 (C-X-C motif chemokine ligand 16), CXCL12 (C-X-C motif chemokine ligand 12), ICAM-1 (intercellular adhesion molecule 1), IL-1RA (interleukin-1 receptor antagonist), IL-18 (interleukin-18), PAI-1 (plasminogen activator inhibitor-1), RAGE (receptor for advanced glycation end products), and SP-D (surfactant protein D) (7). Protein levels were measured separately using the DuoSet ELISA kits and DuoSet Ancillary Reagent Kit2 (RCD Systems, Abingdon, UK) following the manufacturer’s protocol. Protein concentrations were normalized by Box-Cox transformations, and normality was assessed using the Lilliefors test. Further details of the study design and serum measurements are available in **supplementary methods**.

### GWAS and multi-trait associations

To identify pQTLs for the 10 biomarkers, we performed GWAS to test the association of imputed genotypes with the protein levels in each of the three collection time points using a linear Wald test. Further information about genotyping, quality controls, imputation, and association procedures are described in detail in the **supplementary methods**.

To leverage the pleiotropic effects of genetic variants and improve statistical power, we performed a multi-trait analysis based on the GWAS results. For that, we aggregated the three-time points GWAS summary statistics for each biomarker using MTAG v.1.0.8 (16).

Statistical significance for GWAS and multi-trait analysis was declared at the genome-wide threshold (*p*<5×10^-8^). Sentinel variants were deemed as those surpassing the significance threshold and showing weak linkage disequilibrium (LD; r^2^ < 0.1) with others in each locus after PLINK-based clumping (17).

### Variant and gene prioritization

To prioritize putative causal variants and genes of interest, we explored a 1 megabase (Mb) window around the sentinel variants detected on GWAS and multi-trait analyses. We performed a Bayesian fine mapping for the detection of credible set of variants with 95% confidence most likely to drive the association. In each credible set, we prioritized the variant with the highest Combined Annotation Dependent Depletion (CADD) score as that with the most likely highest biological impact (18). We then assigned them the most likely affected gene based on functional evidence, as integrated in the Variant-to-Gene (V2G) score (https://platform.opentargets.org/).

We established a gene set by including genes that were positionally closest to the sentinel variant and those prioritized by the V2G ranking. We then searched the gene set in the Open Targets Platform to identify previous GWAS findings (https://platform.opentargets.org/) and in OMIM to evaluate their relationships with Mendelian diseases (https://www.omim.org/).

Finally, we performed a colocalization analysis between the significant loci and eQTL data from GTEx v8 (https://gtexportal.org/) to detect whether the identified pQTLs could also associate with gene expression. For this analysis, we only included genes whose sentinel variant was an eQTL, and colocalization was declared when the posterior probability (PP) of H4 exceeded 80%. A more detailed description of post-association analyses is available in the **supplementary methods**.

### Rare variant analyses in prioritized genes

We assessed the association of rare variants in prioritized genes with risk to sepsis-associated ARDS. Whole exome sequencing (WES) data were available from 822 patients in the GEN-SEP study: 550 patients with sepsis used as controls and 272 patients with sepsis-associated ARDS considered as cases (19). We performed a SKAT-O test to evaluate two categories of variants (20): i) those with allele frequency (AF) <0.001, and ii) those with AF <0.001 and evidencing high biological impact (see **supplementary methods** for further details). The significance threshold was declared after Bonferroni correction considering the number of genes and the two variant categories tested (*p*=6.25×10^-4^). We also tested the association of synonymous variants at the same AF threshold as an internal control to ensure that residual confounding due to patient heterogeneity did not affect the results.

The genetic damage index (GDI) v2 (https://hgidsoft.rockefeller.edu/GDI/GDIv2.html) was used to annotate the tolerance of genes to mutations. Detailed information about the procedures is provided in **supplementary methods**.

### Polygenic score derivation and testing in independent GEN-SEP patients

We evaluated the association between the polygenic component of the 10 biomarkers and sepsis outcomes using polygenic scores (PGS). GWAS results were used as base data, and we applied a clumping and thresholding approach to screen multiple models. PGS were then tested for association in an independent cohort of 621 GEN-SEP patients to identify associations with susceptibility to: i) sepsis-associated ARDS (237 cases and 384 controls); ii) MOF (491 cases and 130 controls); and iii) ICU mortality (196 cases and 414 controls). The significance threshold for these analyses was established at *p*=5.56×10^-4^ after Bonferroni correction (0.05/90). For the significantly associated PGS, we performed a pathway-based PGS analysis to evaluate the biological processes linking the sepsis outcome with the polygenic component of the biomarker. Further details are provided in the **supplementary methods**.

## Results

After quality control, we included 209 patients with sepsis that had at least one protein measure available at any time point, showing temporal variation in biomarker levels over time (**Table S1** and **Figure S1**). The sample size for the association studies is available in **Supplementary Table S1**. Demographic and clinical characteristics are provided in **Supplementary Table S2**.

### GWAS and multi-trait associations

The association analyses identified 27 significant sentinel signals associated with protein levels. Of these, multi-trait association detected 25, while two were identified by GWAS (**Table 1**, **Figure S2-S5**). The sentinel variants at GWAS significant loci were rs76099260 (*p*=2.04×10^-8^, beta=-3.38, standard error [SE]=0.58), which is intergenic to *CCZ1B* and *MIR3683*, and rs113037345 (*p*=4.92×10^-8^, beta=-0.73, SE=0.13), which is intronic to *ITGA1*. They were significantly associated with reduced levels of Ang-2 and PAI-1 at T1, respectively. Genomic inflation factors (λ) values ranged from 0.96 to 1.02, indicating no evidence of inflation in the association results (**Figures S6-S7**).

**Table 1.** Significant (p<5×10^-8^) sentinel variants obtained from GWAS and multi-trait associations.

| Biomarker (Time) | rsID | Position (hg19) | nEA/EA | Freq | Beta(SE) | p-value | Gene(s) |
| --- | --- | --- | --- | --- | --- | --- | --- |
| Ang-2 (T1) | rs76099260* | 7:7054699 | A/T | 0.01 | -3.38(0.58) | $2.04 \times 10^{-8}$ | CCZ1B\<br>MIR3683 |
| | rs77880844 | 14:81683460 | G/A | 0.04 | -1.30(0.24) | $3.67 \times 10^{-8}$ | GTF2A1 |
| AREG (T2) | rs7970411 | 12:125706376 | T/G | 0.04 | 1.68(0.30) | $2.03 \times 10^{-8}$ | TMEM132B |
| AREG (T7) | rs731153 | 2:240458353 | G/A | 0.29 | 0.82(0.15) | $4.59 \times 10^{-8}$ | HDAC4-AS1\<br>LOC150935 |
| | rs12554442 | 9:8427879 | T/G | 0.15 | 1.15(0.19) | $1.19 \times 10^{-9}$ | PTPRD |
| | rs78986031 | 9:28255496 | G/A | 0.02 | -2.56(0.46) | $2.04 \times 10^{-8}$ | LINGO2 |
| | rs10778455 | 12:106439029 | G/A | 0.45 | 0.77(0.14) | $1.82 \times 10^{-8}$ | CASC18\<br>NUAK1 |
| | rs9510995 | 13:24590355 | G/A | 0.29 | -0.83(0.15) | $3.22 \times 10^{-8}$ | SPATA13 |
| | rs118062838 | 18:58133225 | G/A | 0.03 | -2.17(0.40) | $4.16 \times 10^{-8}$ | MC4R\<br>CDH20 |
| CXCL16 (T1) | rs7517208 | 1:114915192 | A/G | 0.08 | 0.85(0.15) | $2.77 \times 10^{-8}$ | SYT6\<br>TRIM33 |
| | rs73880250 | 3:172441323 | G/A | 0.04 | 1.17(0.21) | $3.13 \times 10^{-8}$ | NCEH1\<br>ECT2 |
| ICAM-1 (T2) | rs148202393 | 1:46618112 | A/G | 0.01 | -2.35(0.43) | $3.70 \times 10^{-8}$ | P3R3URF-PIK3R3\<br>PIK3R3 |
| | rs726017 | 2:234725819 | A/G | 0.18 | 0.69(0.12) | $1.83 \times 10^{-8}$ | MROH2A |
| | rs2220449 | 4:117336713 | G/C | 0.01 | -2.45(0.43) | $9.11 \times 10^{-9}$ | MIR1973\<br>TRAM1L1 |
| IL-1RA (T7) | rs35003 | 5:16617099 | A/G | 0.16 | 2.14(0.37) | $1.06 \times 10^{-8}$ | LOC101929524 |
| | rs11156083 | 6:156474813 | G/A | 0.31 | -1.67(0.29) | $1.30 \times 10^{-8}$ | MIR1202\<br>SNORD28B |
| | rs11658852 | 17:15300324 | G/A | 0.48 | 1.66(0.27) | $9.56 \times 10^{-10}$ | TEKT3\<br>TVP23C-CDRT4 |
| IL-18 (T1) | rs140560482 | 5:94675417 | G/T | 0.02 | 1.87(0.32) | $3.96 \times 10^{-9}$ | MCTP1\<br>FAM81B |
| IL-18 (T2) | rs7235298 | 18:11263135 | C/A | 0.03 | -1.58(0.28) | $1.80 \times 10^{-8}$ | PIEZO2\<br>LINC01928 |
| IL-18 (T7) | rs62519200 | 8:125239839 | C/T | 0.11 | -0.99(0.17) | $7.46 \times 10^{-9}$ | LOC101927588 |
| PAI-1 (T1) | rs113037345* | 5:52217786 | G/A | 0.01 | -0.73(0.13) | $4.92 \times 10^{-8}$ | ITGA1 |
| RAGE (T2) | rs28413183 | 21:45471684 | C/T | 0.12 | 0.85(0.15) | $6.23 \times 10^{-9}$ | TRAPPC10 |
| RAGE (T7) | rs2476174 | 13:80174234 | A/G | 0.01 | 2.85(0.51) | $2.61 \times 10^{-8}$ | LINC01068\<br>LINC01038 |
| | rs142329471 | 13:112875863 | G/A | 0.02 | -2.20(0.40) | $3.49 \times 10^{-8}$ | LINC01070\<br>LOC101928730 |
| | rs117886541 | 17:26831908 | T/C | 0.02 | -2.43(0.44) | $4.59 \times 10^{-8}$ | SLC13A2\<br>FOXN1 |
| SP-D (T1) | rs62500471 | 8:53777234 | A/G | 0.25 | -0.41(0.07) | $2.09 \times 10^{-9}$ | RB1CC1\<br>NPBWR1 |
| SP-D (T7) | rs62500471 | 8:53777234 | A/G | 0.24 | -0.68(0.12) | $7.70 \times 10^{-9}$ | RB1CC1\<br>NPBWR1 |

\*Results from GWAS. Beta: effect; EA: effect allele, Freq: effect allele frequency, nEA: non effect allele, SE: standard error. T1: levels obtained within 24 hours after sepsis diagnosis; T2: levels obtained within 48-72 hours after sepsis diagnosis; T7: levels obtained 7 days after sepsis diagnosis.

### Variant and gene prioritization

Bayesian fine-mapping around the 27 sentinel variants provided the most likely pQTLs that are associated with biomarker levels during sepsis (**Table S3**). We obtained credible sets for all regions except for the locus around the variant rs113037345 in PAI-1 for the GWAS at T1 (**Figure S8**). To assess the biological relevance of variants included in the credible sets, we annotated the CADD score and found 33 variants in 16 loci showing putative biological impact (CADD>10.00) linked to associations with Ang-2, AREG, CXCL16, ICAM-1, IL-1RA, IL-18, RAGE, and SP-D levels (**Table S4**). Among these, two variants were exonic to *MROH2A*, one corresponding to a missense variant (rs1500480, *p*=1.83×10^-8^, CADD=14.41), while the rest of variants identified were intronic (*TMEM132B*, *PTPRD*, *LINGO2*, *TRIM33*, *MAST2*, and *FAM81B)* or intergenic (**Table S4**).

Based on the association findings, we prioritized 56 genes, including those closest to each sentinel variant and those with the strongest biological relevance (**Table S5**). Of these, 40 were protein-coding genes, while 14 genes were classified as non-coding RNAs, including small nucleolar, non-coding, and micro-RNAs (**Table S6**). Based on publicly available GWAS findings available on the Open Targets Platform (https://platform.opentargets.org/), 12 out of the 56 genes (21.4%) were associated with infectious diseases and 14 (25%) with respiratory traits, seven of which were common to both (*LINGO2*, *MC4R*, *MCTP1*, *NUAK1*, *PIEZO2*, *PTPRD*, and *TMEM65).* In addition, most of these genes were associated with endocrine and metabolic traits, as well as with hematopoietic and circulatory system traits (**Figure 2**). Worth highlighting, *PTPRD* was previously associated with severe acute respiratory syndrome in a study of COVID-19 patients (21). Links with Mendelian diseases revealed 11 additional genes based on evidence reported in OMIM, including *FOXN1* which has been implicated in an inborn error of immunity (**Table S7**).

**Figure 2.**
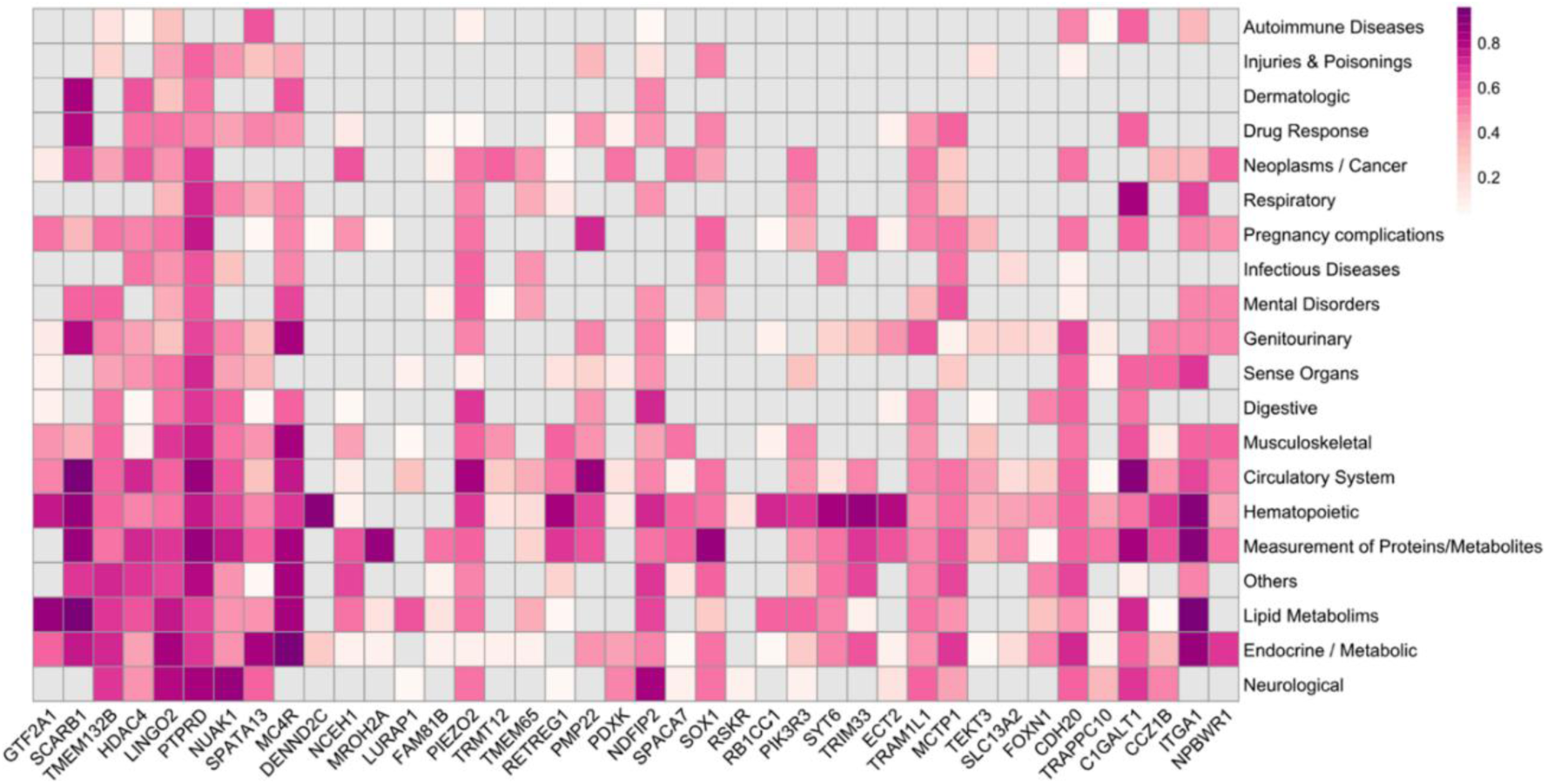
Evidence from previous GWAS associations of the prioritized genes. Data obtained from the Open Targets Platform (https://platform.opentargets.org/).

Colocalization analyses supported molecular connections between gene expression and biomarker pQTLs in four of the prioritized genes. Variants regulating AREG levels colocalized with *CASC18* eQTLs in subcutaneous adipose tissue (T7; H4=91.62%). Additionally, pQTLs for ICAM-1 levels colocalized with *MROH2A* in skin (T2; H4=93.10%) and the esophagus (T2; H4=88.30%). Colocalizations between *RB1CC1* and *NPBWR1* with SP-D levels were found in the gastroesophageal junction of the esophagus (T1; H4=85.41%), and in the liver (T7; H4=87.38%), respectively (**Table S8**). No further colocalizations with the prioritized genes were observed (**Figure SG**).

### Rare variant analyses in prioritized genes

Analysis of rare genetic variation was performed on the 40 prioritized protein coding genes for which WES was available (**Table SG**). Among these, rare variants at *PTPRD* were significantly associated with ARDS risk after Bonferroni correction (*p*=3.11×10^-4^). The association was attenuated when analyses were restricted to rare variants with high biological impact (**Table 2**, **Table S10**). In addition, a rare variant with a high biological impact at *NCEH1* (rs754149316, AF=0.0006, CADD=11.59) was nominally associated with ARDS risk (*p*=0.005) (**Table 2**, **Table S11**). According to the GDI scores, both *PTPRD* (GDI=0.08) and *NCEH1* (GDI=1.15) have reduced tolerance to mutations.

**Table 2.** Significant gene associations based on rare-variant analysis (*p*<0.05).

| Gene | Position | Category | Variants | p-value |
| --- | --- | --- | --- | --- |
| *PTPRD* | chr9:8314246-10612723 | AF<0.001 | 100 | $3.11 \times 10^{-4}$ |
|  |  | AF<0.001& biological impact | 6 | 0.025 |
| *NCEH1* | chr3:172348039-172429008 | AF<0.001 | 36 | 0.253 |
|  |  | AF<0.001& biological impact | 1 | 0.005 |

AF: allele frequency, Position: chromosome, start and end position according to Ensembl annotation (GRCh37/hg19). Significance threshold $p=3.11 \times 10^{-4}$ .

### Polygenic score and evaluation of poor sepsis outcomes

The PGS of PAI-1 levels at T7, which included 3,896 variants, was significantly associated with ICU mortality (Odds Ratio [OR]=0.71, 95% confidence interval [CI]=0.59-0.86, *p*=3.26×10^-4^), surpassing the significance threshold of *p*=5.56×10^-4^ (**Table S12**). No further significant PGS models were observed in association with ARDS risk and MOF (**Table S13**, **S14**). The biological processes enriched on this PGS model of PAI-1 were RNA polymerase I promoter opening (*p*=3.94×10^-5^), and reversal of alkylation damage by DNA dioxygenases (*p*=1.21×10^-3^), although none of them could be considered significant when adjusting for multiple comparisons (*p*=1.83×10^-5^) (**Table S15**).

## Discussion

Genetic regulation of ARDS biomarkers could be key to identify new genetic loci associated with ARDS risk, as well as novel therapeutic targets. Here, we integrated GWAS results with a multi-trait association approach to prioritize genes based on biomarker pQTLs and to test their association with severe outcomes in patients with sepsis. In the process, we also relied on rare variant testing on the prioritized genes to assist in identifying new ARDS risk genes.

Multi-trait analyses were previously applied in different disease traits, including COVID-19 (22–25). However, to our knowledge, this is the first study using multi-trait analysis to assist in the identification of disease genes in sepsis outcomes, allowing to increase the power to prioritize genetic loci beyond the traditional GWAS. The prioritized genes were associated with diverse circulatory, hematopoietic cell, and platelet traits, which could have links with sepsis in the profound dysregulation of immune cell production and apoptosis (26), and the cardiac dysfunction and vasodilation (27).

We identified *PTPRD* as the main biologically relevant gene for AREG, that encodes the protein tyrosine phosphatase D-type receptor. Rare variants at *PTPRD* were also significantly associated with sepsis-associated ARDS in our study. *PTPRD* has been associated with neurogenesis and cancer and has been proposed as a tumor suppressor gene across multiple cancer types (28–31). Additional evidence supports its role in hypertension and blood pressure in response to atenolol, a beta-blocker, as well as in gestational and type 2 diabetes (32–35). *PTPRD* has been proposed as a shared risk gene in both type 2 diabetes and COVID-19 (36). A GWAS of COVID-19 also identified gene variants at *PTPRD* significantly associated with severity to COVID-19 (21). There is compelling evidence supporting its role in other respiratory conditions such as pulmonary hypertension (37), a common complication of ARDS, that has been associated with reduced systemic oxygenation and may contribute to the development of MOF (38,39).

In addition, PTPRD participates in multiple cellular processes, including growth, migration, differentiation, and cell adhesion (30,40). PTPRD interacts in the immune response, particularly with STAT3 and the Wnt/β-catenin axis (31,41). STAT3 is a key transcription factor in immunity, regulating the transcription of cytokines and growth factors (42), and PTPRD downregulates STAT3. Mutations in *PTPRD* have been associated with changes in STAT3 phosphorylation and increased activation in squamous cell carcinoma of the head and neck (30). STAT3 has been linked to ferroptosis in sepsis-associated ARDS, and its inhibition suggests a protective role in mice with acute lung injury (43,44). Thus, further studies are necessary to better understand the role of PTPRD in ARDS risk, in particular in its regulation during the acute inflammatory stage.

We prioritized *FOXN1* (Forkhead Box N1) gene from RAGE multi-trait association, which has been linked to the development and differentiation of T cells in the thymus (45). Defects in this gene, which cause a severe combined immunodeficiency, involve a reduction in T cells, severe and recurrent respiratory infections, and other extra-hematopoietic features (46,47). Furthermore, it has been observed that *FOXN1* expression is downregulated in mouse models of sepsis induced by lipopolysaccharide, affecting thymus function (45). However, further studies are required to investigate the role of *FOXN1* in sepsis outcomes.

Another prioritized gene was *PIEZO2* (Piezo Type Mechanosensitive Ion Channel Component 2), which encodes a mechanotransduction ion channel involved in the conversion of mechanical stimuli into biological signals (48). PIEZO2 has been proposed as a biomarker for ARDS severity (49). Higher protein levels in bronchoalveolar lavage samples were observed in ARDS patients compared to ICU controls and showed positive correlation with severity (49). A WES study identified variants in *PIEZO2* associated with ARDS mortality (50) In addition, this gene has been involved in pulmonary hypertension and angiogenesis (51).

The evaluation of PGS for predicting severe outcomes of sepsis showed a significant association between the polygenic component of PAI-1 levels and ICU mortality among patients with sepsis. PAI-1 inhibits the fibrinolytic process that promotes thrombus formation. Higher levels of PAI-1 have been associated with sepsis severity and mortality (7,52). Our findings are consistent with the previous evidence since patients with lower PGS of PAI-1, i.e., which were associated with decreased PAI-1 levels, were associated with reduced ICU mortality. Given the complexity of the acute immune response during sepsis, additional studies are necessary to understand the effect of variants associated with PAI-1 levels and their value on the severity prediction.

We acknowledge several limitations of our study. First, our analysis was restricted to a selected panel of proteins previously reported as ARDS biomarkers. Given the heterogeneous and complex nature of ARDS, this selection may have overlooked other potentially relevant proteins that contribute to severe outcomes in sepsis patients. Expanding the protein panel in future analyses could increase the number of prioritized loci and enhance our understanding of ARDS. Second, it would be important to increase the sample size and include independent and ethnically diverse replication cohorts to validate our findings and ensure their generalizability. Finally, functional studies are needed to confirm the causal relationships between the identified genetic loci and sepsis outcomes.

### Conclusions

This study supports the utility of biomarker pQTLs as a tool for identifying novel loci of interest for severity outcomes among sepsis patients. In addition, we identified *PTPRD* as a novel gene associated with sepsis-associated ARDS. Further functional studies and replication in independent cohorts are needed to validate our findings.

## Declarations

### Ethics approval and consent to participate

Written informed consent was obtained from all patients. The study was performed according to The Code of Ethics of the World Medical Association (Declaration of Helsinki) and approved by the Ethics Committee for Drug Research from the Hospital Universitario de Canarias-Provincia de Santa Cruz de Tenerife (Code: CHUNSC_2021_40), the Research Ethics Committee/Committee of Ethics of Research with Medicines of Hospital Universitario de Gran Canaria Dr. Negrín (2019-031-1), and the research committees of all participating centers.

### Consent for publication

Not applicable.

### Availability of data and materials

Individual-level genetic and phenotype data cannot publicly available due to privacy or ethical restrictions. Whole-exome sequence data are available for request in EGA (Dataset ID: EGAD50000001613). GWAS and multi-trait summary data are available for requesters following the instructions provided at https://github.com/genomicsITER/HGinfections.

### Authors’ contributions

ES-P, LAR-R, ET-H, MR-F, SG-B, DJ, AM-B, AC, RG-M, JM.L-S, TH-B, MIG-L, and BG-G contributed to analyses, and experiments; TH-B, AC, EE, DD, JV, BG-G, and CF participated in data collection and clinical data; ES-P, SG-B, and CF contributed figures and interpretation of data; ES-P and CF designed and supervised the study; ES-P, and CF wrote the first draft of the manuscript; MIG-L and CF obtained funding. All authors read and approved the final manuscript.

## Acknowledgements

We would like to thank CEGEN-FPGMX for providing the genotyping service.

## References

1. Bos LDJ, Ware LB. Acute respiratory distress syndrome: causes, pathophysiology, and phenotypes. The Lancet. 2022 Oct;400(10358):1145–56.

2. ARDS Definition Task Force, Ranieri VM, Rubenfeld GD, Thompson BT, Ferguson ND, Caldwell E, et al. Acute respiratory distress syndrome: the Berlin Definition. JAMA. 2012 Jun 20;307(23):2526–33.

3. Matthay MA, Arabi Y, Arroliga AC, Bernard G, Bersten AD, Brochard LJ, et al. A New Global Definition of Acute Respiratory Distress Syndrome. Am J Respir Crit Care Med. 2024 Jan 1;209(1):37–47.

4. Ma W, Tang S, Yao P, Zhou T, Niu Q, Liu P, et al. Advances in acute respiratory distress syndrome: focusing on heterogeneity, pathophysiology, and therapeutic strategies. Signal Transduct Target Ther. 2025 Mar 7;10(1):75.

5. Ge R, Wang F, Peng Z. Advances in Biomarkers for Diagnosis and Treatment of ARDS. Diagnostics. 2023 Oct 24;13(21):3296.

6. van der Zee P, Rietdijk W, Somhorst P, Endeman H, Gommers D. A systematic review of biomarkers multivariately associated with acute respiratory distress syndrome development and mortality. Crit Care. 2020 Dec 24;24(1):243.

7. Villar J, Herrán-Monge R, González-Higueras E, Prieto-González M, Ambrós A, Rodríguez-Pérez A, et al. Clinical and biological markers for predicting ARDS and outcome in septic patients. Sci Rep. 2021 Nov 22;11(1):22702.

8. Chakradhar A, Baron RM, Vera MP, Devarajan P, Chawla L, Hou PC. Plasma renin as a novel prognostic biomarker of sepsis-associated acute respiratory distress syndrome. Sci Rep. 2024 Mar 20;14(1):6667.

9. Liao SY, Casanova NG, Bime C, Camp SM, Lynn H, Garcia JGN. Identification of early and intermediate biomarkers for ARDS mortality by multi-omic approaches. Sci Rep. 2021 Sep 23;11(1):18874.

10. Mu Z, Wei W, Fair B, Miao J, Zhu P, Li YI. The impact of cell type and context-dependent regulatory variants on human immune traits. Genome Biol. 2021 Apr 29;22(1):122.

11. Burnham KL, Milind N, Lee W, Kwok AJ, Cano-Gamez K, Mi Y, et al. eQTLs identify regulatory networks and drivers of variation in the individual response to sepsis. Cell Genomics. 2024 Jul;4(7):100587.

12. Guillen-Guio B, Lorenzo-Salazar JM, Ma SF, Hou PC, Hernandez-Beeftink T, Corrales A, et al. Sepsis-associated acute respiratory distress syndrome in individuals of European ancestry: a genome-wide association study. Lancet Respir Med. 2020 Mar;8(3):258–66.

13. Suarez-Pajes E, Shrine N, Tosco-Herrera E, Hernandez-Beeftink T, Rubio-Rodríguez LA, García-Laorden MI, et al. Genetic regulation of the vascular endothelial growth factor receptor 1 during sepsis and association with ARDS susceptibility. 2025.

14. Guillen-Guio B, Suarez-Pajes E, Tosco-Herrera E, Hernandez-Beeftink T, Lorenzo-Salazar JM, Chang D, et al. Genome-wide association study of susceptibility to acute respiratory distress syndrome. EBioMedicine. 2025 Oct;120:105951.

15. Hernandez-Beeftink T, Guillen-Guio B, Lorenzo-Salazar JM, Corrales A, Suarez-Pajes E, Feng R, et al. A genome-wide association study of survival in patients with sepsis. Crit Care. 2022 Nov 5;26(1):341.

16. Turley P, Walters RK, Maghzian O, Okbay A, Lee JJ, Fontana MA, et al. Multi-trait analysis of genome-wide association summary statistics using MTAG. Nat Genet. 2018 Feb 1;50(2):229–37.

17. Chang CC, Chow CC, Tellier LC, Vattikuti S, Purcell SM, Lee JJ. Second-generation PLINK: rising to the challenge of larger and richer datasets. Gigascience. 2015 Dec 25;4(1):7.

18. Rentzsch P, Schubach M, Shendure J, Kircher M. CADD-Splice—improving genome-wide variant effect prediction using deep learning-derived splice scores. Genome Med. 2021 Dec 22;13(1):31.

19. Tosco-Herrera E, Rubio-Rodríguez LA, Muñoz-Barrera A, Jáspez D, Suárez-Pajes E, Corrales A, et al. Rare genetic variant risks in patients with sepsis-associated acute respiratory distress syndrome. 2025.

20. Lee S, Emond MJ, Bamshad MJ, Barnes KC, Rieder MJ, Nickerson DA, et al. Optimal Unified Approach for Rare-Variant Association Testing with Application to Small-Sample Case-Control Whole-Exome Sequencing Studies. The American Journal of Human Genetics. 2012 Aug;91(2):224–37.

21. Słomian D, Szyda J, Dobosz P, Stojak J, Michalska-Foryszewska A, Sypniewski M, et al. Better safe than sorry—Whole-genome sequencing indicates that missense variants are significant in susceptibility to COVID-19. PLoS One. 2023 Jan 20;18(1):e0279356.

22. Kamiza AB, Touré SM, Zhou F, Soremekun O, Cissé C, Wélé M, et al. Multi-trait discovery and fine-mapping of lipid loci in 125,000 individuals of African ancestry. Nat Commun. 2023 Sep 5;14(1):5403.

23. Chen X, Zhu Y, Zhong P, Xie G. Multi-trait GWAS identifies pleiotropic loci associated with colorectal cancer in East Asian populations. Front Genet. 2025 Apr 15;16.

24. Cárcel-Márquez J, Muiño E, Gallego-Fabrega C, Cullell N, Lledós M, Llucià-Carol L, et al. A Polygenic Risk Score Based on a Cardioembolic Stroke Multitrait Analysis Improves a Clinical Prediction Model for This Stroke Subtype. Front Cardiovasc Med. 2022 Jul 8;9.

25. Meng Z, Zhang C, Liu S, Li W, Wang Y, Zhang Q, et al. Exploring genetic loci linked to COVID-19 severity and immune response through multi-trait GWAS analyses. Front Genet. 2025 Feb 17;16.

26. Cao M, Wang G, Xie J. Immune dysregulation in sepsis: experiences, lessons and perspectives. Cell Death Discov. 2023 Dec 19;9(1):465.

27. Habimana R, Choi I, Cho HJ, Kim D, Lee K, Jeong I. Sepsis-induced cardiac dysfunction: a review of pathophysiology. Acute and Critical Care. 2020 May 31;35(2):57–66.

28. Uhl GR, Martinez MJ. PTPRD: neurobiology, genetics, and initial pharmacology of a pleiotropic contributor to brain phenotypes. Ann N Y Acad Sci. 2019 Sep 15;1451(1):112–29.

29. Veeriah S, Brennan C, Meng S, Singh B, Fagin JA, Solit DB, et al. The tyrosine phosphatase PTPRD is a tumor suppressor that is frequently inactivated and mutated in glioblastoma and other human cancers. Proceedings of the National Academy of Sciences. 2009 Jun 9;106(23):9435–40.

30. Peyser ND, Du Y, Li H, Lui V, Xiao X, Chan TA, et al. Loss-of-Function PTPRD Mutations Lead to Increased STAT3 Activation and Sensitivity to STAT3 Inhibition in Head and Neck Cancer. PLoS One. 2015 Aug 12;10(8):e0135750.

31. Huang X, Qin F, Meng Q, Dong M. Protein tyrosine phosphatase receptor type D (PTPRD)—mediated signaling pathways for the potential treatment of hepatocellular carcinoma: a narrative review. Ann Transl Med. 2020 Sep;8(18):1192–1192.

32. AL-Eitan L. PTPRD gene variant rs10739150: A potential game-changer in hypertension diagnosis. PLoS One. 2024 Jun 27;19(6):e0304950.

33. Chen YT, Lin WD, Liao WL, Lin YJ, Chang JG, Tsai FJ. PTPRD silencing by DNA hypermethylation decreases insulin receptor signaling and leads to type 2 diabetes. Oncotarget. 2015 May 30;6(15):12997–3005.

34. Gong Y, McDonough CW, Beitelshees AL, Rouby N El, Hiltunen TP, O’Connell JR, et al. PTPRD gene associated with blood pressure response to atenolol and resistant hypertension. J Hypertens. 2015 Nov;33(11):2278–85.

35. Tsai FJ, Yang CF, Chen CC, Chuang LM, Lu CH, Chang CT, et al. A Genome-Wide Association Study Identifies Susceptibility Variants for Type 2 Diabetes in Han Chinese. PLoS Genet. 2010 Feb 19;6(2):e1000847.

36. Wu KCH, He Q, Bennett AN, Li J, Chan KHK. Shared genetic mechanism between type 2 diabetes and COVID-19 using pathway-based association analysis. Front Genet. 2022 Nov 22;13.

37. Xu J, Zhong Y, Yin H, Linneman J, Luo Y, Xia S, et al. Methylation-mediated silencing of PTPRD induces pulmonary hypertension by promoting pulmonary arterial smooth muscle cell migration via the PDGFRB/PLCγ1 axis. J Hypertens. 2022 Sep;40(9):1795–807.

38. Revercomb L, Hanmandlu A, Wareing N, Akkanti B, Karmouty-Quintana H. Mechanisms of Pulmonary Hypertension in Acute Respiratory Distress Syndrome (ARDS). Front Mol Biosci. 2021 Jan 18;7.

39. Moloney ED, Evans TW. Pathophysiology and pharmacological treatment of pulmonary hypertension in acute respiratory distress syndrome. European Respiratory Journal. 2003 Apr;21(4):720–7.

40. Li F, Zhang W, Wang M, Jia P. IL1RAP regulated by PRPRD promotes gliomas progression via inducing neuronal synapse development and neuron differentiation in vitro. Pathol Res Pract. 2020 Nov;216(11):153141.

41. Funato K, Yamazumi Y, Oda T, Akiyama T. Tyrosine phosphatase PTPRD suppresses colon cancer cell migration in coordination with CD44. Exp Ther Med. 2011;2(3):457–63.

42. Hillmer EJ, Zhang H, Li HS, Watowich SS. STAT3 signaling in immunity. Cytokine Growth Factor Rev. 2016 Oct;31:1–15.

43. Lin S, Yan J, Wang W, Luo L. STAT3-Mediated Ferroptosis is Involved in Sepsis-Associated Acute Respiratory Distress Syndrome. Inflammation. 2024 Aug 18;47(4):1204–19.

44. Zhao J, Yu H, Liu Y, Gibson SA, Yan Z, Xu X, et al. Protective effect of suppressing STAT3 activity in LPS-induced acute lung injury. American Journal of Physiology-Lung Cellular and Molecular Physiology. 2016 Nov 1;311(5):L868–80.

45. Liu Y, Zhao C, Cui L, Chen Y, Peng H, Liu J, et al. Transcription factors FOXN1 and intrathymic cytokines contribute to dysfunction of TECs during lipopolysaccharide-induced thymic atrophy. Int Immunopharmacol. 2025 Jun;157:114791.

46. Gallo V, Cirillo E, Giardino G, Pignata C. FOXN1 Deficiency: from the Discovery to Novel Therapeutic Approaches. J Clin Immunol. 2017 Nov 21;37(8):751–8.

47. Poli MC, Aksentijevich I, Bousfiha AA, Cunningham-Rundles C, Hambleton S, Klein C, et al. Human inborn errors of immunity: 2024 update on the classification from the International Union of Immunological Societies Expert Committee. Journal of Human Immunity. 2025 May 5;1(1).

48. Szczot M, Nickolls AR, Lam RM, Chesler AT. The Form and Function of PIEZO2. Annu Rev Biochem. 2021 Jun 20;90(1):507–34.

49. Xiong H, Yang Y, Yang J, Zhu Y, Ping F, He X, et al. Piezo2 as a novel biomarker of acute respiratory distress syndrome severity: a prospective observational study. Respir Med. 2025 Nov;248:108382.

50. Xu JY, Liu AR, Wu ZS, Xie JF, Qu XX, Li CH, et al. Nucleotide polymorphism in ARDS outcome: a whole exome sequencing association study. Ann Transl Med. 2021 May;9(9):780–780.

51. Wei F, Lin Z, Lu W, Luo H, Feng H, Liu S, et al. Deficiency of Endothelial *Piezo2* Impairs Pulmonary Vascular Angiogenesis and Predisposes Pulmonary Hypertension. Hypertension. 2025 Apr;82(4):583–97.

52. Tipoe TL, Wu WKK, Chung L, Gong M, Dong M, Liu T, et al. Plasminogen Activator Inhibitor 1 for Predicting Sepsis Severity and Mortality Outcomes: A Systematic Review and Meta-Analysis. Front Immunol. 2018 Jun 18;9.

