## Supplementary Material for "Protein-based genomic analysis for the identification of risk loci associated with acute respiratory distress syndrome"

***GEN-SEP study investigators**:

Miryam Prieto-González[1], Aurelio Rodríguez-Pérez[2,3], Demetrio Carriedo[4], Jesús Blanco[5,6], Alfonso Ambrós[7], Elena González-Higueras[8], Elena Espinosa[9], Arturo Muriel-Bombin[6], David Domínguez[9], Abelardo García de Lorenzo[10], José M. Añon[5,10], Marina Soro[11], Jesús Villar[5,12,13,14], Carlos Flores[5,15,16,17]

**GEN-SEP study affiliations:**

1. Intensive Care Unit, Complejo Asistencial Universitario de Palencia, Palencia, Spain.

2. Department of Anesthesiology, Hospital Universitario de Gran Canaria Dr. Negrín, Las Palmas de Gran Canaria, Spain.

3. Department of Medical and Surgical Sciences, University of Las Palmas de Gran Canaria, Las Palmas de Gran Canaria, Spain.

4. Intensive Care Unit, Complejo Hospitalario Universitario de León, León, Spain

5. CIBER de Enfermedades Respiratorias (CIBERES), Instituto de Salud Carlos III, Madrid, Spain.

6. Intensive Care Unit, Hospital Universitario Rio Hortega, Valladolid, Spain.

7. Intensive Care Unit, Hospital General de Ciudad Real, Ciudad Real, Spain.

8.Intensive Care Unit, Hospital Virgen de la Luz, Cuenca, Spain.

9. Department of Anesthesiology, Hospital Universitario Nuestra Señora de Candelaria, Santa Cruz de Tenerife, Spain.

10. Intensive Care Unit, Hospital Universitario La Paz, IdiPAZ, Madrid, Spain.

11. Anesthesiology and Critical Care Department, Hospital IMED Valencia, Valencia, Spain.

12. Research Unit, Hospital Universitario de Gran Canaria Dr. Negrín, Instituto de Investigación Sanitaria de Canarias (IISC), Las Palmas de Gran Canaria, Spain.

13. Faculty of Health Sciences, Universidad del Atlántico Medio, Las Palmas, Spain.

14. Li Ka Shing Knowledge Institute, St. Michael Hospital, Toronto, Canada.

15. Research Unit, Hospital Universitario Nuestra Señora de Candelaria, Instituto de Investigación Sanitaria de Canarias (IISC), Santa Cruz de Tenerife, Spain.

16. Genomics Division, Instituto Tecnológico y de Energías Renovables (ITER), Santa Cruz de Tenerife, Spain.

17. Facultad de Ciencias de la Salud, Universidad Fernando Pessoa Canarias, Las Palmas de Gran Canaria, Spain.

### Supplementary Methods

#### Study description

We included 221 samples from the GEN-SEP cohort, a multicenter observational study conducted across a national network of intensive care units (ICUs) in Spain, for which serum measurements of 10 biomarkers and SNP genotyping array data were available (1). These proteins were selected due to their roles in alveolar epithelial damage, endothelial injury, inflammation, tissue repair, and fibrinolysis (1,2). The cohort included adult patients (>18 years) primarily of European ancestry with a sepsis diagnosis, defined according to the Third International Consensus Definitions for Sepsis (3). Recruitment was conducted in two periods: 2002–2015 and 2016–2019.

#### Protein measurements

Serum samples were collected at three different time points after sepsis diagnosis. T1: 24 hours after sepsis diagnosis; T2: 48-72 hours after sepsis diagnosis; T7: 7 days after sepsis diagnosis. Serum protein measurements for Ang-2, AREG, CXCL16, ICAM-1, IL-1RA, IL-18, PAI-1, RAGE, and SP-D, were obtained by ELISA using DuoSet ELISA kits and the DuoSet Ancillary Reagent Kit 2 (R&D Systems, Abingdon, UK) as described elsewhere in (1). In addition, we included measurement of CXCL12 using the same methodology, which shows a limit of detection of 31.2 pg/mL. Data were normalized using Box-Cox transformations, and the normality of the data was tested using the Lilliefors test, using *car* and *nortest* R packages respectively. Outliers were removed when normality was not achieved.

#### SNP genotyping, variant imputation and GWAS of biomarkers

Genotyping was conducted on blood samples collected at T1, using the Axiom Genome-Wide Human CEU 1 Array (Thermo Fisher Scientific) in the National Genotyping Center (CeGen), Universidad de Santiago de Compostela Node.

Variant calling was performed using AffyPipe v.2.10.0 according to the manufacturer's recommendations. Quality controls were performed using PLINK v.1.9 (4) and R v.3.6.0. Variants with call rate (CR) <95%, minor allele frequency (MAF) <1%, or that strongly deviated from the Hardy-Weinberg equilibrium (HWE) expectations ($p$<1.0x10$^{-6}$), were excluded. Patients with missing clinical information, CR<95%, sex discordance, a high kinship degree (PIHAT>0.2), or that were heterozygosity outliers were excluded from the analysis. Variant imputation was performed with the Michigan Imputation Server using the Haplotype Reference Consortium (HRC) panel v.1.1 (5).

For the GWAS of protein biomarkers, we tested the association of the imputed variants with MAF>1% and imputation quality Rsq >0.3 with the levels of each biomarker for each collection time using the Wald test implemented in EPACTS (6).

After GWAS and multi-trait associations, sentinel variants were identified as those surpassing the significance threshold ($p$=5.0x$10^{-8}$) and showing weak linkage disequilibrium (LD; $r^2$ < 0.1) with other variants in each locus after clumping in PLINK v 1.9 (4).

### Bayesian fine-mapping, gene prioritization, and functional analyses

We performed a Bayesian fine-mapping around the sentinel variants identified in the GWAS and multi-trait analyses to identify the pQTLs most likely driving the associations. We used the R package *corrcoverage* to identify the credible sets containing the causal variant with 95% confidence (7).

To predict the biological impact of the variants included in the credible sets, variants were annotated using Variant Effect Predictor (VEP) v.105 to obtain the Combined Annotation Dependent Depletion (CADD) score v1.6. To derive the gene set, we prioritized genes based on functional evidence integrated into the Variant-to-Gene (V2G) score from Open Targets Genetics (https://platform.opentargets.org/, release 24.09, updated September 2024). We selected the gene with the highest V2G score linked to the variant with the greatest biological impact (highest CADD). In addition, we included the gene closest to the sentinel variants in the set.

We used different in silico tools and databases to evaluate the biological impact of genes in the context of ARDS. We relied on the Open Targets Platform to evaluate previous genetic associations with traits using the "GWAS associations" score (https://platform.opentargets.org/, release 25.06, updated June 2025). This score aggregate information of significant GWAS association and functional genomics to report target-trait relationships. To summarize these results, the traits were classified into 20 categories: autoimmune diseases, circulatory system, dermatologic, digestive, drug response, endocrine/metabolic, genitourinary, hematopoietic, infectious diseases, injuries & poisonings, lipid metabolism, measurement of proteins/metabolites, mental disorders, musculoskeletal, neoplasms/cancer, neurological, pregnancy complications, respiratory, sense organs, and others. In addition, we screened OMIM to determine whether the genes contained in the set had previously been associated with Mendelian diseases (https://www.omim.org/).

### Colocalization

To assess whether the variants associated with biomarker levels also influence the expression of other genes, we evaluated the colocalization between the significant loci, both from GWAS and multi-trait analyses, and expression quantitative trait loci (eQTLs) data from GTEx v8 (https://gtexportal.org/). For that, we included all genes for which the significant variant was an eQTL at nominal level ($p<0.05$). We then used *coloc* and evaluated the following hypotheses (8): H0: variants showed no association with biomarker levels, nor with eQTL data; H1: variants were only associated with biomarker levels; H2: variants were only associated with gene expression; H3: variants were associated with biomarker levels and gene expression, but by different causal SNP; H4: variants were associated with biomarker levels and gene expression by the same causal variant. Colocalization was declared when the posterior probability (PP) of H4 was > 80%.

### Whole-exome sequencing and association of rare variants

We accessed whole-exome sequencing (WES) data from 822 patients of the GEN-SEP study. Sequencing libraries, procedures, and quality controls are described elsewhere (9–11).

We functionally annotated the variants at prioritized genes using Ensembl VEP v105 and ANNOVAR v07.06.20 (https://annovar.openbioinformatics.org/en/latest/). Annotations included population allele frequency (AF) in 1000 Genomes Phase 3 Europeans (1KG) population, and AF in gnomAD v.2.1 non-Finnish Europeans populations, variant type, protein function, pathogenic potential, and the CADD. In addition, the mutation significance cutoff (MSC) scores with 95% confidence were annotated per gene (12).

Associations were tested using two variant categories according to AF on 1KG and gnomAD: i) variants with AF<0.001 or no data available; ii) variants with AF<0.001 and with high biological impact (CADD score > MSC). Rare variant association was performed in EPACTS using a SKAT-O test, and adjusted by sex, age, and the APACHE II (Acute Physiology and Chronic Health Evaluation II) score. The significance threshold was declared after Bonferroni correction considering the number of genes and the two variant categories tested ($p=6.25 \times 10^{-4}$).

### Polygenic scores

We evaluated the association of polygenic scores (PGS) for the 10 biomarkers with susceptibility to ARDS, multiple organ failure (MOF), and ICU mortality in patients with sepsis. The PGS were calculated with PRSice 2.3.5 (13) (R 4.2.2), using the biomarker GWAS summary statistics as base data. For each biomarker, we derived multiple PGS following an approach of thresholding and clumping ($r^2$ = 0.1 in 250 kb windows), excluding palindromic variants (A/T, C/G) from the analyses. PGS models were then tested on a target dataset of 621 independent patients from the GEN-SEP study to calculate individual PGS and test their association with sepsis-associated ARDS (237 cases and 384 controls), MOF (491 cases and 130 controls), and ICU mortality (196 cases and 414 controls). The individual PGS were normalized following the formula:

$$PGS_j = \frac{\sum_i(\beta ixGij) - Mean(PGS)}{SD(PGS)}$$

where β is the logarithm of the odds ratio, G is the genotype of variant i in individual j, and SD is the standard deviation of the PGS. For each biomarker, we selected the best fitting and the significance was declared at $p$=5.56x10$^{-4}$ to control the number of independent tests (0.05/90). All PGS were adjusted by sex, age, and APACHE II score.

Finally, we performed a pathway-based PGS analysis using PRSet (14) implemented in PRSice v2.3.5. Unlike traditional PGS approaches, which can aggregate genetic effects genome-wide, PRSet groups variants according to predefined biological pathways. Pathway-specific scores were calculated only for biomarkers and outcomes that met significance in the PGS models. In this regard, we evaluated the biological mechanisms linking the PAI-1 biomarker levels at T7 to ICU mortality. The GWAS summary statistics of PAI-1 biomarker levels at T7 was used as the base dataset. Pathway-specific scores were then tested for association with ICU mortality in an independent target dataset of 610 patients from the GEN-SEP study. The pathways used were derived from the Reactome database (https://reactome.org/), including 2,725 pathways in the analysis. A GTF file for the GRCh37 genome build was used to map the SNPs to genes. The individual pathways scores were normalized using z-score standardization, and association model was adjusted by sex, age, and APACHE II score. The significance threshold for this analysis was established at $p$= 1.83x10$^{-5}$ after Bonferroni correction (0.05/2,725).

### Supplementary References:

1. Singer M, Deutschman CS, Seymour CW, Shankar-Hari M, Annane D, Bauer M, et al. The Third International Consensus Definitions for Sepsis and Septic Shock (Sepsis-3). *JAMA*. 2016 Feb 23;315(8):801.

2. Villar J, Herrán-Monge R, González-Higueras E, Prieto-González M, Ambrós A, Rodríguez-Pérez A, et al. Clinical and biological markers for predicting ARDS and outcome in septic patients. *Sci Rep*. 2021 Nov 22;11(1):22702.

3. Chang CC, Chow CC, Tellier LC, Vattikuti S, Purcell SM, Lee JJ. Second-generation PLINK: rising to the challenge of larger and richer datasets. *Gigascience*. 2015 Dec 25;4(1):7.

4. Loh PR, Danecek P, Palamara PF, Fuchsberger C, A Reshef Y, K Finucane H, et al. Reference-based phasing using the Haplotype Reference Consortium panel. *Nat Genet*. 2016 Nov 3;48(11):1443–8.

5. EPACTS: Efficient and Parallelizable Association Container Toolbox. (http://genome.sph.umich.edu/wiki/EPACTS).

6. Hutchinson A, Watson H, Wallace C. Improving the coverage of credible sets in Bayesian genetic fine-mapping. *PLoS Comput Biol*. 2020 Apr 13;16(4):e1007829.

7. Giambartolomei C, Vukcevic D, Schadt EE, Franke L, Hingorani AD, Wallace C, et al. Bayesian Test for Colocalisation between Pairs of Genetic Association Studies Using Summary Statistics. *PLoS Genet*. 2014 May 15;10(5):e1004383.

8. Tosco-Herrera E, Rubio-Rodríguez LA, Muñoz-Barrera A, Jáspez D, Suárez-Pajes E, Corrales A, et al. Rare genetic variant risks in patients with sepsis-associated acute respiratory distress syndrome. 2025. *medRxiv*.

9. Guillen-Guio B, Suarez-Pajes E, Tosco-Herrera E, Hernandez-Beeftink T, Lorenzo-Salazar JM, Chang D, et al. Genome-wide association study of susceptibility to acute respiratory distress syndrome. *EBioMedicine*. 2025 Oct;120:105951.

10. Suarez-Pajes E, Shrine N, Tosco-Herrera E, Hernandez-Beeftink T, Rubio-Rodríguez LA, García-Laorden MI, et al. Genetic regulation of the vascular endothelial growth factor receptor 1 during sepsis and association with ARDS susceptibility. 2025. *medRxiv*.

11. Itan Y, Shang L, Boisson B, Ciancanelli MJ, Markle JG, Martinez-Barricarte R, et al. The mutation significance cutoff: gene-level thresholds for variant predictions. *Nat Methods*. 2016 Feb 28;13(2):109–10.

12. Choi SW, O'Reilly PF. PRSice-2: Polygenic Risk Score software for biobank-scale data. *Gigascience*. 2019 Jul 1;8(7).

13. Choi SW, García-González J, Ruan Y, Wu HM, Porras C, Johnson J, et al. PRSet: Pathway-based polygenic risk score analyses and software. *PLoS Genet*. 2023 Feb 7;19(2):e1010624.

### Supplementary Tables

**Table S1. Normalized biomarker concentration (pg/ml) for individuals included in each association study.**

| Biomarker | T1 | T2 | T7 |
|---|---|---|---|
| **Ang-2** | n=204<br>9.63 ± 1.50 | n=125<br>8.46 ± 1.28 | n=109<br>13.38 ± 2.70 |
| **AREG** | n=160<br>1.85 ± 0.14 | n=80<br>1.82 ± 0.15 | n=65<br>1.63 ± 0.10 |
| **CXCL12** | n=135<br>17.52 ± 2.83 | n=128<br>64.25±16.61 | n=113<br>22.74 ± 4.19 |
| **CXCL16** | n=203<br>4.16±0.11 | n=122<br>5.28±0.22 | n=108<br>3.09 ± 0.05 |
| **ICAM-1** | n=204<br>16.08 ± 0.89 | n=125<br>8.41 ± 0.25 | n=109<br>38.92 ± 3.77 |
| **IL-1RA** | n=135<br>5.04 ± 0.86 | n=46<br>3.91±0.81 | n=29<br>3.75 ± 0.87 |
| **IL-18** | n=204<br>4.08 ± 0.41 | n=125<br>4.41 ± 0.52 | n=109<br>4.59 ± 0.54 |
| **PAI-1** | n=204<br>2.65 ± 0.33 | n=125<br>3.10 ± 0.62 | n=106<br>3.58 ± 0.76 |
| **RAGE** | n=204<br>5.75 ± 0.66 | n=125<br>4.49 ± 0.43 | n=109<br>3.58 ± 0.24 |
| **SP-D** | n=204<br>10.47 ± 1.92 | n=125<br>20.46 ± 5.56 | n=109<br>10.13 ± 1.64 |

T1: within 24 hours after sepsis diagnosis; T2: 48-72 hours after sepsis diagnosis; T7: 7 days after sepsis diagnosis.

**Table S2. Demographic and clinical features of GEN-SEP patients after quality controls.**

| | Patients with protein measurement (n=209) | Independent patients for PGS model association testing (n=621) | *p*-value |
|---|---|---|---|
| **Females, % (proportion)** | 39.71 (83/209) | 35.10 (218/621) | 0.265 |
| **Males, % (proportion)** | 60.29 (126/209) | 64.90 (403/621) | |
| **Age, mean years ± SD** | 63.43 ± 15.17 (209) | 63.42 ± 14.53 (621) | 0.930 |
| **BMI, mean kg/m2 ± SD** | 27.48+-9.44 (192) | 27.63 ± 13.70 (221) | 0.534 |
| **APACHE II, mean ± SD** | 16.88 ± 7.05 (206) | 20.36 ± 7.10 (621) | $1.62x10^{-10}$ |
| **Comorbidities[$], % (proportion)** | 75.11 (157/209) | 75.52 (469/621) | 0.981 |
| **MOF, % (proportion)** | 71.29 (149/209) | 79.07 (491/621) | 0.027 |
| **ARDS, % (proportion)** | 35.41 (74/209) | 38.16 (237/621) | 0.528 |
| **ICU mortality, % (proportion)** | 15.79 (33/209) | 32.13 (196/610) | $8.44x10^{-6}$ |
| **Sepsis of pulmonary origin, % (proportion)** | 36.59 (75/205) | 35.14 (195/555) | 0.775 |
| **Pathogen, % (proportion)** | | | |
| **Gram-positive** | 20.83 (40/192) | 31.22 (113/362) | 0.037 |
| **Gram-negative** | 35.42 (68/192) | 38.40 (139/362) | 0.944 |
| **Gram-positive and Gram-negative** | 13.54 (26/192) | 14.36 (52/362) | 1 |
| **Virus** | 6.77 (13/192) | 1.93 (7/362) | $5.00x10^{-3}$ |
| **Others[+]** | 9.38 (18/192) | 14.09 (51/362) | 0.272 |

[$]Comorbidities include cancer, age >80 years, hepatopathy, valvular disease, immunodeficiency, morbid obesity, chronic disease, pregnancy, autoimmune disease, ischemic cardiopathy, pneumonia, and serious recurrent infections. +Others include both fungi and polymicrobial. APACHE II, Acute Physiology and Chronic Health Evaluation II; ARDS, Acute Respiratory Distress Syndrome; BMI, Body Mass Index; ICU, Intensive Care Unit; PGS: polygenic scores.

**Table S3. Results of detailed Bayesian fine mapping analyses and variants with highest impact for GWAS and multi-trait significant loci.**

| | | | Highest biological impact variant | | | | | | | |
|---|---|---|---|---|---|---|---|---|---|---|
| **Biomarker** | **Sentinel variant (rsID)** | **CS** | **SNP** | **rsID** | **R2** | **PP** | **CADD** | **EAF** | ***p*-value** | **Beta(SE)** |
| **Ang-2 (T1)** | rs76099260* | 46 | 7:7080055:C:G | rs79660405 | 1 | 0.03 | 13.09 | 0.01 | $2.04x10^{-8}$ | -3.38(0.58) |
| | rs77880844 | 16 | 14:81745399:G:T | rs6574635 | 0.12 | 0.02 | 7.73 | 0.14 | $5.19x10^{-3}$ | 0.36(0.13) |
| **AREG (T2)** | rs7970411 | 79 | 12:125717261:T:G | rs11058091 | 0.96 | 0.02 | 11.05 | 0.04 | $3.05x10^{-8}$ | 1.66(0.30) |
| **AREG (T7)** | rs731153 | 16 | 2:240453552:T:C | rs7592100 | 0.29 | 0.02 | 6.08 | 0.42 | $2.33x10^{-5}$ | -0.59(0.14) |
| | rs12554442 | 28 | 9:8417661:C:T | rs10114603 | 0.33 | 0.01 | 12.75 | 0.15 | $9.22x10^{-7}$ | 0.93(0.19) |
| | rs78986031 | 36 | 9:28290685:A:G | rs4878276 | 0.17 | 0.01 | 10.56 | 0.11 | $6.26x10^{-3}$ | -0.60(0.22) |
| | rs10778455 | 11 | 12:106444919:C:T | rs4964434 | 0.96 | 0.07 | 5.71 | 0.45 | $1.09x10^{-7}$ | 0.73(0.14) |
| | rs9510995 | 2 | 13:24590355:G:A | rs9510995 | 1 | 0.48 | 0.89 | 0.29 | $3.22x10^{-8}$ | -0.83(0.15) |
| | rs118062838 | 22 | 18:58285144:G:A | rs79553803 | 0.40 | 0.05 | 11.92 | 0.07 | $3.65x10^{-5}$ | -1.11(0.27) |
| **CXCL16 (T1)** | rs7517208 | 32 | 1:114939079:A:G | rs41296188 | 0.63 | 0.001 | 14.90 | 0.06 | $8.18x10^{-5}$ | 0.69(0.18) |
| | rs73880250 | 61 | 3:172481895:G:A | rs76627568 | 0.59 | 0.01 | 9.92 | 0.05 | $1.34x10^{-6}$ | 0.94(0.20) |
| **ICAM1 (T2)** | rs148202393 | 8 | 1:46327833:G:C | rs4660894 | 0.46 | 0.17 | 11.06 | 0.02 | $2.38x10^{-6}$ | -1.56(0.33) |
| | rs726017 | 5 | 2:234732311:G:A | rs17868361 | 0.76 | 0.09 | 18.25 | 0.16 | $9.32x10^{-8}$ | -0.68(0.13) |
| | rs2220449 | 7 | 4:116945894:A:C | rs57771678 | 0.15 | 0.07 | 4.39 | 0.02 | 0.009 | -0.87(0.33) |
| **IL1RA (T7)** | rs35003 | 37 | 5:16616264:C:T | rs14153 | 0.27 | 0.004 | 15.24 | 0.34 | 0.157 | 0.40(0.29) |
| | rs11156083 | 32 | 6:156468407:C:G | rs6928850 | 0.56 | 0.01 | 21.50 | 0.36 | $1.98x10^{-4}$ | -1.05(0.28) |
| | rs11658852 | 4 | 17:15300324:G:A | rs11658852 | 1 | 0.42 | 8.80 | 0.48 | $9.56x10^{-10}$ | 1.66(0.27) |
| **IL18 (T1)** | rs140560482 | 4 | 5:94734878:T:C | rs79284594 | 0.80 | 0.04 | 15.71 | 0.02 | $2.17x10^{-5}$ | 1.35(0.32) |
| **IL18 (T2)** | rs7235298 | 22 | 18:11307242:T:G | rs2101977 | 0.34 | 0.01 | 11.78 | 0.02 | $1.40x10^{-6}$ | -1.60(0.33) |

| (Cont. Table S3) | | | Highest biological impact variant | | | | | | | |
|---|---|---|---|---|---|---|---|---|---|---|
| **Biomarker** | **Sentinel variant (rsID)** | **CS** | **SNP** | **rsID** | **R2** | **PP** | **CADD** | **EAF** | ***p*-value** | **Beta(SE)** |
| **IL18 (T7)** | rs62519200 | 12 | 8:125237307:C:A | rs12674849 | 0.28 | 0.01 | 8.64 | 0.30 | $4.62 \times 10^{-5}$ | -0.47(0.11) |
| **PAI-1 (T1)** | rs113037345* | 1 | 5:52217786:G:A | rs113037345 | 1 | 1 | 0.15 | 0.01 | $4.92 \times 10^{-8}$ | -0.73(0.13) |
| **RAGE (T2)** | rs28413183 | 12 | 21:45376739:A:G | rs3788092 | 0.14 | 0.06 | 7.74 | 0.45 | $2.58 \times 10^{-6}$ | -0.46(0.10) |
| **RAGE (T7)** | rs2476174 | 12 | 13:80307810:T:C | rs78031890 | 0.12 | 0.05 | 12.66 | 0.02 | 0.43 | 0.31(0.40) |
| | rs142329471 | 15 | 13:112875863:G:A | rs142329471 | 1 | 0.74 | 3.08 | 0.02 | $3.49 \times 10^{-8}$ | -2.20(0.40) |
| | rs117886541 | 2 | 17:26825894:T:G | rs79677589 | 0.10 | 0.33 | 14.86 | 0.10 | $3.85 \times 10^{-3}$ | -0.58(0.20) |
| **SPD (T1)** | rs62500471 | 29 | 8:53692261:A:T | rs3849809 | 0.64 | 0.003 | 10.67 | 0.23 | $1.44 \times 10^{-7}$ | -0.37(0.07) |
| **SPD (T7)** | rs62500471 | 58 | 8:53725030:G:T | rs62501739 | 0.65 | 0.002 | 14.29 | 0.22 | $9.32 \times 10^{-7}$ | -0.60(0.12) |

*Results from GWAS. CADD: Combined Annotation Dependent Depletion score (scaled) v.1.6, CS: number of variants in the credible set, EAF: effect allele frequency, PP: posterior probability, R2: linkage disequilibrium from the sentinel variant, SE: standard error, SNP: chromosome: position base (GRCh37/hg19): non-effect allele: effect allele. T1: within 24 hours after sepsis diagnosis; T2: 48-72 hours after sepsis diagnosis; T7: 7 days after sepsis diagnosis.

**Table S4. Variants included in credible sets with high biological impact (CADD>10.00) obtained in fine mapping analyses.**

| Biomarker | rsID | EAF | Beta(SE) | p-value | R2 | PP | Gene | CADD | AF |
|---|---|---|---|---|---|---|---|---|---|
| Ang-2* (T1) | rs79660405 | 0.01 | -3.38(0.58) | $2.04x10^{-8}$ | 1 | 0.030 | *CCZ1B\MIR3683* | 13.09 | 0.012 |
| | rs61739548 | 0.01 | -3.11(0.59) | $3.07x10^{-7}$ | 0.76 | 0.021 | *LOC100131257* (ncRNA-exonic) | 12.44 | 0.016 |
| | rs10272510 | 0.08 | -0.54(0.28) | 0.058 | 0.11 | 0.008 | *LOC100131257* (ncRNA-exonic) | 12.43 | 0.058 |
| | rs61742328 | 0.02 | -1.70(0.47) | $4.23x10^{-4}$ | 0.20 | 0.012 | *LOC100131257* (ncRNA-exonic) | 12.40 | 0.018 |
| | rs6970740 | 0.01 | -3.38(0.58) | $2.04x10^{-8}$ | 1 | 0.030 | *CCZ1B\MIR3683* | 11.79 | 0.012 |
| | rs10271901 | 0.08 | -0.54(0.28) | 0.058 | 0.11 | 0.008 | *LOC100131257* (ncRNA-exonic) | 10.77 | 0.058 |
| | rs77807109 | 0.10 | -0.71(0.26) | 0.007 | 0.11 | 0.016 | *LOC100131257\ C1GALT1* | 10.48 | 0.089 |
| AREG (T2) | rs11058091 | 0.04 | 1.66(0.30) | $3.05x10^{-8}$ | 0.96 | 0.015 | *TMEM132B* (intronic) | 11.05 | 0.05 |
| AREG (T7) | rs10114603 | 0.15 | 0.93(0.19) | $9.22x10^{-7}$ | 0.33 | 0.009 | *PTPRD* (intronic) | 12.75 | 0.13 |
| | rs4878276 | 0.11 | -0.60(0.22) | $6.26x10^{-3}$ | 0.17 | 0.008 | *LINGO2* (intronic) | 10.56 | 0.09 |
| | rs79553803 | 0.07 | -1.11(0.27) | $3.65x10^{-5}$ | 0.40 | 0.045 | *MC4R\CDH20* | 11.92 | 0.05 |
| CXCL16 (T1) | rs41296188 | 0.06 | 0.69(0.18) | $8.18x10^{-5}$ | 0.63 | 0.001 | *TRIM33* (UTR-3') | 14.90 | 0.04 |
| | rs139333764 | 0.05 | 0.79(0.19) | $2.08 x10^{-5}$ | 0.68 | 0.001 | *SYT6\TRIM33* | 13.66 | 0.04 |
| | rs77069792 | 0.06 | 0.69(0.18) | $8.18x10^{-5}$ | 0.63 | 0.001 | *SYT6\TRIM33* | 12.33 | 0.04 |
| | rs74115209 | 0.05 | 0.79(0.19) | $2.08x10^{-5}$ | 0.63 | 0.001 | *TRIM33* (intronic) | 11.35 | 0.03 |
| | rs74113167 | 0.06 | 0.69(0.18) | $8.18x10^{-5}$ | 0.63 | 0.001 | *SYT6\TRIM33* | 10.84 | 0.04 |
| ICAM1 (T2) | rs4660894 | 0.02 | -1.56(0.33) | $2.38x10^{-6}$ | 0.46 | 0.166 | *MAST2* (intronic) | 11.06 | 0.01 |
| | rs17868361 | 0.16 | -0.68(0.13) | $9.32x10^{-8}$ | 0.76 | 0.091 | *MROH2A* (exonic, splice donor) | 18.25 | 0.16 |
| | rs1500480 | 0.18 | 0.69(0.12) | $1.83x10^{-8}$ | 1 | 0.368 | *MROH2A* (exonic, missense) | 14.41 | 0.84 |
| IL-1RA (T7) | rs14153 | 0.34 | 0.40(0.29) | 0.157 | 0.27 | 0.004 | *LOC101929524* (ncRNA-exonic) | 15.24 | 0.55 |
| | rs35003 | 0.16 | 2.14(0.37) | $1.06x10^{-8}$ | 1 | 0.108 | LOC101929524 (ncRNA-exonic) | 11.46 | 0.78 |
| | rs6928850 | 0.36 | -1.05(0.28) | $1.98x10^{-4}$ | 0.56 | 0.009 | *MIR1202\ SNORD28B* | 21.50 | 0.30 |
| | rs2354393 | 0.28 | -1.63(0.30) | $7.33x10^{-8}$ | 0.89 | 0.076 | *MIR1202\ SNORD28B* | 16.44 | 0.22 |
| | rs6905034 | 0.17 | -1.23(0.36) | $6.23x10^{-4}$ | 0.25 | 0.004 | *MIR1202\ SNORD28B* | 15.32 | 0.21 |
| IL18 (T1) | rs79284594 | 0.02 | 1.35(0.32) | $2.17x10^{-5}$ | 0.80 | 0.039 | *FAM81B* (intronic) | 15.71 | 0.03 |
| IL18 (T2) | rs2101977 | 0.02 | -1.60(0.33) | $1.40x10^{-6}$ | 0.34 | 0.013 | *PIEZO2\ LINC01928* | 11.78 | 0.03 |

**(Cont. Table S4)**

| Biomarker | rsID | EAF | Beta(SE) | p-value | R2 | PP | Gene | CADD | AF |
|---|---|---|---|---|---|---|---|---|---|
| RAGE (T7) | rs78031890 | 0.02 | 0.31(0.40) | 0.43 | 0.12 | 0.05 | *LINC01068\ LINC01038* | 12.66 | 0.02 |
| | rs117886541 | 0.10 | -0.58(0.20) | $3.85x10^{-3}$ | 0.10 | 0.33 | *SLC13A2\ FOXN1* | 14.86 | 0.09 |
| SP-D (T1) | rs3849809 | 0.23 | -0.37(0.07) | $1.44x10^{-7}$ | 0.64 | 0.003 | *RB1CC1\ NPBWR1* | 10.67 | 0.22 |
| SP-D (T7) | rs62501739 | 0.22 | -0.60(0.12) | $9.32x10^{-7}$ | 0.65 | 0.002 | *RB1CC1\ NPBWR1* | 14.29 | 0.23 |
| | rs62501710 | 0.21 | -0.64(0.12) | $2.54x10^{-7}$ | 0.64 | 0.005 | *RB1CC1\ NPBWR1* | 10.97 | 0.22 |
| | rs3849809 | 0.21 | -0.64(0.12) | $2.54x10^{-7}$ | 0.64 | 0.005 | *RB1CC1\ NPBWR1* | 10.67 | 0.22 |
| | rs62501740 | 0.22 | -0.60(0.12) | $9.32x10^{-7}$ | 0.65 | 0.002 | *RB1CC1\ NPBWR1* | 10.17 | 0.23 |

*Results from GWAS. AF: allele frequency obtained for gnomAD non-Finnish European (nFE); CADD: Combined Annotation Dependent Depletion score (scaled) v.1.6; EAF: effect allele frequency; SE: standard error; PP: posterior probability; R2: linkage disequilibrium between variant with the sentinel pQTL; T1: within 24 hours after sepsis diagnosis; T2: 48-72 hours after sepsis diagnosis; T7: 7 days after sepsis diagnosis.

**Table S5. Gene prioritization on the basis of the Bayesian fine mapping.**

| Biomarker | Position | Closet gene(s) | Highest CADD | |
|---|---|---|---|---|
| | | | **Gene** | **Top rank V2G** |
| Ang-2 (T1) | 14:81683460 | *GTF2A1* | *GTF2A1* | 0.153 |
| | 7:7054699* | *CCZ1B\MIR3683* | *C1GALT1* | 0.053 |
| AREG (T2) | 12:125706376 | *TMEM132B* | *SCARB1* | 0.080 |
| | | | *TMEM132B* | 0.080 |
| AREG (T7) | 2:240458353 | *HDAC4-AS1\LOC150935* | *HDAC4* | 0.066 |
| | 9:8427879 | *PTPRD* | *PTPRD* | 0.020 |
| | 9:28255496 | *LINGO2* | *LINGO2* | 0.141 |
| | 12:106439029 | *CASC18\NUAK1* | *NUAK1* | 0.106 |
| | 13:24590355 | *SPATA13* | *SPATA13* | 0.133 |
| | 18:58133225 | *MC4R\CDH20* | *MC4R* | 0.033 |
| CXCL16 (T1) | 1:114915192 | *SYT6\TRIM33* | *DENND2C* | 0.114 |
| | 3:172441323 | *NCEH1\ECT2* | *NCEH1* | 0.100 |
| ICAM-1 (T2) | 1:46618112 | *P3R3URF-PIK3R3\PIK3R3* | *LURAP1* | 0.146 |
| | 2:234725819 | *MROH2A* | *MROH2A* | 0.475 |
| | 4:117336713 | *MIR1973\TRAM1L1* | NA | NA |
| IL-1RA (T7) | 5:16617099 | *LOC101929524* | *RETREG1* | 0.153 |
| | 6:156474813 | *MIR1202\SNORD28B* | NA | NA |
| | 17:15300324 | *TEKT3\TVP23C-CDRT4* | *PMP22* | 0.066 |
| IL-18 (T1) | 5:94675417 | *MCTP1\FAM81B* | *FAM81B* | 0.140 |
| IL-18 (T2) | 18:11263135 | *PIEZO2\LINC01928* | *PIEZO2* | 0.046 |
| IL-18 (T7) | 8:125239839 | *LOC101927588* | *TRMT12* | 0.046 |
| | | | *TMEM65* | 0.046 |
| PAI-1 (T1) | 5:52217786* | *ITGA1* | *ITGA1* | 0.126 |
| RAGE (T2) | 21:45471684 | *TRAPPC10* | *PDXK* | 0.139 |
| RAGE (T7) | 13:80174234 | *LINC01068\LINC01038* | *NDFIP2* | 0.033 |
| | 13:112875863 | *LINC01070\LOC101928730* | *SPACA7* | 0.046 |
| | | | *SOX1* | 0.046 |
| | 17:26831908 | *SLC13A2\FOXN1* | *RSKR* | 0.113 |
| SP-D (T1) | 8:53777234 | *RB1CC1*\NPBWR1 | *RB1CC1* | 0.060 |
| SP-D (T7) | 8:53777234 | *RB1CC1\NPBWR1* | *RB1CC1* | 0.060 |

*Results from GWAS. CADD: Combined Annotation Dependent Depletion score (scaled) v.1.6; V2G: Variant-to-Gene score from (https://platform.opentargets.org/). T1: within 24 hours after sepsis diagnosis; T2: 48-72 hours after sepsis diagnosis; T7: 7 days after sepsis diagnosis.

**Table S6. Description of the prioritized genes.**

| Symbol | Description | Category |
|---|---|---|
| *C1GALT1* | Core 1 Synthase, Glycoprotein-N-Acetylgalactosamine 3-Beta-Galactosyltransferase | Protein Coding |
| *CASC18* | Cancer Susceptibility 18 | RNA Gene (lncRNA) |
| *CCZ1B* | CCZ1 Homolog B, Vacuolar Protein Trafficking And Biogenesis Associated | Protein Coding |
| *CDH20* | Cadherin 20 | Protein Coding |
| *DENND2C* | DENN Domain Containing 2C | Protein Coding |
| *ECT2* | Epithelial Cell Transforming 2 | Protein Coding |
| *FAM81B* | Family With Sequence Similarity 81 Member B | Protein Coding |
| *FOXN1* | Forkhead Box N1 | Protein Coding |
| *GTF2A1* | General Transcription Factor IIA Subunit 1 | Protein Coding |
| *HDAC4* | Histone Deacetylase 4 | Protein Coding |
| *HDAC4-AS1* | HDAC4 Antisense RNA | RNA Gene (lncRNA) |
| *ITGA1* | Integrin Subunit Alpha 1 | Protein Coding |
| *LINC01038* | Long Intergenic Non-Protein Coding RNA 1038 | RNA Gene (lncRNA) |
| *LINC01068* | Long Intergenic Non-Protein Coding RNA 1068 | RNA Gene (lncRNA) |
| *LINC01070* | Long Intergenic Non-Protein Coding RNA 1070 | RNA Gene (lncRNA) |
| *LINC01928* | Long Intergenic Non-Protein Coding RNA 1928 | RNA Gene (lncRNA) |
| *LINGO2* | Leucine Rich Repeat And Ig Domain Containing 2 | Protein Coding |
| *LOC101927588* | Uncharacterized LOC101927588 | RNA Gene (lncRNA) |
| *LOC101928730* | Uncharacterized LOC101928730 | RNA Gene (ncRNA) |
| *LOC101929524* | Uncharacterized LOC101929524 | RNA Gene |
| *LOC150935* | Uncharacterized LOC150935 | RNA Gene (lncRNA) |
| *LURAP1* | Leucine Rich Adaptor Protein 1 | Protein Coding |
| *MC4R* | Melanocortin 4 Receptor | Protein Coding |
| *MCTP1* | Multiple C2 And Transmembrane Domain Containing 1 | Protein Coding |
| *MIR1202* | MicroRNA 1202 | RNA Gene (miRNA) |
| *MIR1973* | MicroRNA 1973 | RNA Gene (miRNA) |
| *MIR3683* | MicroRNA 3683 | RNA Gene (miRNA) |
| *MROH2A* | Maestro Heat Like Repeat Family Member 2A | Protein Coding |
| *NCEH1* | Neutral Cholesterol Ester Hydrolase 1 | Protein Coding |
| *NDFIP2* | Nedd4 Family Interacting Protein 2 | Protein Coding |
| *NUAK1* | NUAK Family Kinase 1 | Protein Coding |
| *PDXK* | Pyridoxal Kinase | Protein Coding |
| *P3R3URF-PIK3R3* | P3R3URF-PIK3R3 Readthrough | Protein Coding |
| *PIEZO2* | Piezo Type Mechanosensitive Ion Channel Component 2 | Protein Coding |
| *PIK3R3* | Phosphoinositide-3-Kinase Regulatory Subunit 3 | Protein Coding |
| *PMP22* | Peripheral Myelin Protein 22 | Protein Coding |
| *PTPRD* | Protein Tyrosine Phosphatase Receptor Type D | Protein Coding |
| *RB1CC1* | RB1 Inducible Coiled-Coil 1 | Protein Coding |
| *RETREG1* | Reticulophagy Regulator 1 | Protein Coding |
| *RSKR* | Ribosomal Protein S6 Kinase Related | Protein Coding |
| *SCARB1* | Scavenger Receptor Class B Member 1 | Protein Coding |
| *SLC13A2* | Solute Carrier Family 13 Member 2 | Protein Coding |

**(Cont. Table S6)**

| Symbol | Description | Category |
|---|---|---|
| *SNORD28B* | Small Nucleolar RNA, C/D Box 28B | RNA Gene (snoRNA) |
| *SOX1* | SRY-Box Transcription Factor 1 | Protein Coding |
| *SPACA7* | Sperm Acrosome Associated 7 | Protein Coding |
| *SPATA13* | Spermatogenesis Associated 13 | Protein Coding |
| *SYT6* | Synaptotagmin 6 | Protein Coding |
| *TEKT3* | Tektin 3 | Protein Coding |
| *TMEM132B* | Transmembrane Protein 132B | Protein Coding |
| *TMEM65* | Transmembrane Protein 65 | Protein Coding |
| *TRAM1L1* | Translocation Associated Membrane Protein 1 Like 1 | Protein Coding |
| *TRAPPC10* | Trafficking Protein Particle Complex Subunit 10 | Protein Coding |
| *TRIM33* | Tripartite Motif Containing 33 | Protein Coding |
| *TRMT12* | TRNA Wybutosine-Synthesizing Protein 2 | Protein Coding |
| *TVP23C-CDRT4* | TVP23C-CDRT4 Readthrough | Protein Coding |
| *NPBWR1* | Neuropeptides B And W Receptor 1 | Protein Coding |

**Table S7. Prioritized genes involved in Mendelian disorders.**

| Gene | Phenotype | Phenotype MIM number | Inheritance |
|---|---|---|---|
| *FOXN1* | T-cell immunodeficiency, congenital alopecia, and nail dystrophy | 601705 | AR |
| | T-cell lymphopenia, infantile, with or without nail dystrophy, autosomal dominant | 618806 | AD |
| *HDAC4* | Neurodevelopmental disorder with central hypotonia and dysmorphic facies | 619797 | AD |
| *MC4R* | Obesity, resistance to BMIQ20 | 618406 | AD, AR |
| | Obesity (BMIQ20) | 618406 | AD, AR |
| *PDXK* | Neuropathy, hereditary motor and sensory, type VIC, with optic atrophy | 618511 | AR |
| *PIEZO2* | Marden-Walker syndrome* | 248700 | AD |
| | Arthrogryposis, distal, type 3 | 114300 | AD |
| | Arthrogryposis, distal, type 5 | 108145 | AD |
| | Arthrogryposis, distal, with impaired proprioception and touch | 617146 | AR |
| *PMP22* | Neuropathy, inflammatory demyelinating | 139393 | AD* |
| | Charcot-Marie-Tooth disease, type 1A | 118220 | AD |
| | Charcot-Marie-Tooth disease, type 1E | 118300 | AD |
| | Dejerine-Sottas disease | 145900 | AD, AR |
| | Neuropathy, recurrent, with pressure palsies | 162500 | AD |
| | Roussy-Levy syndrome | 180800 | AD |
| *RB1CC1* | Breast cancer, somatic | 114480 | NA |
| *RETREG1* | Neuropathy, hereditary sensory and autonomic, type IIB | 613115 | AR |
| *SCARB1* | High density lipoprotein cholesterol level QTL6 | 610762 | NA |
| *TRAPPC10* | Neurodevelopmental disorder with microcephaly, short stature, and speech delay | 620027 | AR |
| *TRIM33* | Developmental dysplasia of the hip 4* | 621311 | NA |

*Provisional relationship. AD: Autosomal dominant, AR: Autosomal recessive. Phenotypes according to OMIM https://www.omim.org/; NA: Not available.

**Table S8. Colocalization results between GTEx expression data and significant loci with PP H4>80%.**

| Biomarker | Gene | Tissue | PP(H4)% |
|---|---|---|---|
| AREG (T7) | *lnc-NUAK1-1* | Adipose Subcutaneous | 92.37 |
| | *CASC18* | Adipose Subcutaneous | 91.62 |
| | *lnc-NUAK1-1* | Artery Aorta | 92.35 |
| | *lnc-NUAK1-1* | Artery Tibial | 92.37 |
| | *lnc-NUAK1-1* | Brain Cerebellar Hemisphere | 90.99 |
| | *lnc-TCP11L2-1* | Brain Cerebellum | 86.69 |
| | *lnc-NUAK1-1* | Brain Cerebellum | 82.09 |
| | *lnc-NUAK1-1* | Brain Cortex | 92.50 |
| | *lnc-NUAK1-1* | Breast Mammary Tissue | 92.37 |
| | *lnc-NUAK1-1* | Nerve Tibial | 92.37 |
| | *lnc-NUAK1-1* | Prostate | 92.35 |
| | *lnc-NUAK1-1* | Skin Not Sun Exposed Suprapubic | 92.37 |
| | *lnc-NUAK1-1* | Skin Sun Exposed Lower leg | 92.37 |
| | *lnc-NUAK1-1* | Uterus | 86.06 |
| ICAM-1 (T2) | *MROH2A* | Esophagus Mucosa | 88.30 |
| | | Skin Not Sun Exposed Suprapubic | 93.10 |
| | | Skin Sun Exposed Lower leg | 89.29 |
| SP-D (T1) | *RB1CC1* | Esophagus Gastroesophageal Junction | 85.41 |
| | *ALKAL1* | Stomach | 85.34 |
| SP-D (T7) | *NPBWR1* | Liver | 87.38 |

PP: posterior probability. T1: within 24 hours after sepsis diagnosis; T2: 48-72 hours after sepsis diagnosis; T7: 7 days after sepsis diagnosis.

**Table S9. Association results of rare variants in the prioritized genes with sepsis-associated ARDS.**

| Gene | Position | GDI (v2) | P-value | | |
|---|---|---|---|---|---|
| | | | AF<0.001 | AF<0.001& biological impact | AF<0.001 & synonyms |
| *C1GALT1* | chr7:7196565-7288282 | 0.32 | 0.468 | 0.673 | NA |
| *CCZ1B* | chr7:6833765-6866401 | 2.39 | 0.626 | 0.509 | NA |
| *CDH20* | chr18:59000815-5922306 | 1.12 | 0.437 | 0.680 | 0.053 |
| *CSTB* | chr21:45192393-45196326 | 2.64 | 0.431 | 0.684 | NA |
| *DENND2C* | chr1:115125469-115213043 | 0.47 | 0.833 | 0.441 | NA |
| *ECT2* | chr3:172468472-172539264 | 0.92 | 0.181 | 0.490 | 0.883 |
| *FAM134B* | chr5:16473147-16617167 | 1.49 | 0.694 | NA | 0.689 |
| *FAM81B* | chr5:94727048-94786158 | 10.70 | 0.853 | 0.849 | 0.746 |
| *FOXN1* | chr17:26833261-26865914 | 2.03 | 0.255 | 0.112 | 0.178 |
| *GTF2A1* | chr14:81641796-81687721 | 0.30 | 0.205 | 0.571 | NA |
| *HDAC4* | chr2:239969864-240323348 | 0.10 | 0.304 | 0.216 | 0.827 |
| *ITGA1* | chr5:52083730-52255040 | 0.07 | 0.864 | 0.865 | 0.225 |
| *LINGO2* | chr9:27948076-28670283 | 1.59 | 0.556 | 0.790 | 0.511 |
| *LURAP1* | chr1:46669006-46686933 | 3.41 | 0.642 | 0.473 | 0.920 |
| *MC4R* | chr18:58038564-58040001 | 4.37 | 0.714 | 0.322 | 0.140 |
| *MCTP1* | chr5:94039446-94620279 | 0.53 | 0.695 | 0.254 | 0.180 |
| *MROH2A* | chr2:234684370-234742069 | 19.60 | 0.173 | 0.314 | 0.243 |
| *NCEH1* | chr3:172348039-172429008 | 1.15 | 0.253 | $5.00\times10^{-3}$ | 0.708 |
| *NDFIP2* | chr13:80055287-80130210 | 0.79 | 0.330 | 0.230 | 0.752 |
| *NUAK1* | chr12:106457118-106533811 | 0.30 | 0.500 | 0.730 | 0.156 |
| *PDXK* | chr21:45138975-45182188 | 0.19 | 0.334 | 0.185 | 0.113 |
| *PIEZO2* | chr18:10666480-11148587 | 0.43 | 0.616 | 0.259 | 0.099 |
| *PIK3R3* | chr1:46505812-46642160 | 0.98 | 0.975 | 0.211 | 0.733 |
| *PMP22* | chr17:15133095-15168643 | 3.32 | 0.606 | 0.175 | 0.426 |
| *PTPRD* | chr9:8314246-10612723 | 0.08 | $3.11\times10^{-4}$ | 0.025 | 0.022 |
| *RB1CC1* | chr8:53535016-53658403 | 0.47 | 0.480 | 0.770 | 0.479 |
| *SCARB1* | chr12:125261402-125367214 | 2.31 | 0.468 | 0.124 | 0.570 |
| *SGK494* | chr17:26934982-26941218 | NA | 0.202 | 0.956 | 0.704 |
| *SLC13A2* | chr17:26800311-26824799 | 3.63 | 0.123 | 0.237 | 0.360 |
| *SOX1* | chr13:112721913-112726020 | 1.19 | 0.761 | 0.761 | 0.761 |
| *SPACA7* | chr13:113030633-113089003 | 11.50 | 0.922 | 0.555 | 0.604 |
| *SPATA13* | chr13:24553944-24881212 | 0.14 | 0.475 | 0.334 | 0.734 |

| (Cont. Table S9) | | | P-value | | |
|---|---|---|---|---|---|
| Gene | Position | GDI (v2) | AF<0.001 | AF<0.001& biological impact | AF<0.001 & synonyms |
| *SYT6* | chr1:114631913-114696541 | 0.96 | 0.975 | 0.648 | 0.624 |
| *TEKT3* | chr17:15207128-15244958 | 10.45 | 0.604 | 0.170 | 0.871 |
| *TMEM132B* | chr12:125671382-126146917 | 0.06 | 0.991 | 0.202 | 0.749 |
| *TMEM65* | chr8:125324231-125384933 | 1.42 | 0.558 | 0.360 | 0.748 |
| *TRAM1L1* | chr4:118004718-118006736 | 2.90 | 0.428 | 0.428 | NA |
| *TRAPPC10* | chr21:45432200-45526433 | 0.37 | 0.692 | 0.645 | 0.296 |
| *TRIM33* | chr1:114935399-115053781 | 0.32 | 0.969 | 0.508 | 0.416 |
| *TRMT12* | chr8:125463048-125474391 | 6.22 | 0.714 | 0.714 | NA |
| *TVP23C-CDRT4* | chr17:15339338-15466875 | NA | 0.352 | 0.352 | NA |

AF: Allele frequency; GDI v2: Gene Damage Index version 2, position: chromosome, start and end according to Ensembl (GRCh37/hg19); NA: Not available. GDI from: https://hgidsoft.rockefeller.edu/GDI/GDIv2.html

**Table S10. Annotation of variants in *PTPRD* included in the rare variant analysis.**

| Variant ID | rsID | AF gnomAD | CADD | Exon | HGVSp | Codons |
|---|---|---|---|---|---|---|
| chr9:8319735:T:C | rs562333762 | NA | 4.91 | . | . | . |
| chr9:8319841:A:T* | rs1200247931 | $2.65x10^{-5}$ | 31 | 30/31 | p.Val1481Glu | gTa/gAa |
| chr9:8319965:C:T* | rs141403124 | $2.00x10^{-4}$ | 33 | 30/31 | p.Ala1440Thr | Gcg/Acg |
| chr9:8319966:G:A | rs748716978 | $3.55x10^{-5}$ | 10.28 | 30/31 | p.Ser1439= | agC/agT |
| chr9:8331520:G:A | rs563708117 | NA | 0.59 | . | . | . |
| chr9:8331644:G:A | rs140620944 | $2.50x10^{-3}$ | 5.31 | 29/31 | p.Ile1418= | atC/atT |
| chr9:8338890:A:G | rs1446833918 | $9.21x10^{-6}$ | 2.80 | . | . | . |
| chr9:8339091:G:A | rs1378114841 | $1.02x10^{-5}$ | 0.61 | . | . | . |
| chr9:8340301:G:C | rs148055998 | $1.00x10^{-4}$ | 1.21 | . | . | . |
| chr9:8340316:G:T | rs1392514 | $3.00x10^{-4}$ | 1.22 | . | . | . |
| chr9:8341185:A:G | rs12351899 | $7.06x10^{-5}$ | 9.01 | 26/31 | p.Asn1271= | aaT/aaC |
| chr9:8341272:T:C* | rs199799750 | $5.00x10^{-4}$ | 23.1 | . | . | . |
| chr9:8341930:T:C | rs146073475 | 0 | 1.52 | 25/31 | p.Leu1164= | ttA/ttG |
| chr9:8376501:T:C | rs188044460 | NA | 4.99 | . | . | . |
| chr9:8376681:C:T* | rs751203177 | $1.76x10^{-5}$ | 32 | 23/31 | p.Gly1072Arg | Gga/Aga |
| chr9:8376685:G:C | rs745835445 | $8.81x10^{-6}$ | 8.43 | 23/31 | p.Thr1070= | acC/acG |
| chr9:8376686:G:T* | rs756058132 | $7.05x10^{-5}$ | 24.5 | 23/31 | p.Thr1070Asn | aCc/aAc |
| chr9:8389213:G:C | rs373568470 | $2.00x10^{-4}$ | 1.40 | . | . | . |
| chr9:8389343:A:G | rs75966719 | $2.00x10^{-4}$ | 7.41 | 22/31 | p.Tyr1019= | taT/taC |
| chr9:8404547:T:A | rs550824027 | $3.53x10^{-5}$ | 9.81 | 21/31 | p.Ser994= | tcA/tcT |
| chr9:8404589:G:A | rs771214124 | $8.83x10^{-6}$ | 6.29 | 21/31 | p.Tyr980= | taC/taT |
| chr9:8436658:C:G* | rs773244173 | $8.84x10^{-6}$ | 22.7 | 20/31 | p.Leu934Phe | ttG/ttC |
| chr9:8437141:G:C* | rs10977104 | NA | 18.67 | . | . | . |
| chr9:8437199:T:G* | rs190764618 | $2.00x10^{-4}$ | 22.4 | 17/29 | p.Ser922= | tcA/tcC |
| chr9:8449734:G:C* | rs773381158 | $8.81x10^{-6}$ | 24.1 | 19/31 | p.Gln921Glu | Caa/Gaa |
| chr9:8449870:A:C | rs140068185 | 0 | 9.33 | . | . | . |
| chr9:8451816:T:A | rs4742505 | NA | 1.90 | . | . | . |
| chr9:8451817:T:C | rs4742506 | NA | 6.18 | . | . | . |
| chr9:8454495:G:A | rs73640916 | NA | 11.39 | . | . | . |
| chr9:8454532:T:C | rs756332284 | $9.11x10^{-6}$ | 4.33 | . | . | . |
| chr9:8454609:A:C* | rs762716454 | $6.24x10^{-5}$ | 22.5 | . | . | . |
| chr9:8454676:C:T* | rs182563984 | NA | 21 | . | . | . |
| chr9:8460220:G:A | rs187702894 | NA | 1.86 | . | . | . |
| chr9:8460321:A:G | rs377744836 | $1.00x10^{-4}$ | 11.31 | . | . | . |
| chr9:8460369:G:T | rs780834539 | 0 | 0.69 | . | . | . |
| chr9:8460504:G:A* | rs148300682 | $4.42x10^{-5}$ | 23.2 | 17/31 | p.Thr850Met | aCg/aTg |
| chr9:8465634:A:G | rs570130168 | $1.00x10^{-4}$ | 7.43 | 16/31 | p.Tyr771= | taT/taC |
| chr9:8465663:T:G* | rs561964160 | $9.77x10^{-5}$ | 22.9 | 16/31 | p.Ile762Leu | Ata/Cta |
| chr9:8470914:G:A | rs142050629 | NA | 1.26 | . | . | . |
| chr9:8470937:C:T | rs557861197 | NA | 0.03 | . | . | . |
| chr9:8470960:C:T | rs200356129 | $4.00x10^{-4}$ | 4.25 | . | . | . |
| chr9:8471046:G:A | rs7865681 | $1.00x10^{-4}$ | 7.55 | 15/31 | p.Arg740= | cgC/cgT |
| chr9:8484190:G:A | rs61733195 | $7.03x10^{-5}$ | 11.97 | 14/31 | p.Phe703= | ttC/ttT |
| chr9:8484240:T:G | rs12344148 | $4.00x10^{-4}$ | 10.45 | 14/31 | p.Arg687= | Agg/Cgg |
| chr9:8484298:C:A* | rs7869444 | $2.00x10^{-4}$ | 19.36 | 14/31 | p.Glu667Asp | gaG/gaT |
| chr9:8484315:C:T* | rs140371409 | 0 | 23.3 | 14/31 | p.Val662Ile | Gtc/Atc |
| chr9:8485177:G:T | rs117337760 | $3.00x10^{-4}$ | 2.30 | . | . | . |

**(Cont. Table S10)**

| Variant ID | rsID | AF gnomAD | CADD | Exon | HGVSp | Codons |
|---|---|---|---|---|---|---|
| chr9:8485193:T:C | rs76971582 | $3.57x10^{-5}$ | 6.85 | . | . | . |
| chr9:8485787:C:G* | rs143674254 | $1.78x10^{-5}$ | 23.4 | 17/35 | p.Gln1010His | caG/caC |
| chr9:8485882:T:C | rs762428743 | $3.52x10^{-5}$ | 13.43 | 17/35 | p.Met979Val | Atg/Gtg |
| chr9:8485885:T:C* | rs139259906 | $3.00x10^{-4}$ | 19.56 | 17/35 | p.Thr978Ala | Act/Gct |
| chr9:8485903:T:C* | rs200885047 | $4.00x10^{-4}$ | 17.47 | 17/35 | p.Ile972Val | Att/Gtt |
| chr9:8485917:G:A* | rs757868633 | $1.76x10^{-5}$ | 23.7 | 17/35 | p.Pro967Leu | cCg/cTg |
| chr9:8485928:G:A | rs3824417 | $1.00x10^{-4}$ | 8.98 | 17/35 | p.Ile963= | atC/atT |
| chr9:8485996:G:C* | rs61733170 | $4.40x10^{-5}$ | 22.6 | 17/35 | p.Pro941Ala | Cca/Gca |
| chr9:8486080:T:C* | rs374125865 | $1.00x10^{-4}$ | 15.23 | 17/35 | p.Ile913Val | Att/Gtt |
| chr9:8486182:G:A* | rs201583436 | $2.64x10^{-5}$ | 23.3 | 17/35 | p.His879Tyr | Cac/Tac |
| chr9:8486201:C:T | rs1246819365 | 0 | 6.47 | 17/35 | p.Glu872= | gaG/gaA |
| chr9:8486335:G:A* | rs141603921 | $8.92x10^{-5}$ | 23.4 | 17/35 | p.Arg828Trp | Cgg/Tgg |
| chr9:8486409:T:C | rs185917936 | $9.00x10^{-4}$ | 0.35 | . | . | . |
| chr9:8486440:A:G | rs370464763 | $9.34x10^{-6}$ | 5.07 | . | . | . |
| chr9:8486547:A:G | rs28491557 | NA | 7.52 | . | . | . |
| chr9:8492804:T:A | rs201064171 | NA | 0.26 | . | . | . |
| chr9:8492898:G:A* | rs377557345 | $4.40x10^{-5}$ | 32 | 16/35 | p.Arg811Cys | Cgc/Tgc |
| chr9:8492902:A:T | rs12346849 | $6.00x10^{-4}$ | 9.70 | 16/35 | p.Gly809= | ggT/ggA |
| chr9:8492977:G:A | rs142228841 | $4.00x10^{-4}$ | 12.52 | 16/35 | p.Asp784= | gaC/gaT |
| chr9:8497287:G:A* | rs375498800 | $4.00x10^{-4}$ | 16.34 | . | . | . |
| chr9:8499610:T:G | rs761743935 | $3.82x10^{-5}$ | 0.45 | . | . | . |
| chr9:8499743:G:T | rs757978113 | $8.82x10^{-6}$ | 10.27 | 14/35 | p.Gly742= | ggC/ggA |
| chr9:8499805:C:A* | rs150444130 | $1.50x10^{-3}$ | 22.6 | 14/35 | p.Ala722Ser | Gct/Tct |
| chr9:8499856:T:C* | rs199575422 | $8.00x10^{-4}$ | 16.53 | . | . | . |
| chr9:8500736:G:A* | rs774806301 | $8.93x10^{-6}$ | 15.93 | . | . | . |
| chr9:8500896:A:G | rs760953800 | $1.76x10^{-5}$ | 11.67 | 13/35 | p.Ile662= | atT/atC |
| chr9:8504361:T:C | rs139810753 | $1.10x10^{-3}$ | 11.43 | 12/31 | p.Gly574= | ggA/ggG |
| chr9:8507260:G:A | rs202009280 | $3.70x10^{-5}$ | 2.70 | . | . | . |
| chr9:8518056:A:G | rs200537516 | $2.00x10^{-4}$ | 1.51 | 10/31 | p.Asn445= | aaT/aaC |
| chr9:8518098:C:A | rs145325302 | $8.00x10^{-4}$ | 0.004 | 10/31 | p.Ser431= | tcG/tcT |
| chr9:8521482:G:A | rs765489687 | $8.84x10^{-6}$ | 9.43 | 9/31 | p.Ser252= | agC/agT |
| chr9:8523537:G:A* | rs776467229 | $5.32x10^{-5}$ | 21.9 | . | . | . |
| chr9:8524854:C:T | rs2273998 | NA | 1.76 | . | . | . |
| chr9:8524862:G:A | rs72694784 | NA | 2.17 | . | . | . |
| chr9:8524896:A:G | rs1357467837 | 0 | 10.22 | . | . | . |
| chr9:8524950:G:A | rs201210585 | $9.711x10^{-5}$ | 9.65 | 7/31 | p.Ser218= | tcC/tcT |
| chr9:8525110:A:C* | rs753241424 | $5.38x10^{-5}$ | 19.62 | . | . | . |
| chr9:8525139:G:T | rs200358210 | $9.97x10^{-5}$ | 13.72 | . | . | . |
| chr9:8525192:G:A* | rs1032045827 | 0 | 20.2 | . | . | . |
| chr9:8526634:T:C | rs201761674 | $4.00x10^{-4}$ | 12.77 | 6/31 | p.Pro187= | ccA/ccG |
| chr9:8527281:A:C* | rs547799522 | NA | 19.23 | . | . | . |
| chr9:8528661:G:A | rs149990369 | 0 | 11.46 | 4/31 | p.Ile157= | atC/atT |
| chr9:8528767:G:A* | rs752353348 | 0 | 29.3 | 4/31 | p.Pro122Leu | cCc/cTc |
| chr9:8528828:C:T | rs373726631 | $2.00x10^{-4}$ | 3.50 | . | . | . |
| chr9:8633287:A:G | rs372661922 | $4.00x10^{-4}$ | 8.83 | . | . | . |
| chr9:8633425:G:A* | rs776040231 | $8.82x10^{-6}$ | 26.8 | 3/31 | p.Leu82Phe | Ctc/Ttc |
| chr9:8633450:C:T | rs74945428 | 0 | 11.54 | 3/31 | p.Glu73= | gaG/gaA |
| chr9:8633542:G:A | rs187327262 | NA | 4.23 | . | . | . |

**(Cont. Table S10)**

| Variant ID | rsID | AF gnomAD | CADD | Exon | HGVSp | Codons |
|---|---|---|---|---|---|---|
| chr9:8636765:C:T | rs146589803 | 0 | 6.16 | 2/31 | p.Thr48= | acG/acA |
| chr9:8636792:T:A | rs138451374 | $1.00x10^{-4}$ | 11.45 | 2/31 | p.Gly39= | ggA/ggT |
| chr9:8733701:G:C* | rs556216909 | NA | 15.56 | . | . | . |
| chr9:8733788:T:A* | rs199621346 | $3.00x10^{-4}$ | 18.92 | 1/31 | p.Asp19Val | gAt/gTt |
| chr9:8733869:G:A | rs192265997 | $6.00x10^{-4}$ | 11.94 | 1/31 | . | . |

*Variant with high biological impact; AF: allele frequency; AF gnomAD: AF in non-Finish Europeans; CADD: Combined Annotation Dependent Depletion; Variant ID: chromosome, base pair, reference and alternative allele based on GRCh37/hg19; NA: Not available.

**Table S11. Annotation of variants in *NCEH1* included in the rare variant analysis.**

| Variant ID | rsID | AF gnomAD | CADD | Exon | HGVSp | Codons |
|---|---|---|---|---|---|---|
| chr3:172353692:G:A | rs768432542 | $1.76 \times 10^{-5}$ | 6.12 | . | . | . |
| chr3:172363391:A:T* | rs754149316 | $9.13 \times 10^{-6}$ | 11.59 | . | . | . |
| chr3:172365715:C:T | rs377294886 | $8.80 \times 10^{-6}$ | 7.26 | 2/5 | p.Val142Ile | Gtt/Att |
| chr3:172365821:C:T | rs147779888 | $3.00 \times 10^{-4}$ | 0.09 | 2/5 | p.Ala106= | gcG/gcA |
| chr3:172365939:G:A | rs754699748\| | $4.47 \times 10^{-5}$ | 0.15 | . | . | . |
| chr3:172428745:G:T | rs376815925 | $3.52 \times 10^{-5}$ | 9.66 | 1/5 | p.Ala42= | gcC/gcA |

*Variant with high biological impact; AF: allele frequency; AF gnomAD: AF in non-Finish Europeans; CADD: Combined Annotation Dependent Depletion; Variant ID: chromosome, base pair, reference and alternative allele based on GRCh37/hg19.

**Table S12. Results of the best PGS prediction models for Intensive Care Unit mortality for each biomarker.**

| GWAS | Time | Threshold | OR[95%CI] | *p*-value | #SNPs |
|---|---|---|---|---|---|
| Ang-2 | T1 | 0.027 | 0.87[0.71-1.07] | 0.181 | 25,141 |
| | T2 | $3.83 \times 10^{-4}$ | 1.34[1.11-1.62] | $2.28 \times 10^{-3}$ | 697 |
| | T7 | 1 | 0.81[0.67-0.98] | 0.026 | 313,430 |
| AREG | T1 | $1.02 \times 10^{-3}$ | 0.80[0.67-0.96] | 0.016 | 1,140 |
| | T2 | 0.230 | 0.76[0.63-0.91] | $3.47 \times 10^{-3}$ | 116,565 |
| | T7 | $7.60 \times 10^{-4}$ | 0.80[0.67-0.97] | 0.022 | 739 |
| CXCL12 | T1 | 0.127 | 1.24[1.03-1.49] | 0.025 | 87,695 |
| | T2 | $6.10 \times 10^{-4}$ | 0.79[0.66-0.95] | 0.014 | 708 |
| | T7 | $9.63 \times 10^{-5}$ | 1.33[1.10-1.59] | $2.84 \times 10^{-3}$ | 117 |
| CXCL16 | T1 | $1.66 \times 10^{-3}$ | 0.81[0.68-0.98] | 0.028 | 2,245 |
| | T2 | $3.51 \times 10^{-3}$ | 1.21[1.01-1.47] | 0.043 | 3,789 |
| | T7 | $1.41 \times 10^{-4}$ | 0.78[0.65-0.94] | $8.66 \times 10^{-3}$ | 157 |
| ICAM-1 | T1 | $1.40 \times 10^{-6}$ | 1.16[0.97-1.39] | 0.112 | 3 |
| | T2 | $4.11 \times 10^{-3}$ | 0.79[0.66-0.95] | 0.014 | 4,554 |
| | T7 | 0.014 | 0.75[0.62-0.91] | $3.03 \times 10^{-3}$ | 13,319 |
| IL-1RA | T1 | $8.70 \times 10^{-6}$ | 0.81[0.67-0.97] | 0.024 | 5 |
| | T2 | $5.40 \times 10^{-6}$ | 0.77[0.64-0.93] | $6.42 \times 10^{-3}$ | 3 |
| | T7 | $9.38 \times 10^{-3}$ | 1.30[1.08-1.57] | $5.41 \times 10^{-3}$ | 5,747 |
| IL-18 | T1 | $5.80 \times 10^{-6}$ | 1.25[1.04-1.50] | 0.015 | 12 |
| | T2 | 0.041 | 0.85[0.71-1.03] | 0.097 | 34,381 |
| | T7 | $1.49 \times 10^{-4}$ | 1.28[1.06-1.55] | 0.010 | 242 |
| PAI-1 | T1 | $9.79 \times 10^{-5}$ | 1.27[1.06-1.52] | 0.011 | 210 |
| | T2 | $4.04 \times 10^{-3}$ | 0.84[0.70-1.01] | 0.062 | 5,856 |
| | T7 | $3.32 \times 10^{-3}$ | 0.71[0.59-0.86] | $3.26 \times 10^{-4}$ | 3,896 |
| RAGE | T1 | $2.03 \times 10^{-5}$ | 0.79[0.66-0.96] | 0.015 | 29 |
| | T2 | $1.34 \times 10^{-3}$ | 0.81[0.67-0.97] | 0.023 | 2,067 |
| | T7 | 0.045 | 0.86[0.72-1.04] | 0.125 | 36,089 |
| SP-D | T1 | $5.15 \times 10^{-5}$ | 1.20[1.00-1.44] | 0.046 | 110 |
| | T2 | $2.65 \times 10^{-4}$ | 0.82[0.68-0.99] | 0.037 | 404 |
| | T7 | $1.75 \times 10^{-6}$ | 0.85[0.71-1.03] | 0.090 | 3 |

#SNPs: number of SNPs included in the model. CI: confident interval; OR: odds ratio. T1: within 24 hours after sepsis diagnosis; T2: 48-72 hours after sepsis diagnosis; T7: 7 days after sepsis diagnosis.

**Table S13. Results of the best PGS prediction models for sepsis-associated ARDS for each biomarker.**

| GWAS | Time | Threshold | OR[95%CI] | *p*-value | #SNPs |
|---|---|---|---|---|---|
| Ang-2 | T1 | $3.55x10^{-6}$ | 1.20 [1.01-1.42] | 0.041 | 13 |
| | T2 | $8.39x10^{-4}$ | 1.20 [1.01-1.42] | 0.035 | 1,406 |
| | T7 | $2.34x10^{-4}$ | 0.87 [0.74-1.03] | 0.116 | 295 |
| AREG | T1 | $2.30x10^{-4}$ | 1.18 [0.99-1.39] | 0.057 | 260 |
| | T2 | 0.424 | 1.17 [0.99-1.39] | 0.067 | 178,470 |
| | T7 | $2.90x10^{-6}$ | 0.83 [0.70-0.98] | 0.032 | 7 |
| CXCL12 | T1 | $1.55x10^{-4}$ | 0.79 [0.67-0.94] | $6.98x10^{-3}$ | 204 |
| | T2 | $1.90x10^{-6}$ | 1.24 [1.05-1.47] | 0.012 | 2 |
| | T7 | $2.75x10^{-6}$ | 1.15 [0.96-1.37] | 0.121 | 2 |
| CXCL16 | T1 | $1.92x10^{-5}$ | 0.76 [0.64-0.91] | $2.25x10^{-3}$ | 40 |
| | T2 | $6.13x10^{-3}$ | 1.18 [0.99-1.40] | 0.060 | 6,433 |
| | T7 | $1.32x10^{-5}$ | 1.12 [0.95-1.33] | 0.174 | 15 |
| ICAM-1 | T1 | 0.048 | 1.20 [1.01-1.44] | 0.040 | 40,518 |
| | T2 | $1.02x10^{-4}$ | 1.22 [1.03-1.45] | 0.023 | 124 |
| | T7 | $2.13x10^{-4}$ | 1.27 [1.07-1.50] | $6.82x10^{-3}$ | 286 |
| IL-1RA | T1 | $2.78x10^{-3}$ | 1.19 [1.00-1.41] | 0.048 | 2,706 |
| | T2 | $5.40x10^{-6}$ | 0.84 [0.70-0.99] | 0.040 | 3 |
| | T7 | $1.00x10^{-6}$ | 0.79 [0.67-0.94] | $7.78x10^{-3}$ | 2 |
| IL-18 | T1 | 0.0278 | 0.83 [0.69-0.99] | 0.035 | 26,032 |
| | T2 | $3.21x10^{-4}$ | 1.20 [1.01-1.42] | 0.033 | 480 |
| | T7 | $5.20x10^{-6}$ | 0.82 [0.69-0.97] | 0.022 | 14 |
| PAI-1 | T1 | $2.81x10^{-5}$ | 0.79 [0.67-0.94] | $7.81x10^{-3}$ | 63 |
| | T2 | $1.76x10^{-3}$ | 0.87 [0.73-1.03] | 0.101 | 2,990 |
| | T7 | $6.20x10^{-6}$ | 0.78 [0.66-0.93] | $4.54x10^{-3}$ | 8 |
| RAGE | T1 | 0.0475 | 1.19 [1.00-1.41] | 0.050 | 40,149 |
| | T2 | $1.84x10^{-5}$ | 1.22 [1.02-1.44] | 0.026 | 42 |
| | T7 | $1.19x10^{-4}$ | 0.82 [0.69-0.97] | 0.023 | 219 |
| SP-D | T1 | 0.497 | 1.29 [1.08-1.53] | $4.53x10^{-3}$ | 228,104 |
| | T2 | $3.01x10^{-7}$ | 0.83 [0.70-0.99] | 0.036 | 1 |
| | T7 | 0.0352 | 1.16 [0.98-1.38] | 0.080 | 29,228 |

#SNPs: number of SNPs included in the model. CI: confident interval; OR: odds ratio; T1: within 24 hours after sepsis diagnosis; T2: 48-72 hours after sepsis diagnosis; T7: 7 days after sepsis diagnosis.

**Table S14. Results of the best PGS prediction models for multiple organ failure for each biomarker.**

| GWAS | Time | Threshold | OR[95%CI] | *p*-value | #SNPs |
|---|---|---|---|---|---|
| Ang-2 | T1 | $1.15x10^{-6}$ | 0.87[0.70-1.07] | 0.189 | 8 |
| | T2 | $2.35x10^{-4}$ | 1.28[1.04-1.58] | 0.021 | 435 |
| | T7 | $1.72x10^{-3}$ | 1.43[1.16-1.78] | $9.61x10^{-4}$ | 1,932 |
| AREG | T1 | $1.55x10^{-4}$ | 0.84[0.69-1.03] | 0.096 | 168 |
| | T2 | $4.28x10^{-3}$ | 0.79[0.65-0.98] | 0.029 | 3,792 |
| | T7 | 0.054 | 0.84[0.68-1.04] | 0.105 | 34,365 |
| CXCL12 | T1 | $9.76x10^{-5}$ | 0.82[0.67-1.01] | 0.062 | 121 |
| | T2 | $6.96x10^{-5}$ | 1.16[0.94-1.43] | 0.171 | 80 |
| | T7 | $3.16x10^{-5}$ | 1.28[1.04-1.59] | 0.023 | 36 |
| CXCL16 | T1 | $3.76x10^{-5}$ | 0.77[0.63-0.95] | 0.013 | 81 |
| | T2 | $5.94x10^{-3}$ | 1.33[1.09-1.64 | $6.03x10^{-3}$ | 6,264 |
| | T7 | $6.34x10^{-2}$ | 1.17[0.95-1.43] | 0.137 | 46,919 |
| ICAM-1 | T1 | $1.16x10^{-3}$ | 0.86[0.70-1.06] | 0.149 | 1,603 |
| | T2 | 0.016 | 0.78[0.63-0.96] | 0.019 | 15,037 |
| | T7 | $1.17x10^{-5}$ | 1.20[0.98-1.47] | 0.078 | 22 |
| IL-1RA | T1 | $1.67x10^{-5}$ | 0.79[0.65-0.97] | 0.026 | 16 |
| | T2 | $9.51x10^{-7}$ | 0.75[0.61-0.93] | 0.010 | 2 |
| | T7 | 0.017 | 0.75[0.61-0.92] | $5.28x10^{-3}$ | 9,130 |
| IL-18 | T1 | $5.85x10^{-6}$ | 1.43[1.16-1.77] | $9.88x10^{-4}$ | 13 |
| | T2 | $7.86x10^{-4}$ | 0.86[0.70-1.05] | 0.146 | 1,109 |
| | T7 | $1.17x10^{-3}$ | 1.24[1.00-1.53] | 0.046 | 1,411 |
| PAI-1 | T1 | $1.30x10^{-3}$ | 0.83[0.68-1.02] | 0.071 | 2,018 |
| | T2 | $1.51x10^{-7}$ | 1.44[1.15-1.81] | $1.80x10^{-3}$ | 2 |
| | T7 | $1.76x10^{-5}$ | 0.80[0.65-0.99] | 0.039 | 19 |
| RAGE | T1 | $1.55x10^{-4}$ | 1.22[1.00-1.50] | 0.051 | 260 |
| | T2 | $9.42x10^{-3}$ | 0.80[0.65-0.98] | 0.030 | 10,520 |
| | T7 | $1.27x10^{-4}$ | 0.73[0.59-0.90] | $3.62x10^{-3}$ | 234 |
| SP-D | T1 | $1.85x10^{-6}$ | 1.25[1.01-1.55] | 0.039 | 6 |
| | T2 | $2.78x10^{-5}$ | 0.76[0.61-0.93] | $7.83x10^{-3}$ | 48 |
| | T7 | 0.013 | 0.81[0.66-1.00] | 0.050 | 12,496 |

#SNPs: number of SNPs included in the model. CI: confident interval; OR: odds ratio; T1: within 24 hours after sepsis diagnosis; T2: 48-72 hours after sepsis diagnosis; T7: 7 days after sepsis diagnosis.

**Table S15. Most significant results of pathway-based polygenic scores for PAI-1 levels at T7 and mortality.**

| Pathway | Coefficient | SE | P-value | #SNPs |
|---|---|---|---|---|
| RNA Polymerase I Promoter Opening | -0.397 | 0.097 | $3.94 \times 10^{-5}$ | 23 |
| Reversal of alkylation damage by DNA dioxygenases | -0.310 | 0.096 | $1.21 \times 10^{-3}$ | 177 |
| Keratinization | -0.301 | 0.095 | $1.50 \times 10^{-3}$ | 814 |
| Negative epigenetic regulation of rRNA expression | -0.295 | 0.096 | $2.14 \times 10^{-3}$ | 414 |
| NoRC negatively regulates rRNA expression | -0.291 | 0.096 | $2.44 \times 10^{-3}$ | 394 |
| Membrane Trafficking | -0.285 | 0.096 | $2.83 \times 10^{-3}$ | 7,601 |
| Formation of the cornified envelope | -0.279 | 0.094 | $3.06 \times 10^{-3}$ | 757 |
| Vesicle-mediated transport | -0.280 | 0.095 | $3.30 \times 10^{-3}$ | 8 ,237 |
| Defective CYP26C1 causes FFDD4 | -0.295 | 0.102 | $3.93 \times 10^{-3}$ | 5 |
| Astrocytic Glutamate-Glutamine Uptake and Metabolism | -0.278 | 0.096 | $3.99 \times 10^{-3}$ | 79 |
| Neurotransmitter uptake and metabolism in glial cells | -0.278 | 0.096 | $3.99 \times 10^{-3}$ | 79 |

#SNPs: number of SNPs included in the model. SE: standard error

### Supplementary Figures

**Figure S1. Box plot of normalized serum levels (pg/ml) at T1, T2 and, T7 of Ang-2; AREG, CXCL12, CXCL16, ICAM-I, IL-1RA, IL-18, PAI-1, RAGE, and SP-D.** The *p*-values show the results derived from the t-test. T1: levels obtained within 24 hours after sepsis diagnosis; T2: levels obtained within 48-72 hours after sepsis diagnosis; T7: levels obtained 7 days after sepsis diagnosis.

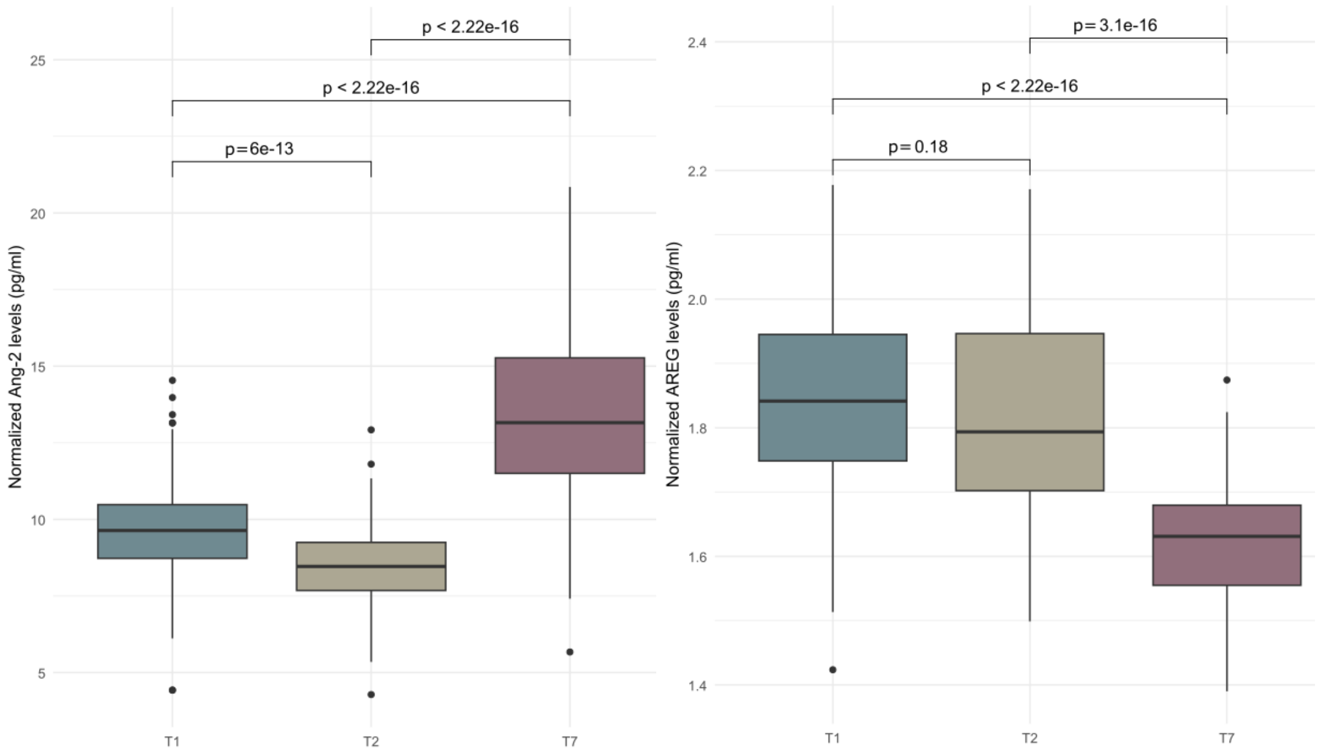

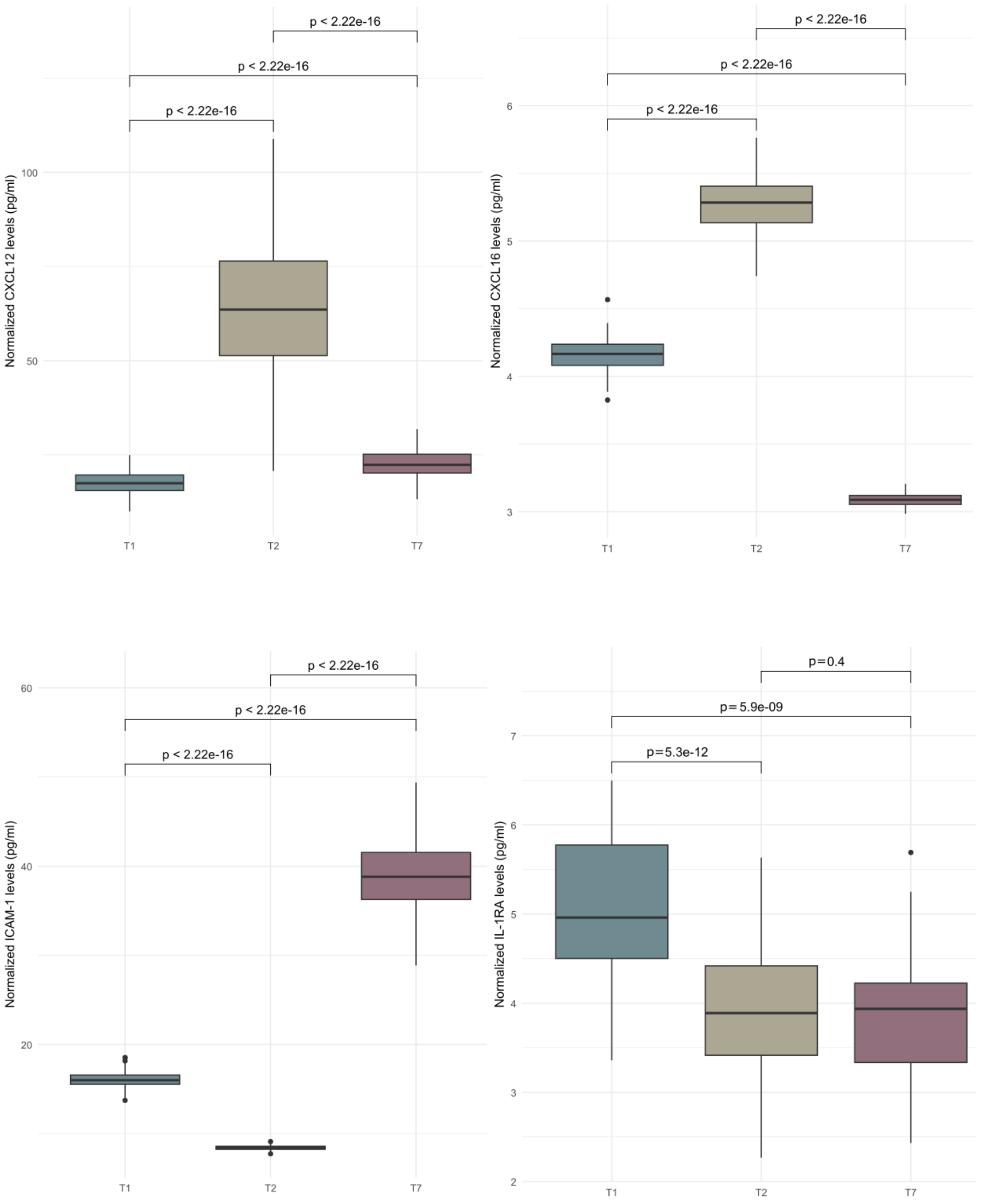
Normalized CXCL12 levels (pg/ml)
p < 2.22e-16
p < 2.22e-16
p < 2.22e-16
100
50
T1
T2
T7
Normalized CXCL16 levels (pg/ml)
p < 2.22e-16
p < 2.22e-16
p < 2.22e-16
6
5
4
3
T1
T2
T7
Normalized ICAM-1 levels (pg/ml)
p < 2.22e-16
p < 2.22e-16
p < 2.22e-16
60
40
20
T1
T2
T7
Normalized IL-1RA levels (pg/ml)
p=0.4
p= 5.9e-09
p=5.3e-12
7
6
5
4
3
2
T1
T2
T7

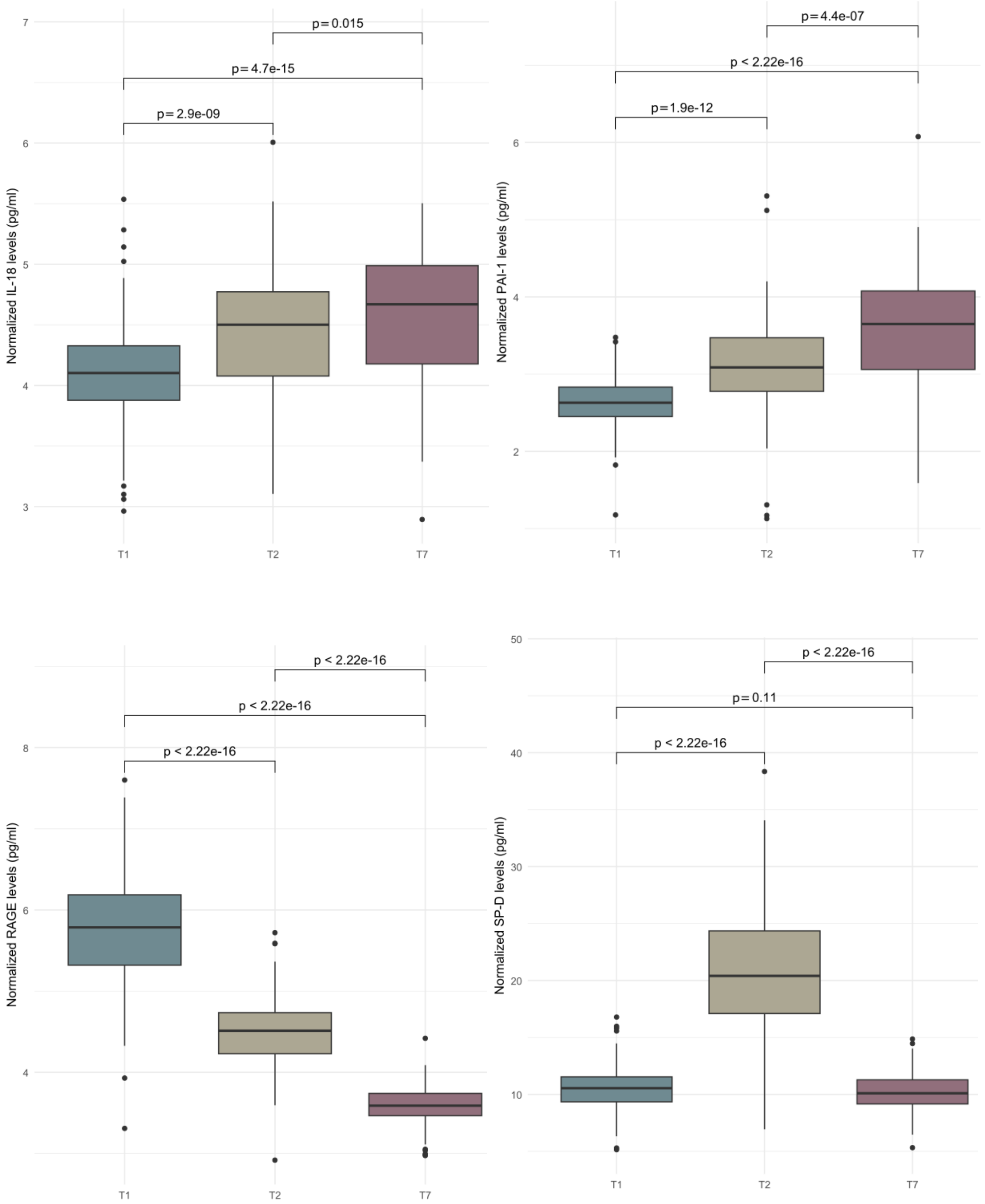
Normalized IL-18 levels (pg/ml)
p = 0.015
p = 4.7e-15
p = 2.9e-09
7
6
5
4
3
T1
T2
T7
Normalized PAI-1 levels (pg/ml)
p = 4.4e-07
p < 2.22e-16
p = 1.9e-12
6
4
2
T1
T2
T7
Normalized RAGE levels (pg/ml)
p < 2.22e-16
p < 2.22e-16
p < 2.22e-16
8
6
4
T1
T2
T7
Normalized SP-D levels (pg/ml)
p < 2.22e-16
p = 0.11
p < 2.22e-16
50
40
30
20
10
T1
T2
T7

**Figure S2. Manhattan plots of the GWAS association results.**

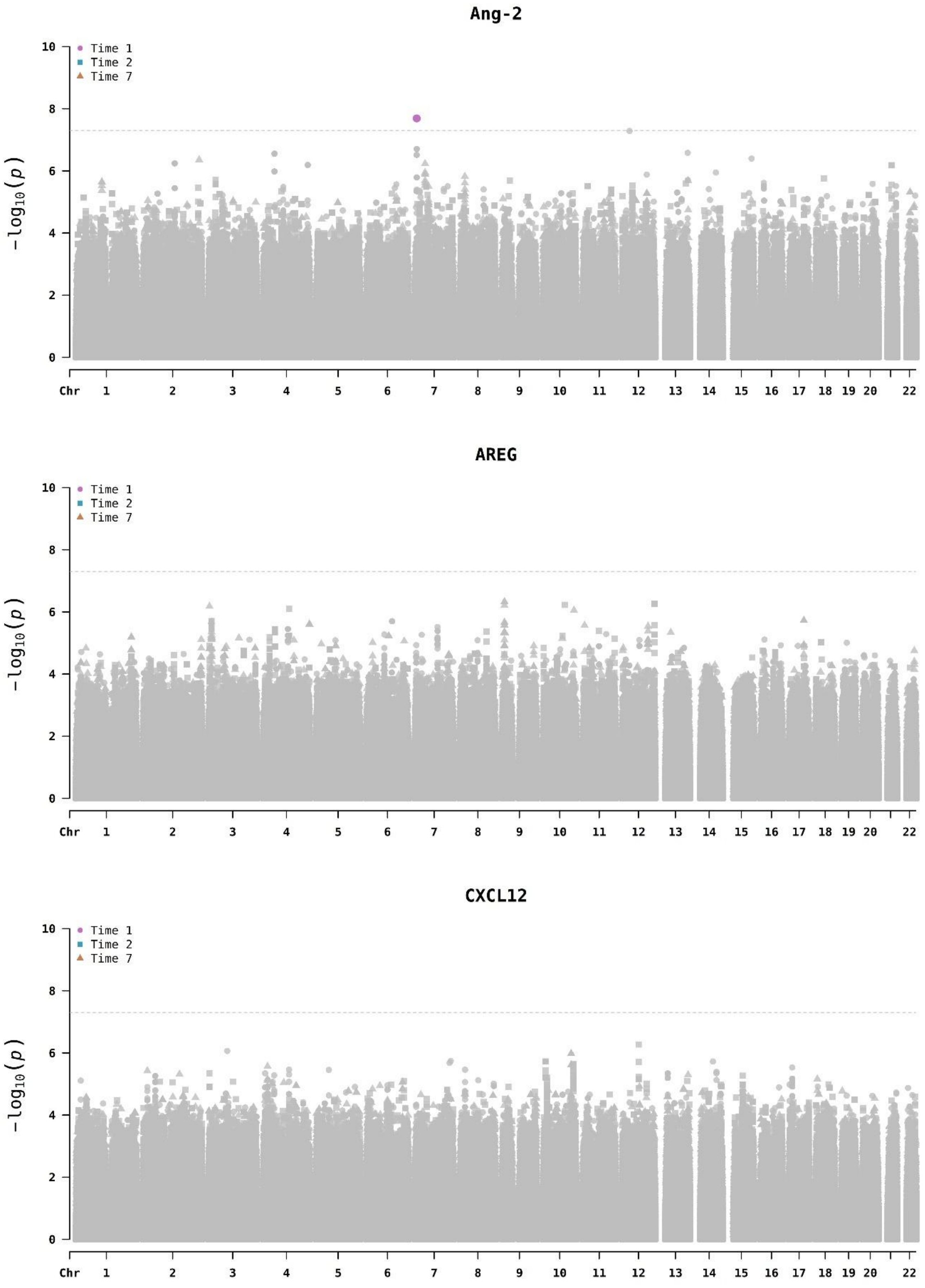

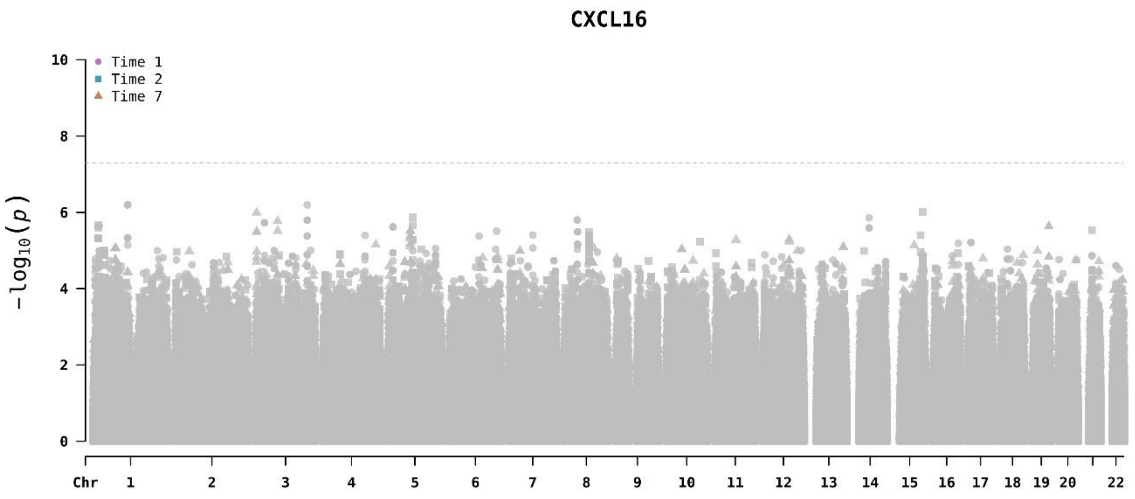
CXCL16
Time 1
Time 2
Time 7
10
8
6
4
2
0
$-\log_{10}(p)$
Chr 1 2 3 4 5 6 7 8 9 10 11 12 13 14 15 16 17 18 19 20 22

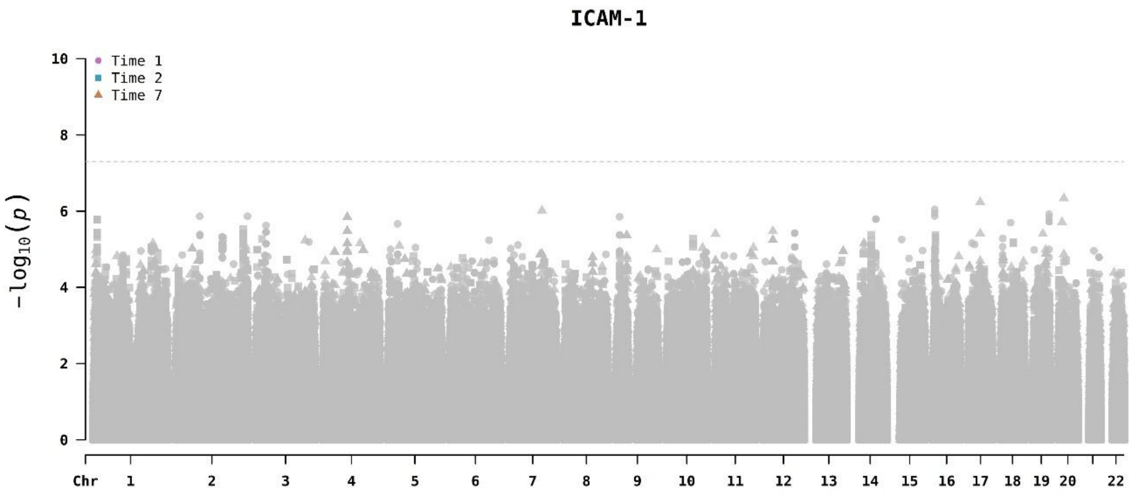
ICAM-1
Time 1
Time 2
Time 7
10
8
6
4
2
0
$-\log_{10}(p)$
Chr 1 2 3 4 5 6 7 8 9 10 11 12 13 14 15 16 17 18 19 20 22

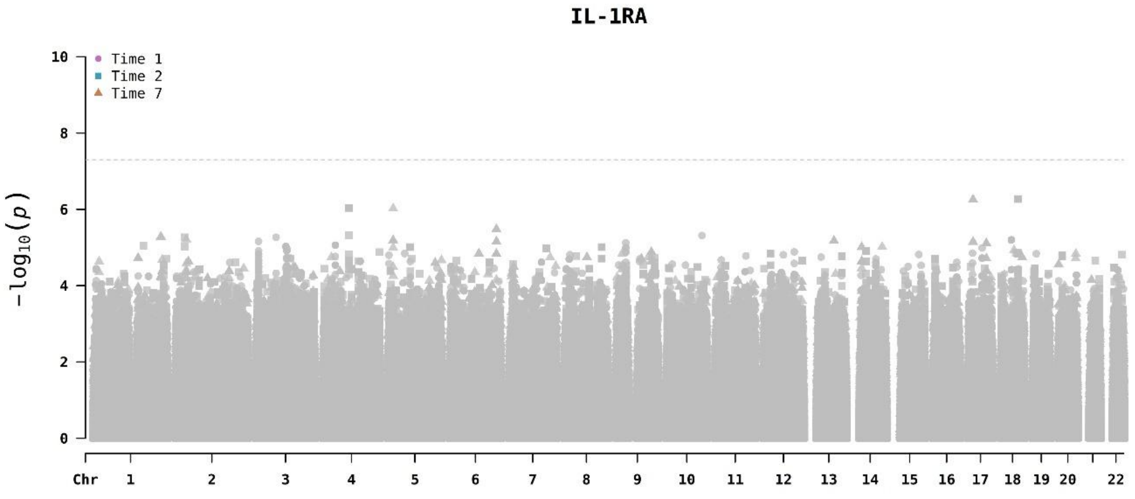
IL-1RA
Time 1
Time 2
Time 7
10
8
6
4
2
0
$-\log_{10}(p)$
Chr 1 2 3 4 5 6 7 8 9 10 11 12 13 14 15 16 17 18 19 20 22

**IL-18**

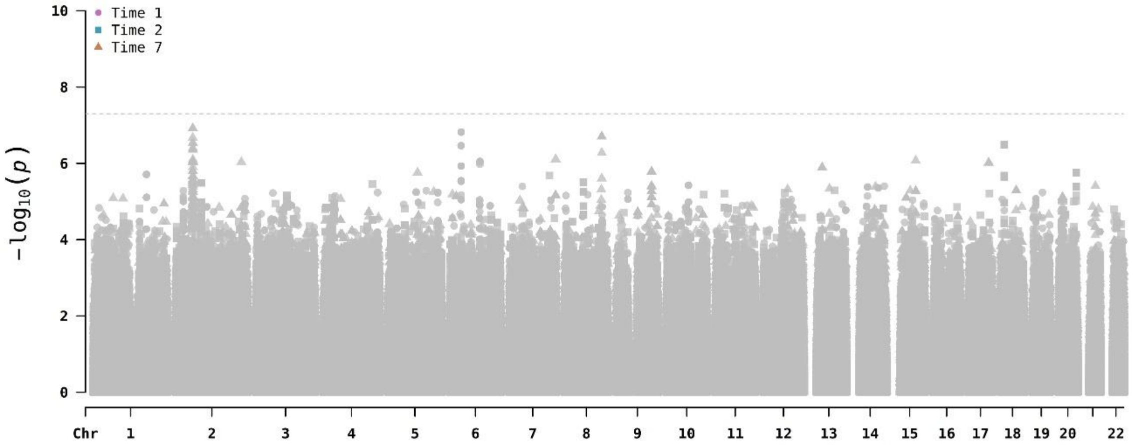

**PAI-1**

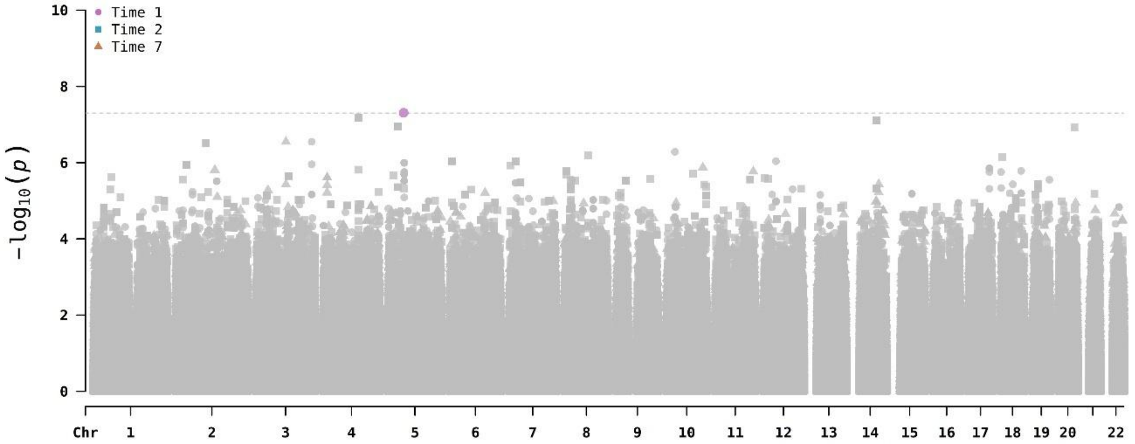

**RAGE**

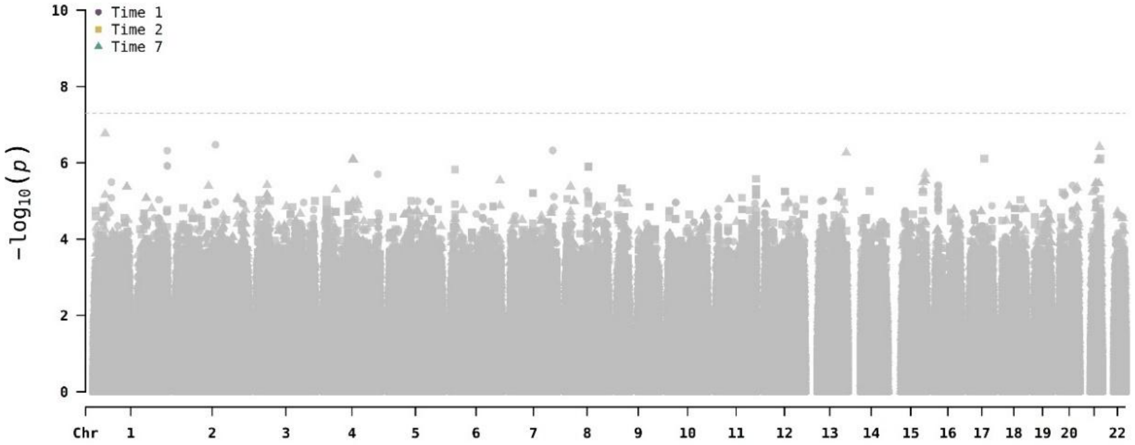

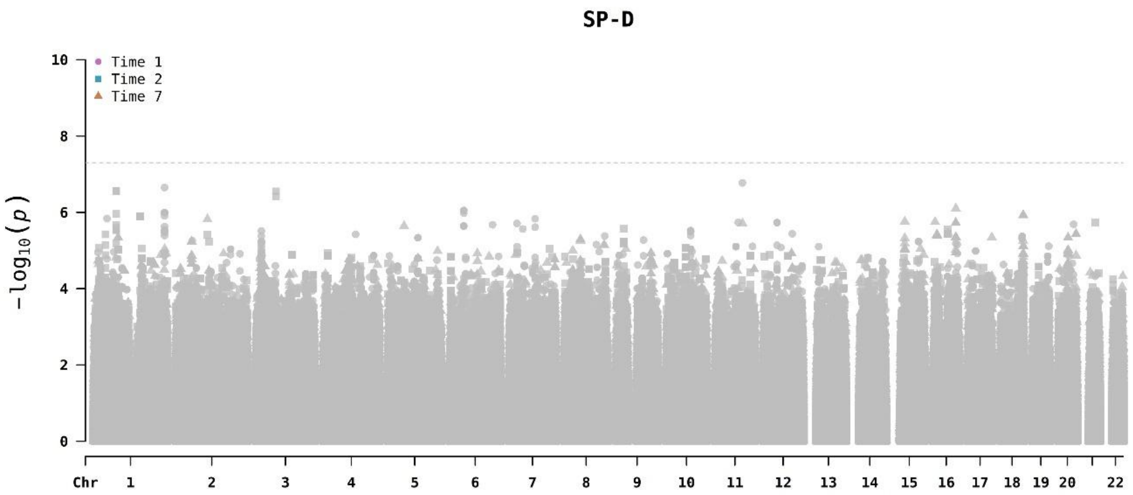

SP-D
Time 1
Time 2
Time 7
10
8
6
4
2
0
$-\log_{10}(p)$
Chr
22

**Figure S3. Regional plots of the association results for the genome-wide significant loci of Ang-2 and PAI-1 levels at T1.** The y-axis shows the -log10(*p*-value) and the x-axis represents the chromosomal positions (GRCh37/hg19). The horizontal dash line indicates the genome-wide significance threshold (*p*-value=5.0x$10^{-8}$). The color scheme of the top left legend represents the linkage disequilibrium (LD) values ($r^2$) with the significant variants, based on the European population data from The 1000 Genomes Project. The plots were generated using LocusZoom (http://locuszoom.org/).

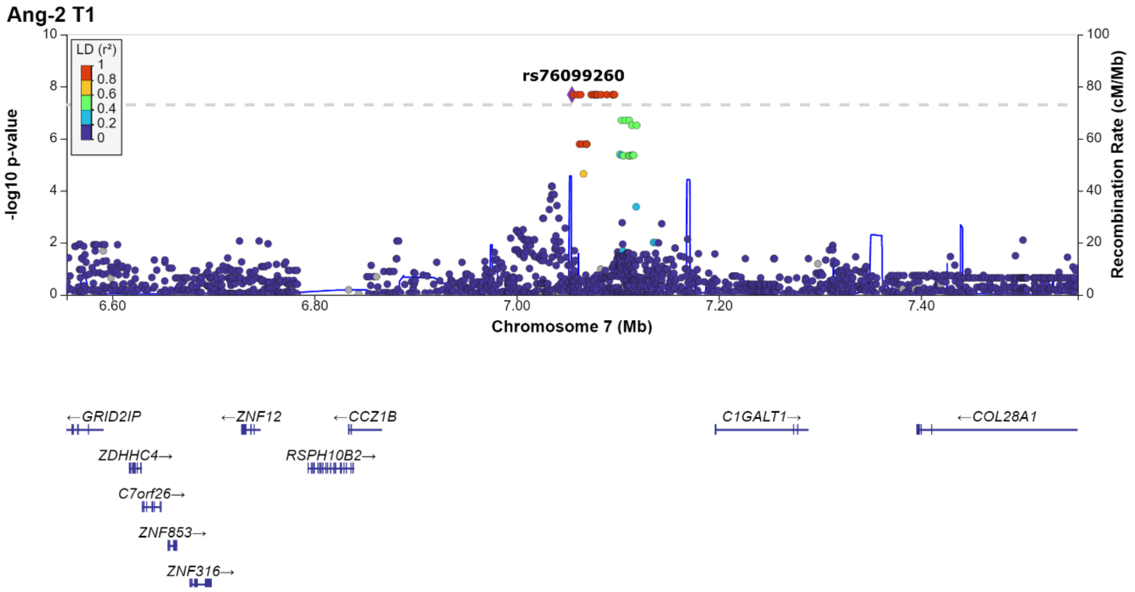

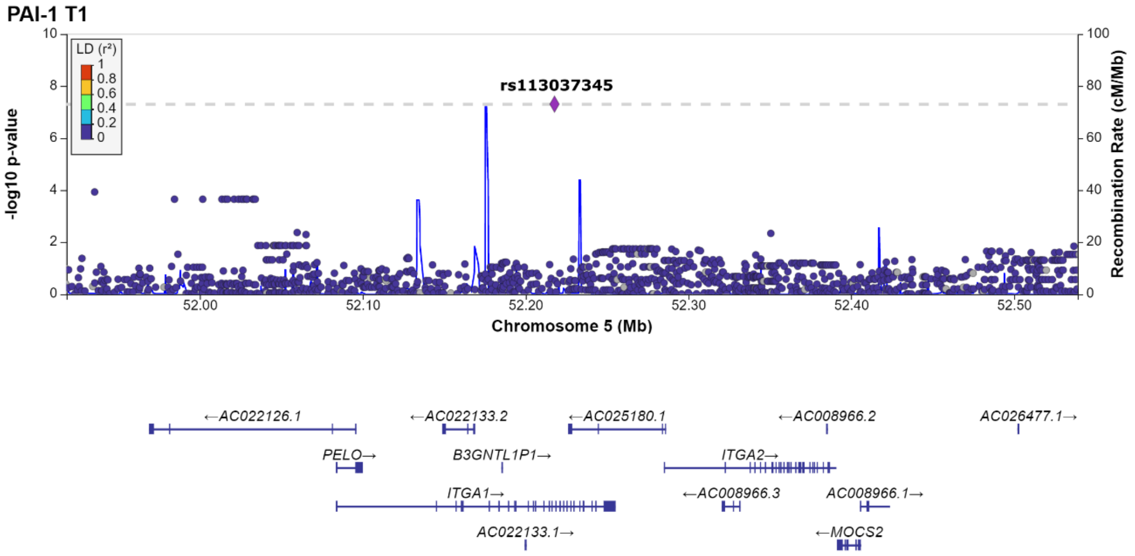

**Figure S4. Manhattan plots of the multi-trait analysis association results.**

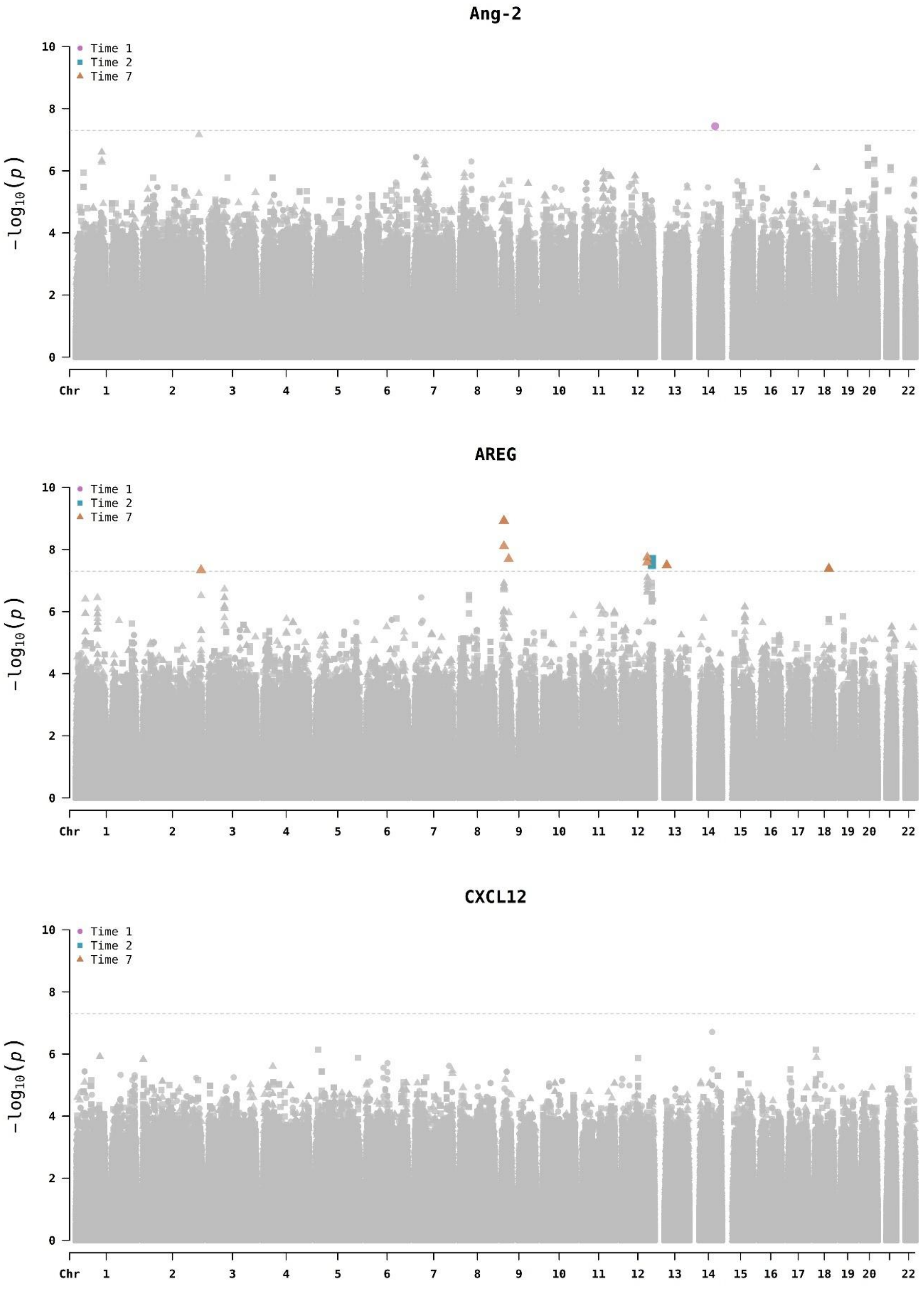

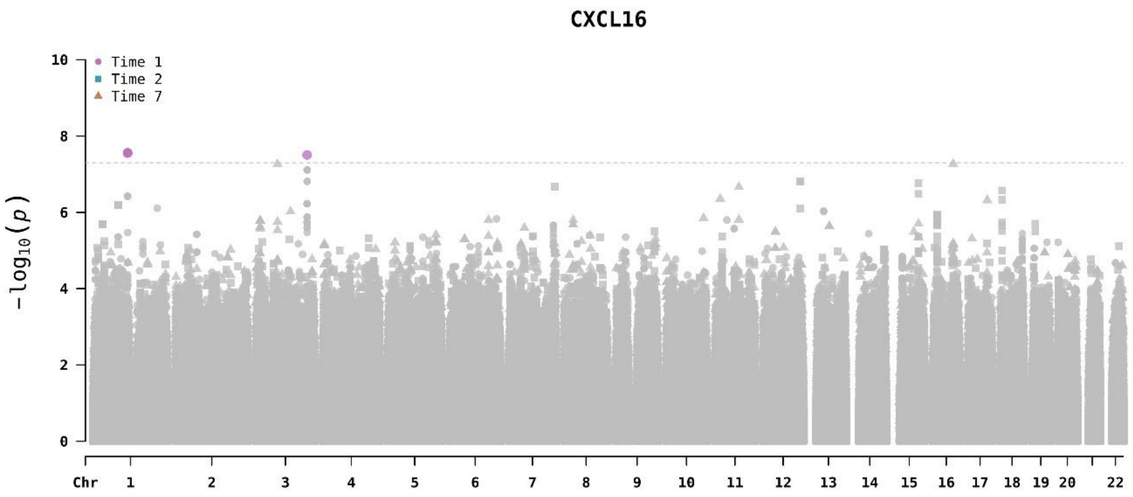
CXCL16
Time 1
Time 2
Time 7
−log10(p)
Chr 1 2 3 4 5 6 7 8 9 10 11 12 13 14 15 16 17 18 19 20 22

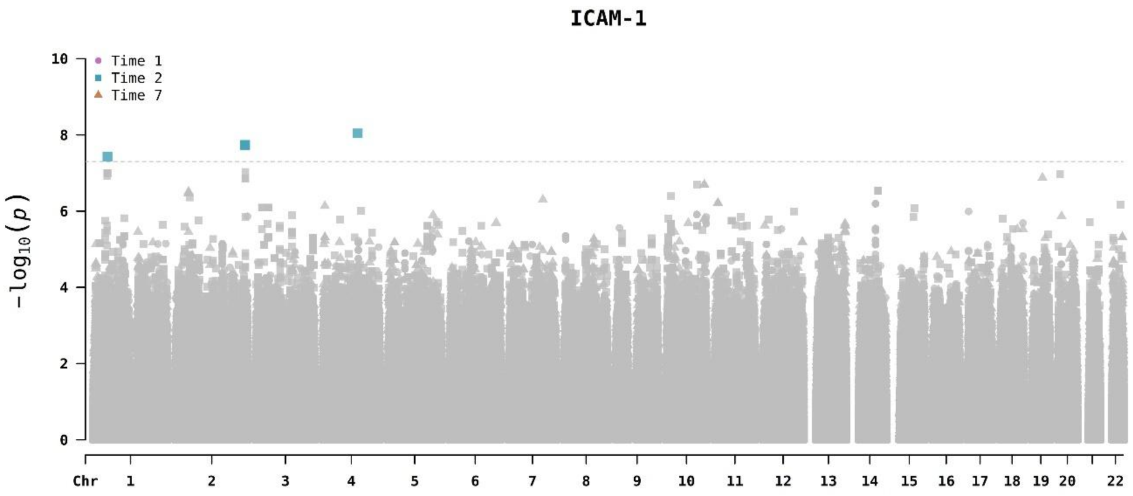
ICAM-1
Time 1
Time 2
Time 7
−log10(p)
Chr 1 2 3 4 5 6 7 8 9 10 11 12 13 14 15 16 17 18 19 20 22

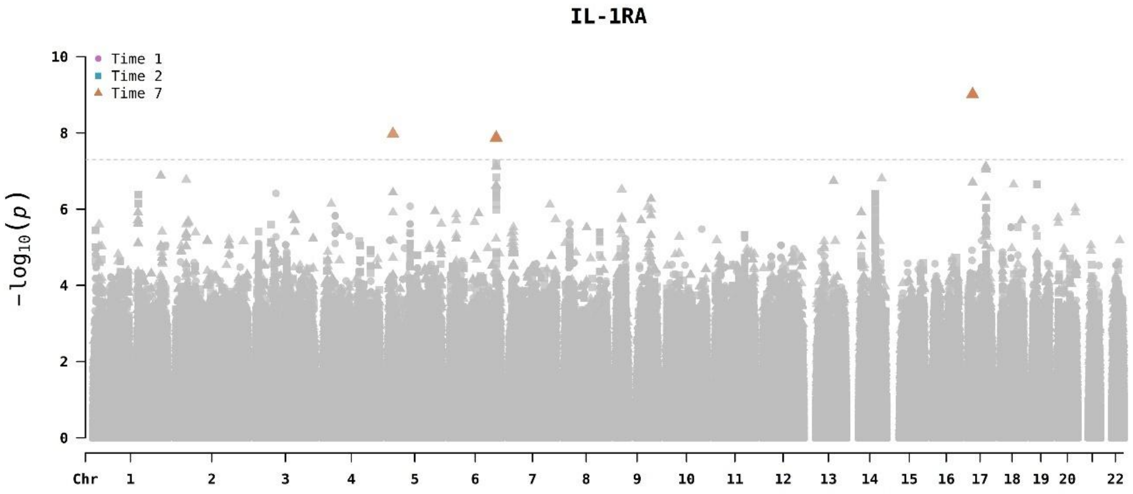
IL-1RA
Time 1
Time 2
Time 7
−log10(p)
Chr 1 2 3 4 5 6 7 8 9 10 11 12 13 14 15 16 17 18 19 20 22

**IL-18**

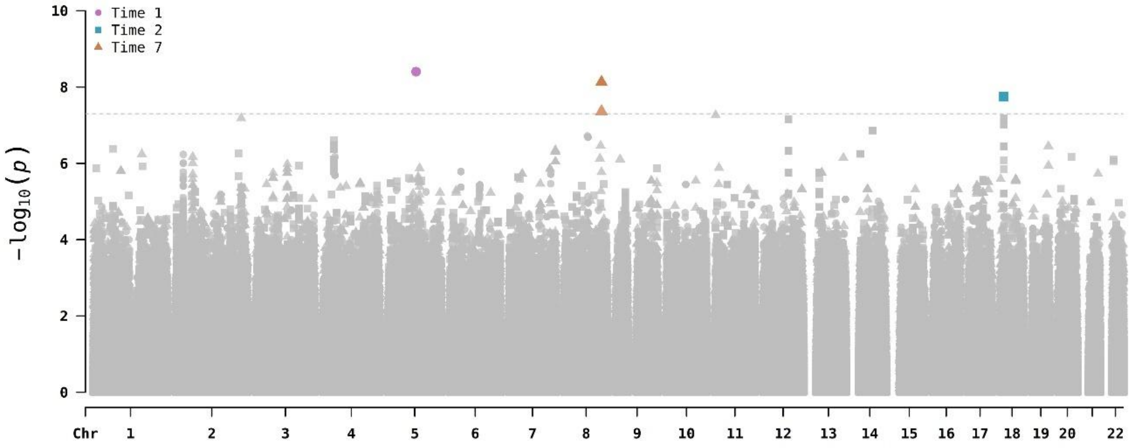

**PAI-1**

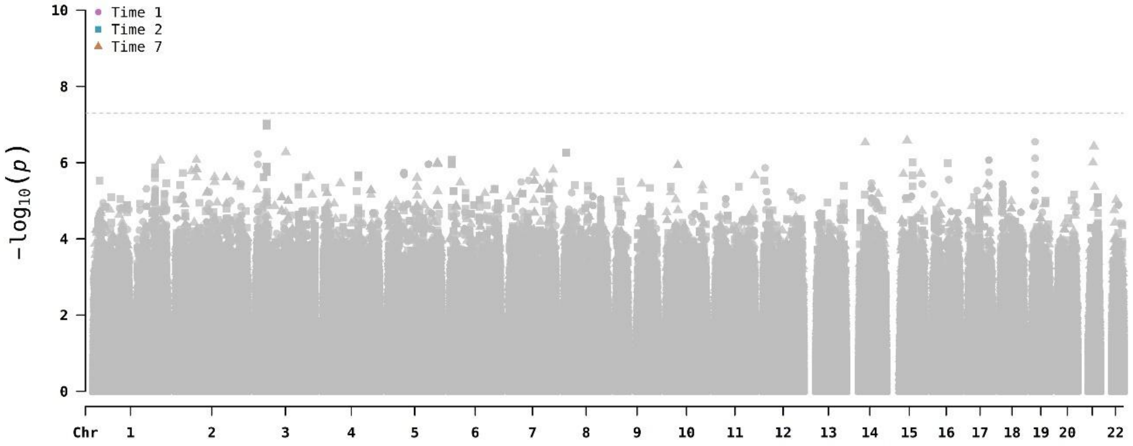

**RAGE**

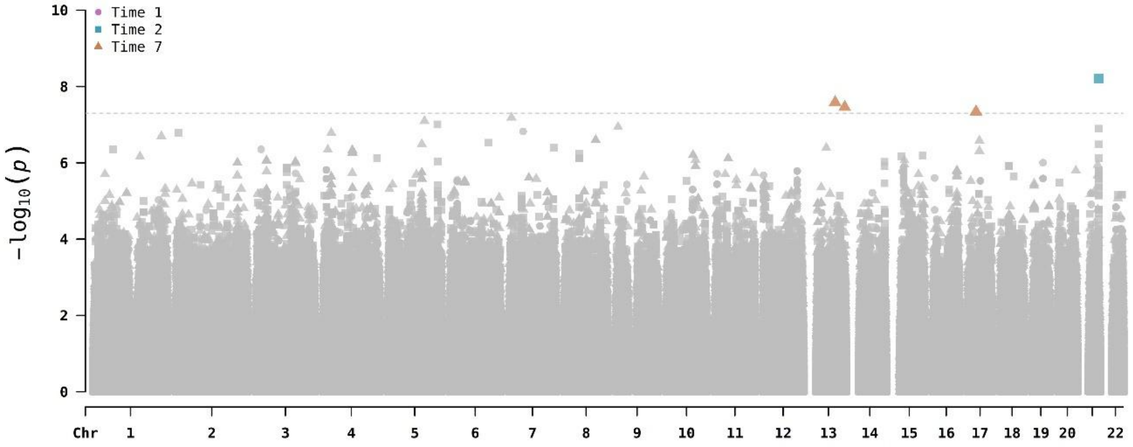

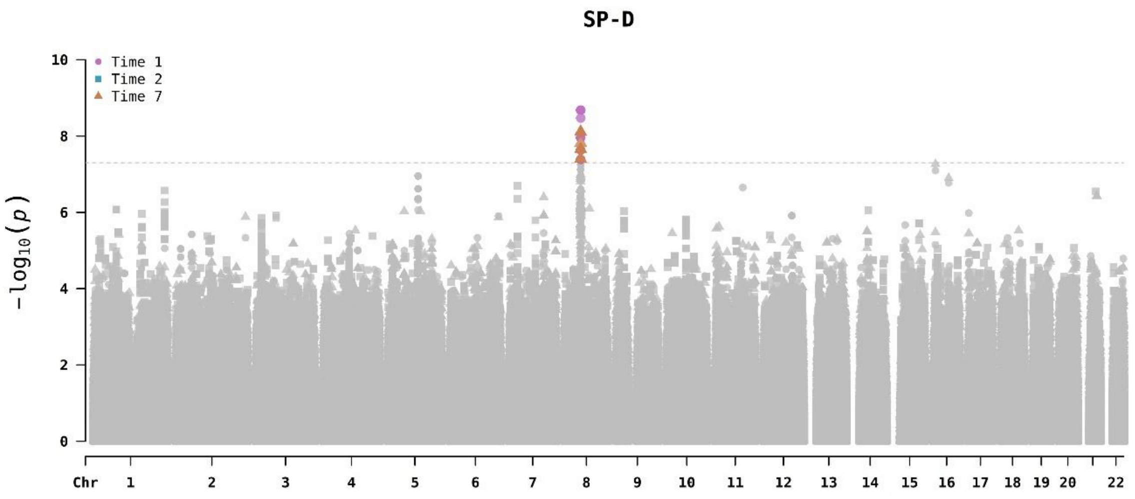
SP-D
Time 1
Time 2
Time 7
10
8
6
4
2
0
−log10(p)
Chr
22

**Figure S5. Regional plots of the significant loci identified in the multi-trait analyses.** The y-axis shows the -log10(*p*-value) and the x-axis represents the chromosomal positions (GRCh37/hg19). The horizontal dash line indicates the genome-wide significance threshold (*p*-value=5.0x10$^{-8}$). The color scheme of the top left legend represents the linkage disequilibrium (LD) values ($r^2$) with the significant variants, based on the European population data from The 1000 Genomes Project. The plots were generated using LocusZoom (http://locuszoom.org/).

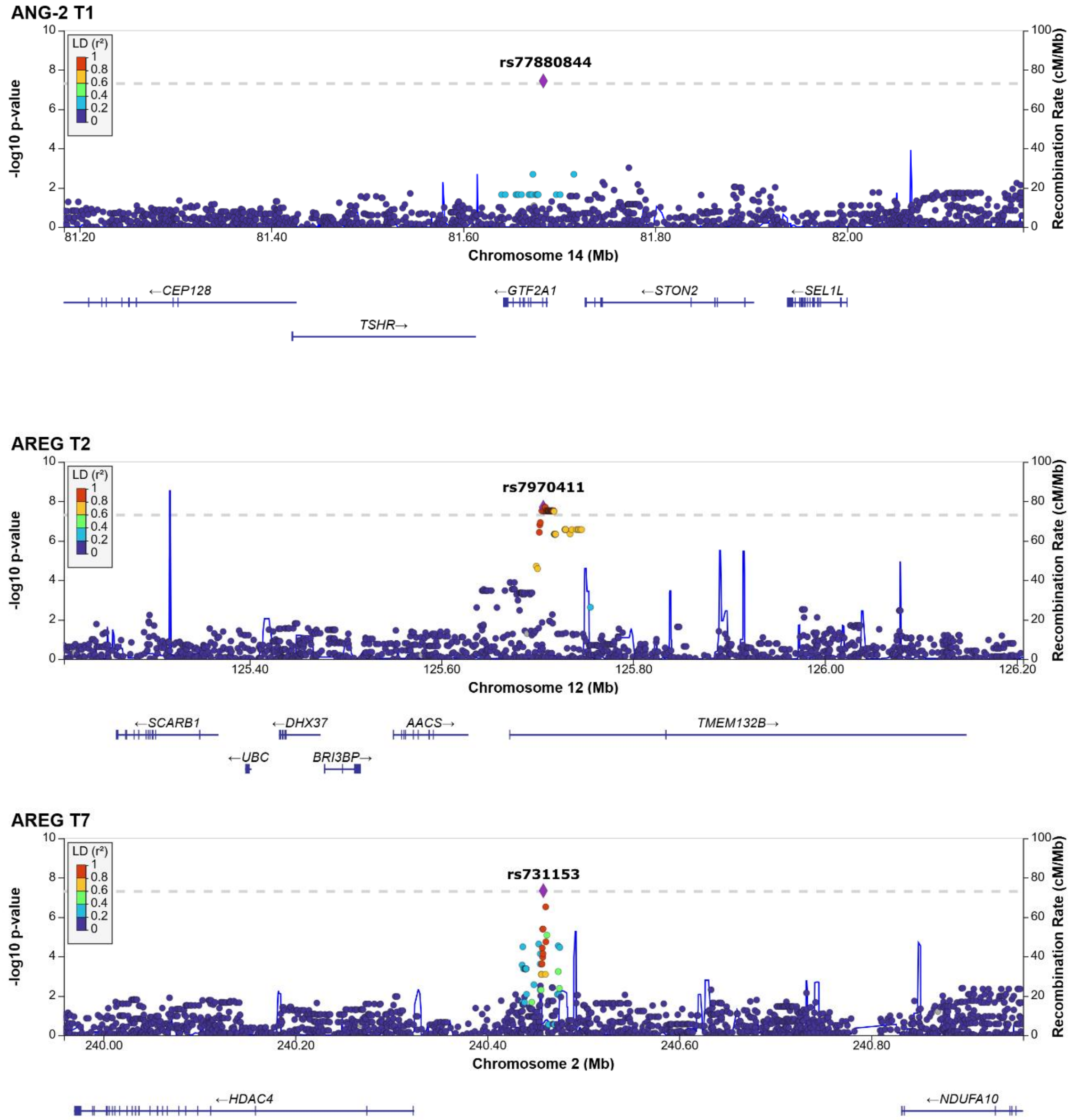

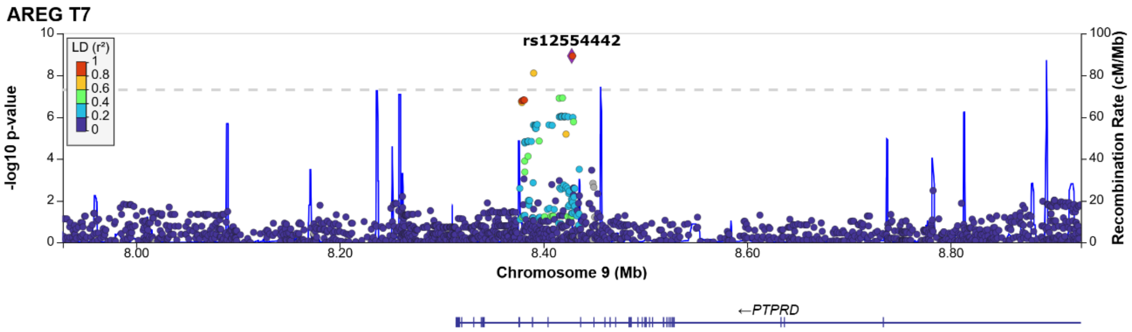
AREG T7
rs12554442
LD (r²)
1
0.8
0.6
0.4
0.2
0
-log10 p-value
Recombination Rate (cM/Mb)
Chromosome 9 (Mb)
8.00
8.20
8.40
8.60
8.80
←PTPRD

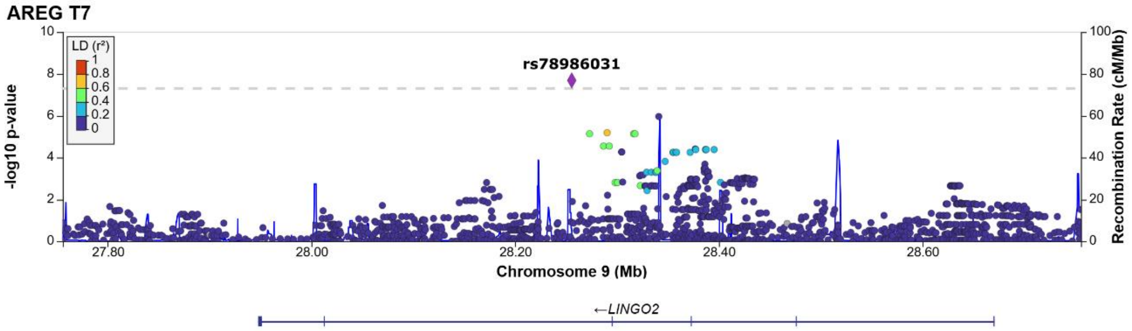
AREG T7
rs78986031
LD (r²)
-log10 p-value
Recombination Rate (cM/Mb)
Chromosome 9 (Mb)
27.80
28.00
28.20
28.40
28.60
←LINGO2

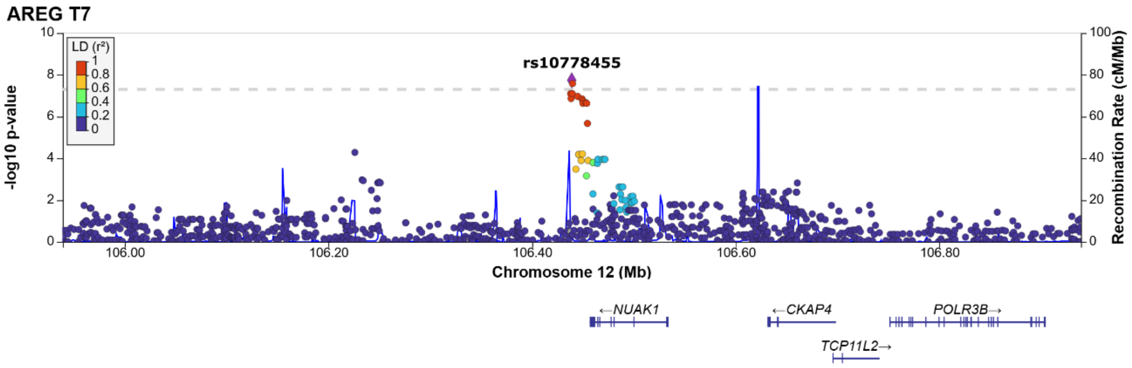
AREG T7
rs10778455
LD (r²)
-log10 p-value
Recombination Rate (cM/Mb)
Chromosome 12 (Mb)
106.00
106.20
106.40
106.60
106.80
←NUAK1
←CKAP4
POLR3B→
TCP11L2→

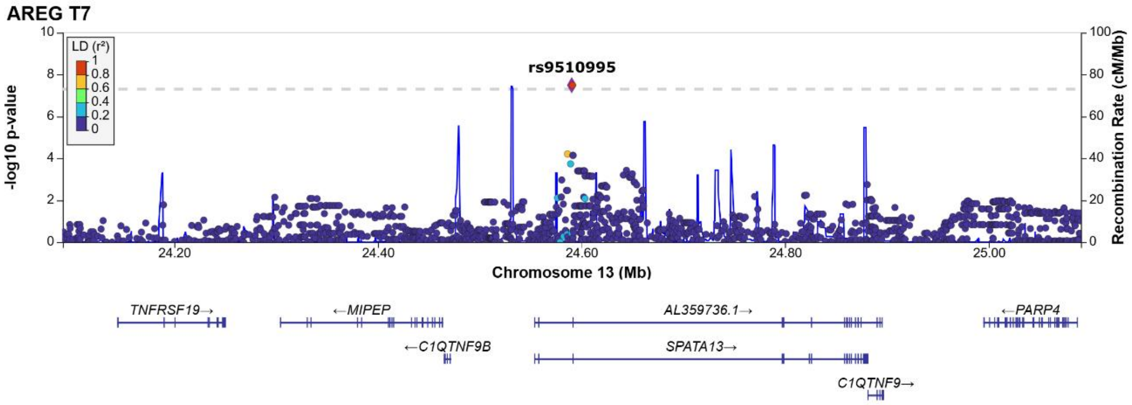
AREG T7
rs9510995
LD (r²)
-log10 p-value
Recombination Rate (cM/Mb)
Chromosome 13 (Mb)
24.20
24.40
24.60
24.80
25.00
TNFRSF19→
←MIPEP
AL359736.1→
←PARP4
←C1QTNF9B
SPATA13→
C1QTNF9→

**AREG T7**

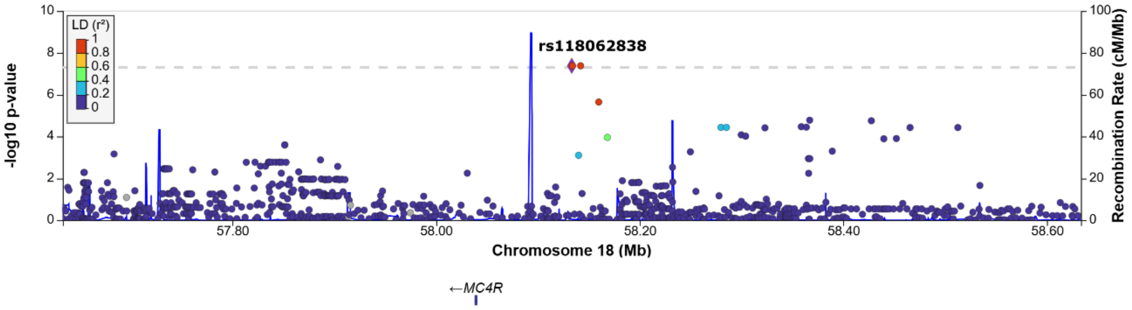

**CXCL16 T1**

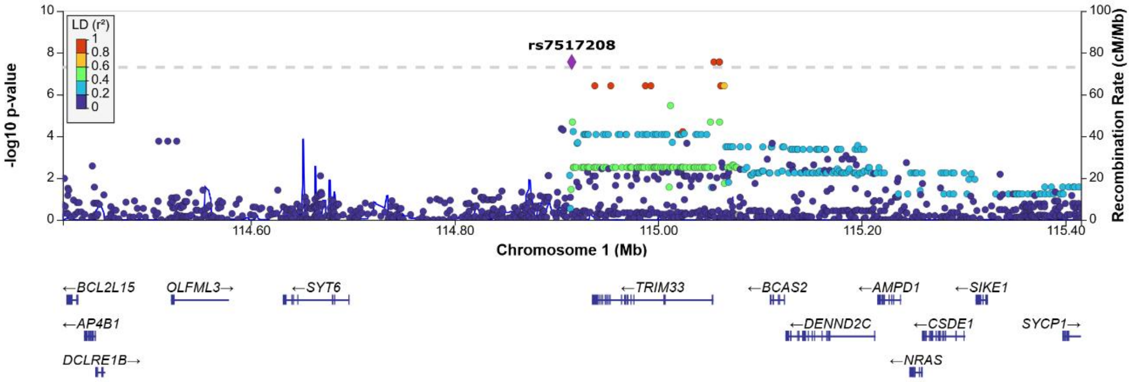

**CXCL16 T1**

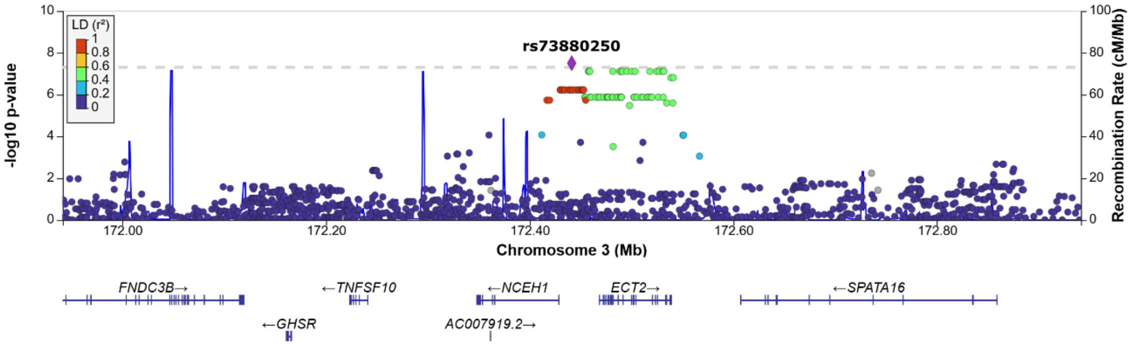

**ICAM1 T2**

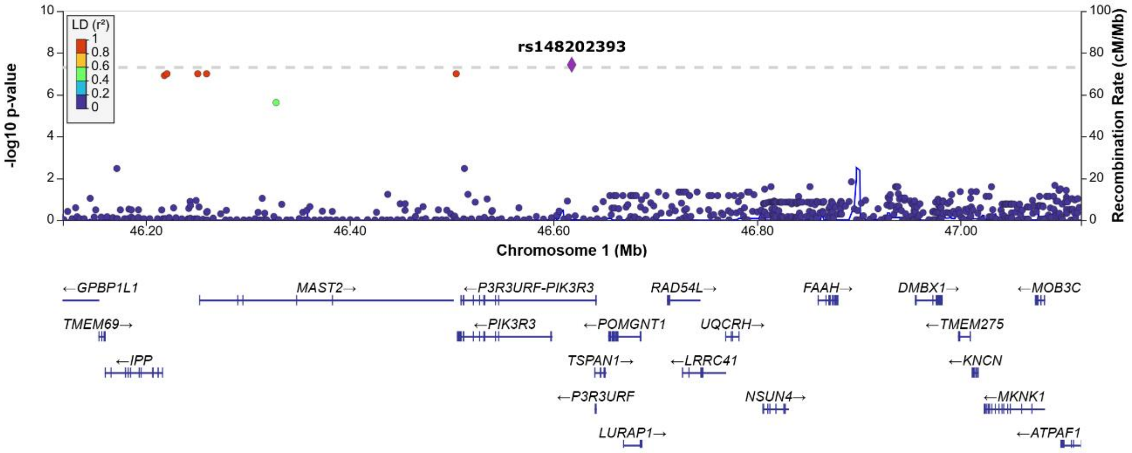

ICAM1 T2

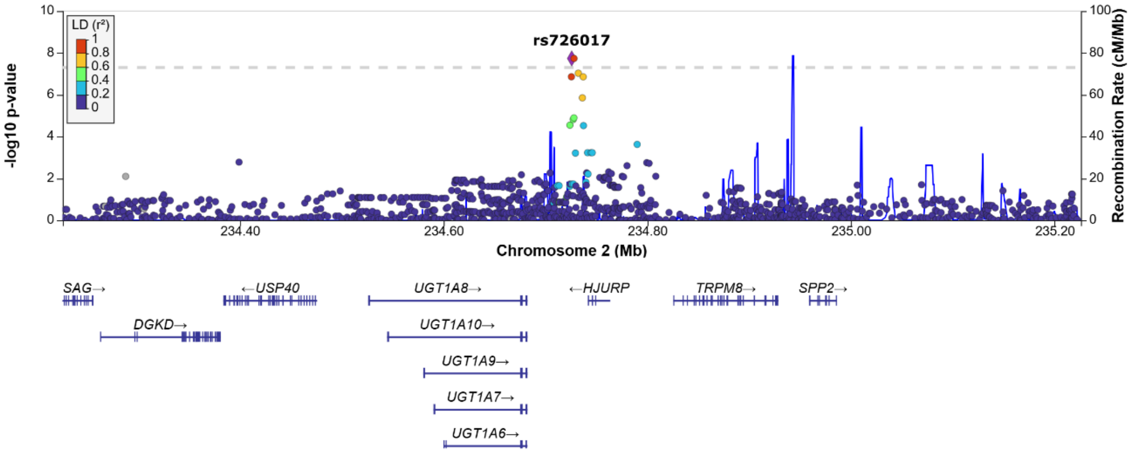

ICAM1 T2

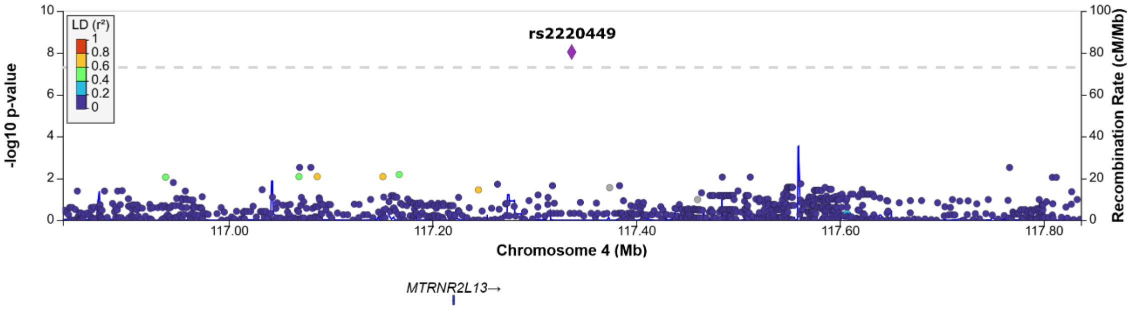

IL1RA T7

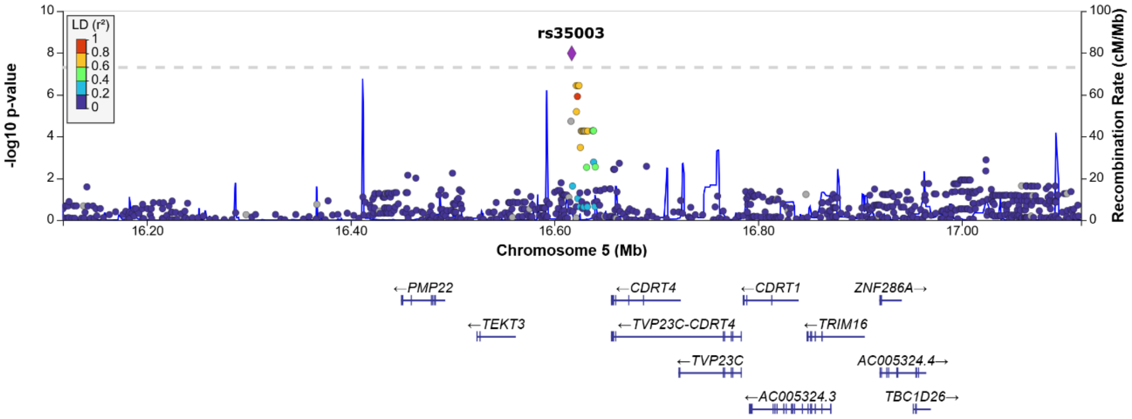

IL1RA T7

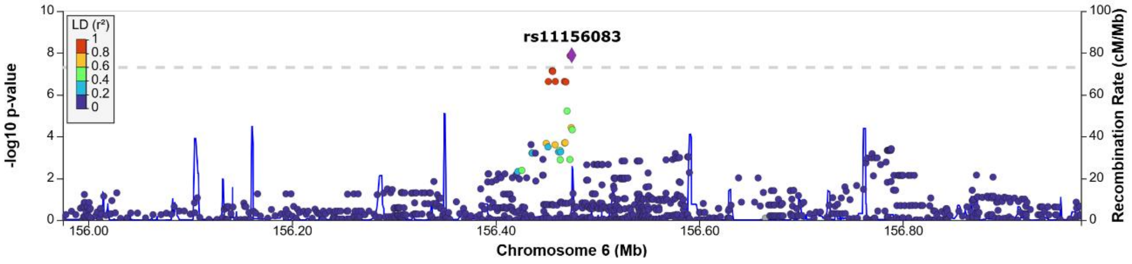

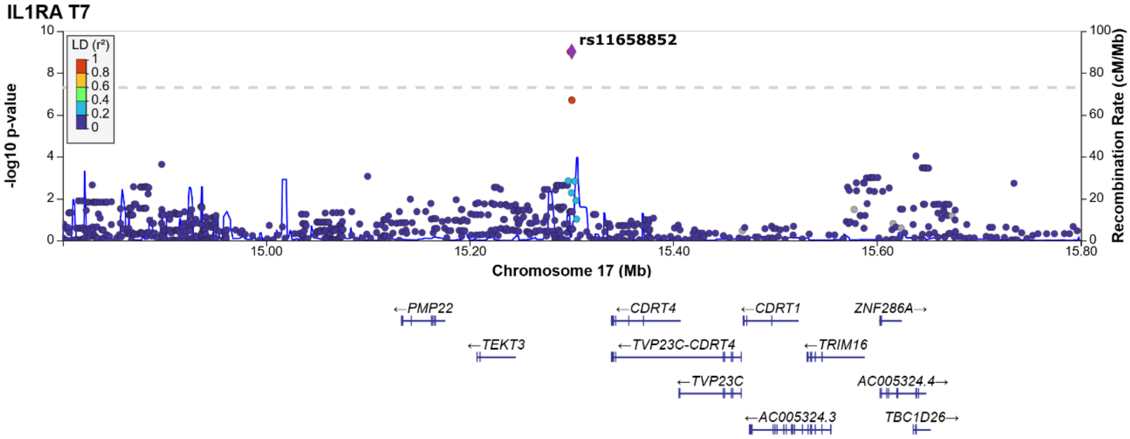

IL1RA T7
LD (r²)
rs11658852
-log10 p-value
Recombination Rate (cM/Mb)
Chromosome 17 (Mb)
←PMP22
←TEKT3
←CDRT4
←TVP23C-CDRT4
←TVP23C
←AC005324.3
←CDRT1
←TRIM16
ZNF286A→
AC005324.4→
TBC1D26→

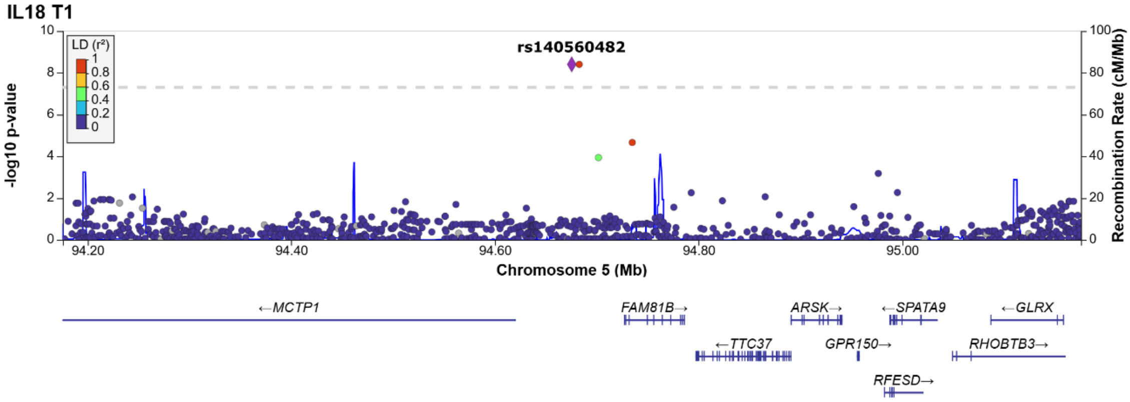

IL18 T1
LD (r²)
rs140560482
-log10 p-value
Recombination Rate (cM/Mb)
Chromosome 5 (Mb)
←MCTP1
FAM81B→
←TTC37
ARSK→
GPR150→
RFESD→
←SPATA9
←GLRX
RHOBTB3→

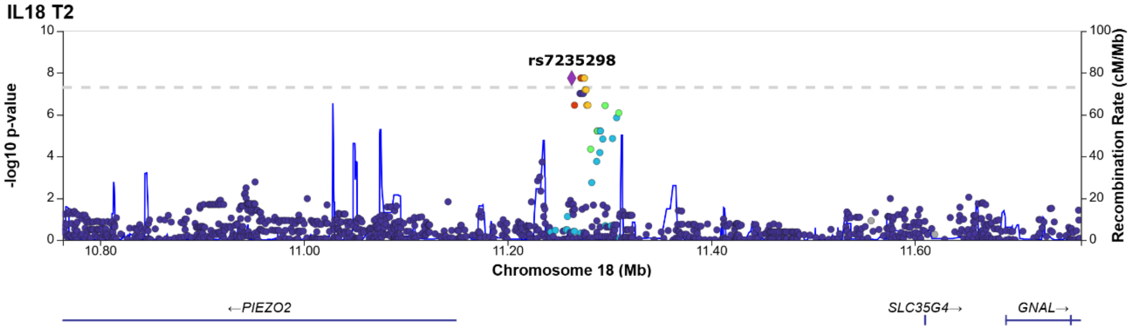

IL18 T2
rs7235298
-log10 p-value
Recombination Rate (cM/Mb)
Chromosome 18 (Mb)
←PIEZO2
SLC35G4→
GNAL→

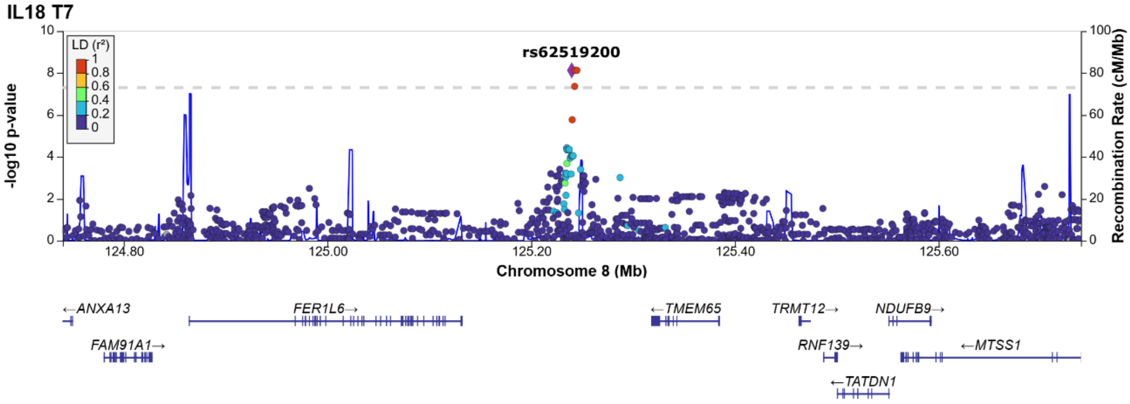

IL18 T7
LD (r²)
rs62519200
-log10 p-value
Recombination Rate (cM/Mb)
Chromosome 8 (Mb)
←ANXA13
FAM91A1→
FER1L6→
←TMEM65
TRMT12→
RNF139→
←TATDN1
NDUFB9→
←MTSS1

### RAGE T2

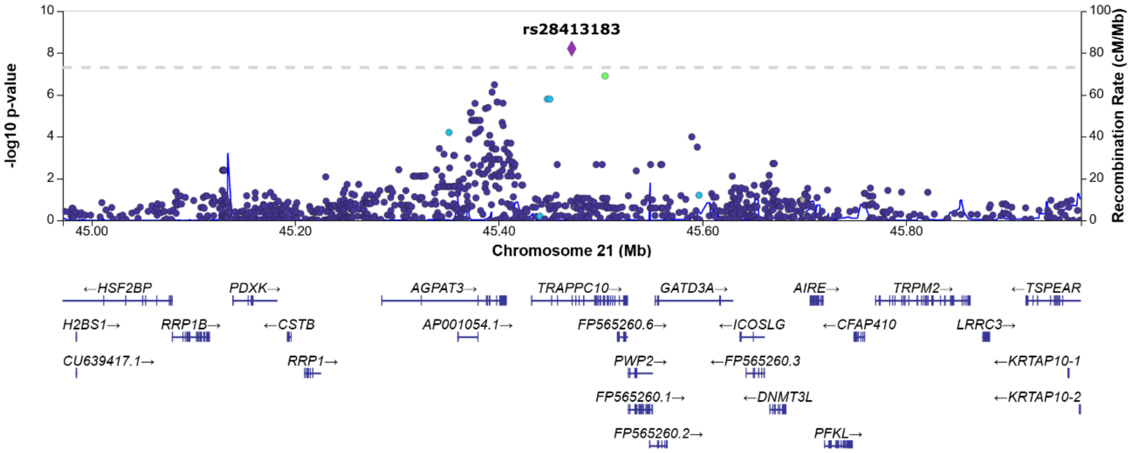

### RAGE T7

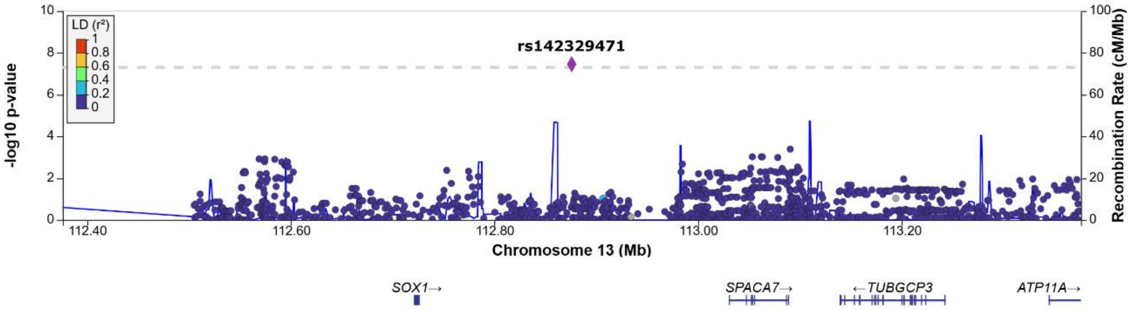

### RAGE T7

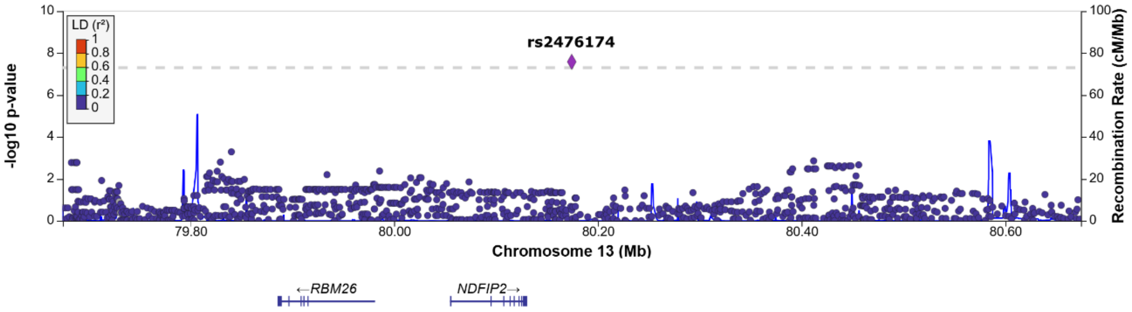

### RAGE T7

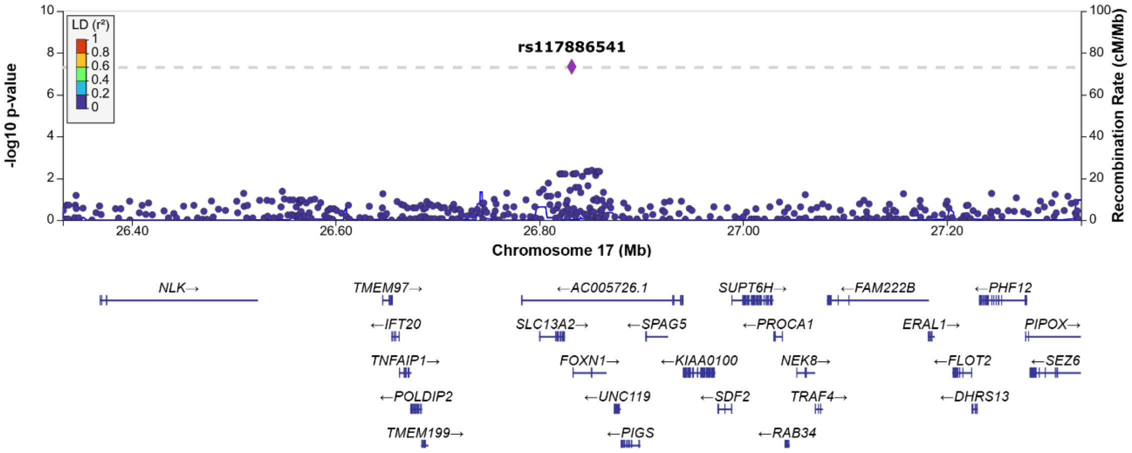

SPD T1

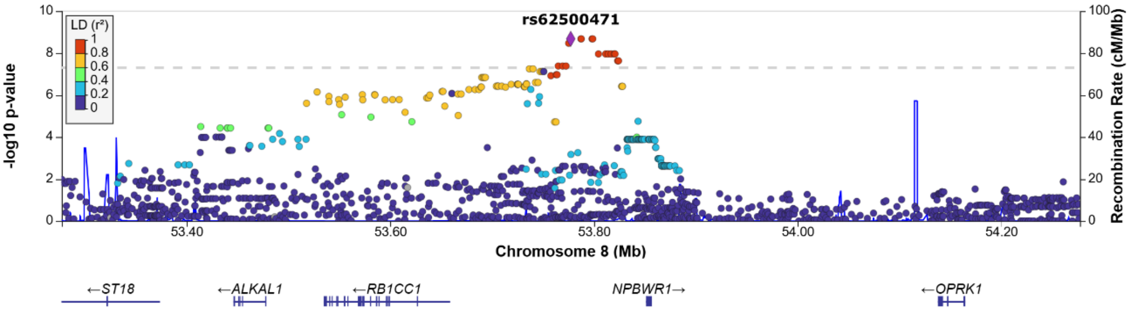
rs62500471
LD (r²)
1
0.8
0.6
0.4
0.2
0
-log10 p-value
Recombination Rate (cM/Mb)
Chromosome 8 (Mb)
53.40
53.60
53.80
54.00
54.20
←ST18
←ALKAL1
←RB1CC1
NPBWR1→
←OPRK1

SPD T7

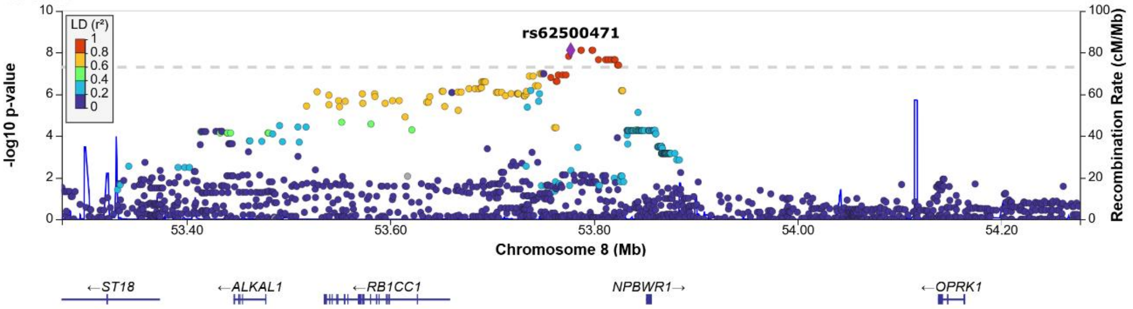
rs62500471
LD (r²)
1
0.8
0.6
0.4
0.2
0
-log10 p-value
Recombination Rate (cM/Mb)
Chromosome 8 (Mb)
53.40
53.60
53.80
54.00
54.20
←ST18
←ALKAL1
←RB1CC1
NPBWR1→
←OPRK1

**Figure S6. Quantile-Quantile (Q-Q) plots of GWAS results.**

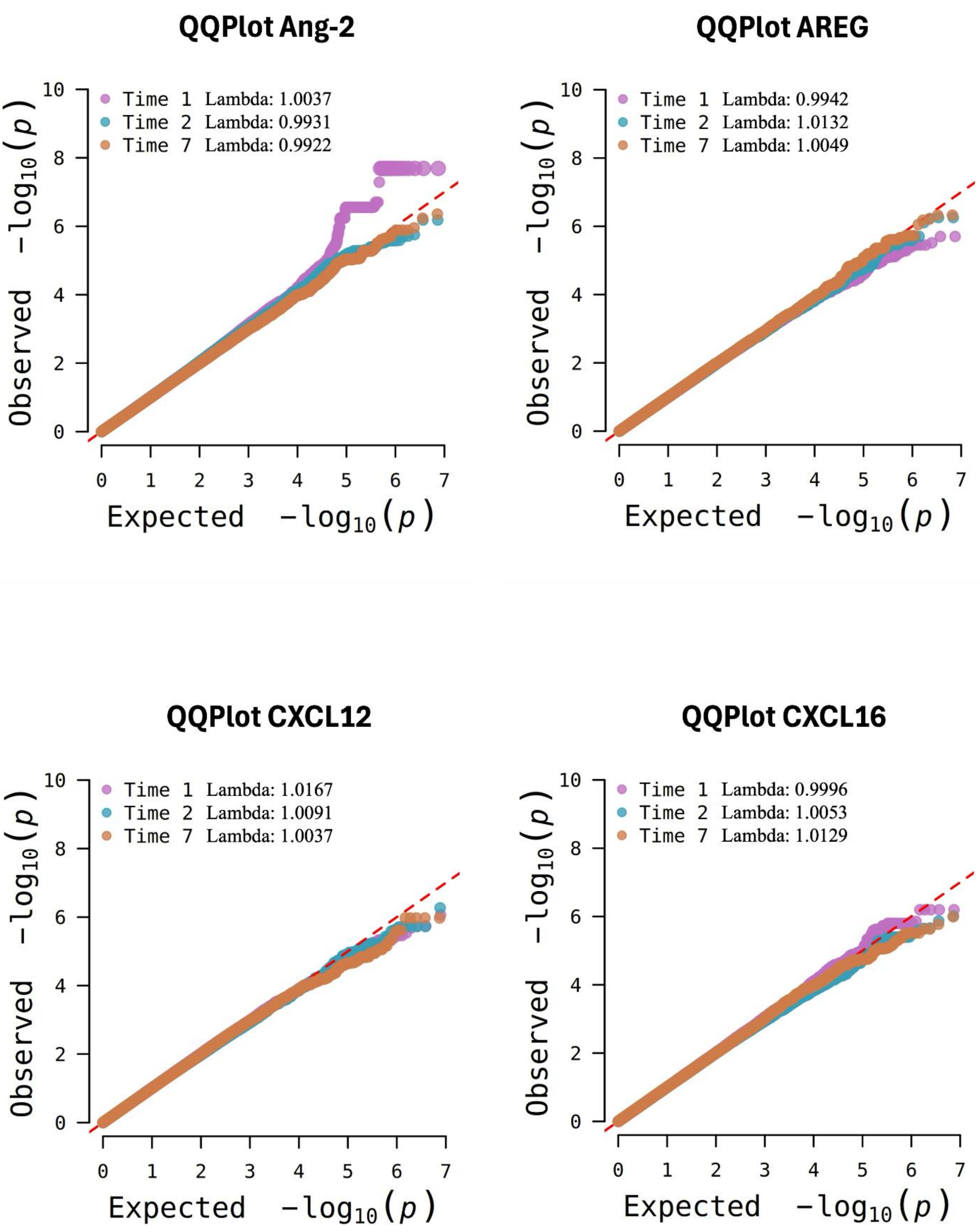

**QQPlot ICAM-1**

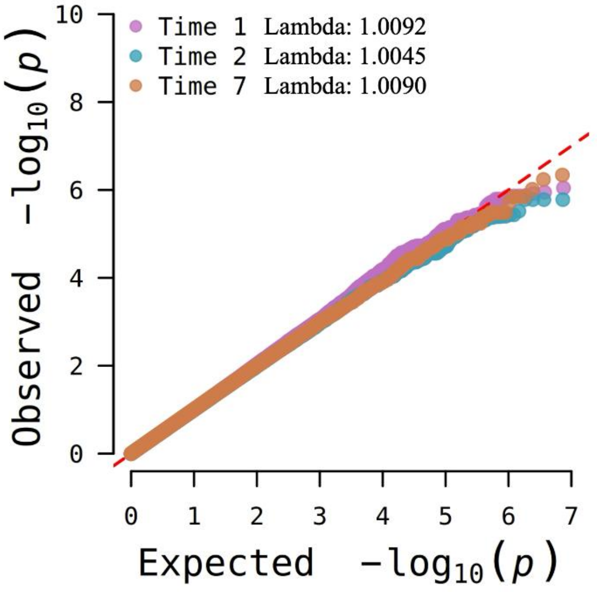

**QQPlot IL-1RA**

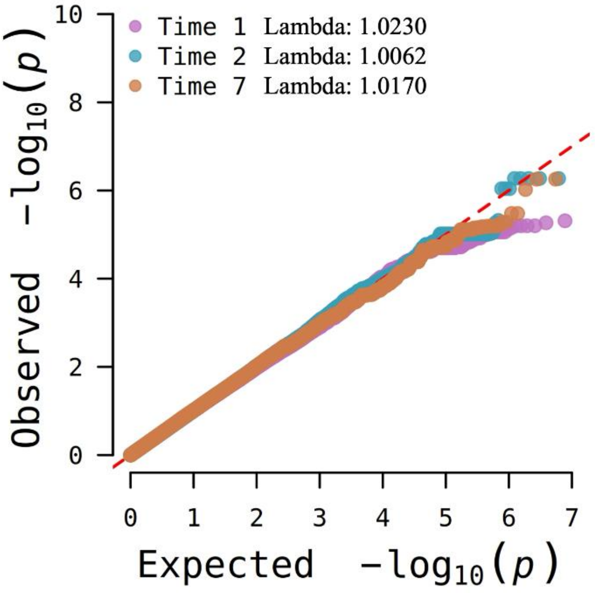

**QQPlot IL-18**

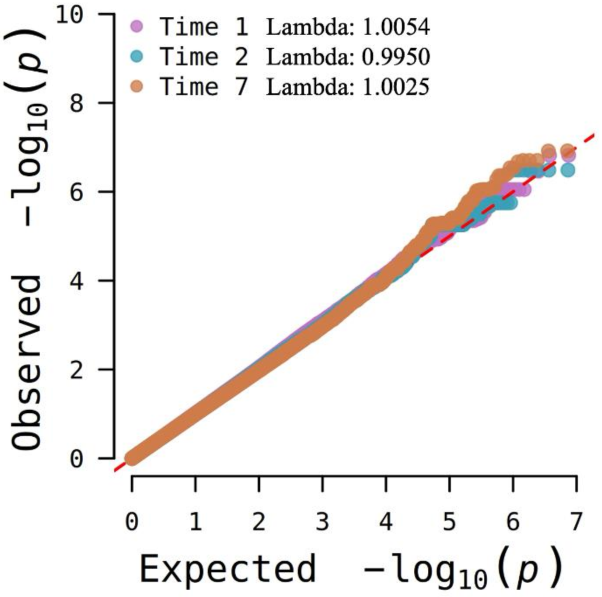

**QQPlot PAI-1**

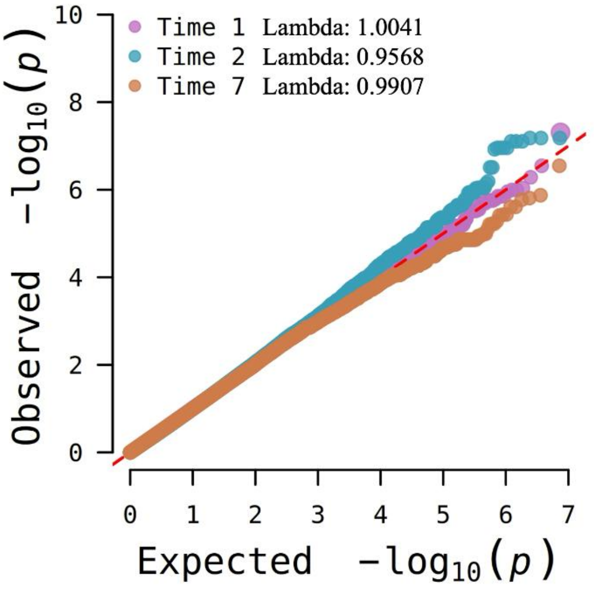

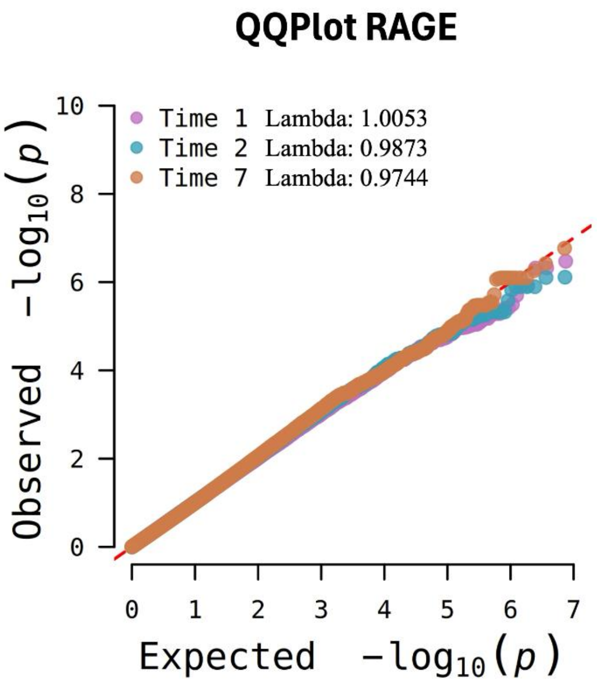
QQPlot RAGE
Time 1 Lambda: 1.0053
Time 2 Lambda: 0.9873
Time 7 Lambda: 0.9744
Observed −log10(p)
Expected −log10(p)

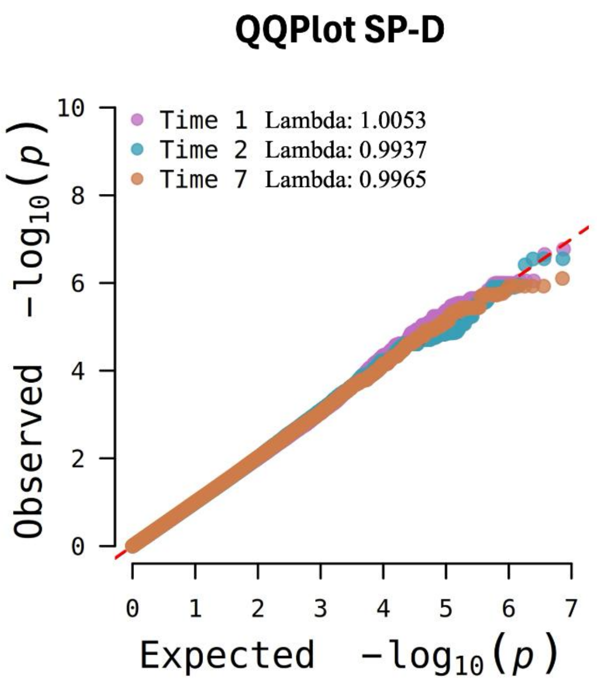
QQPlot SP-D
Time 1 Lambda: 1.0053
Time 2 Lambda: 0.9937
Time 7 Lambda: 0.9965
Observed −log10(p)
Expected −log10(p)

**Figure S7. Quantile-Quantile (Q-Q) plots of multi-trait analyses.**

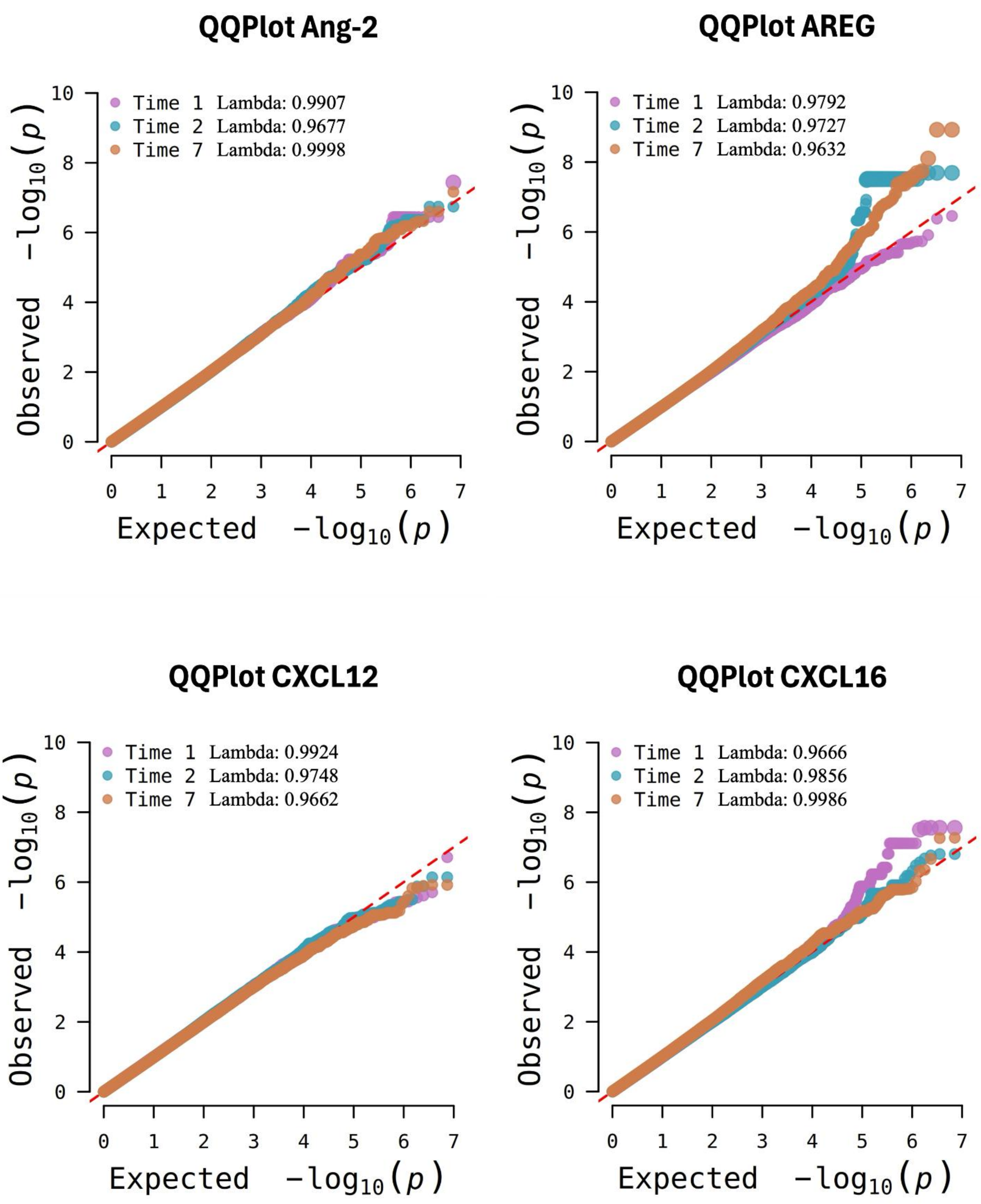

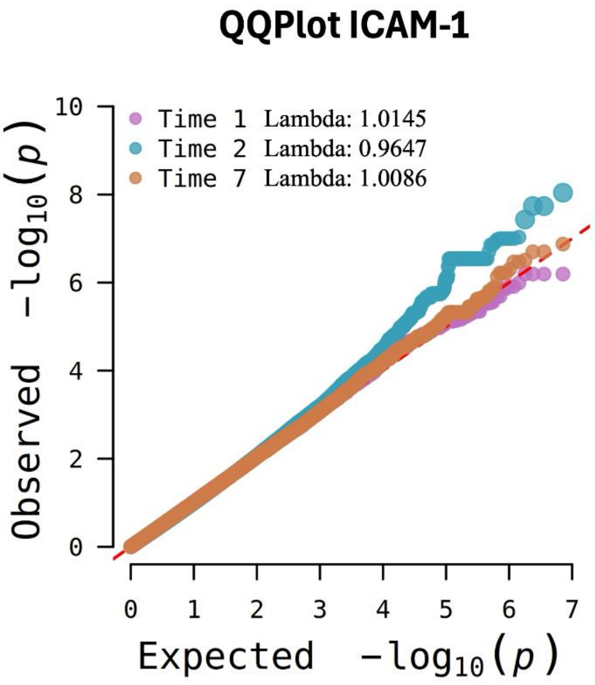
QQPlot ICAM-1
Time 1 Lambda: 1.0145
Time 2 Lambda: 0.9647
Time 7 Lambda: 1.0086
Observed $-\log_{10}(p)$
Expected $-\log_{10}(p)$

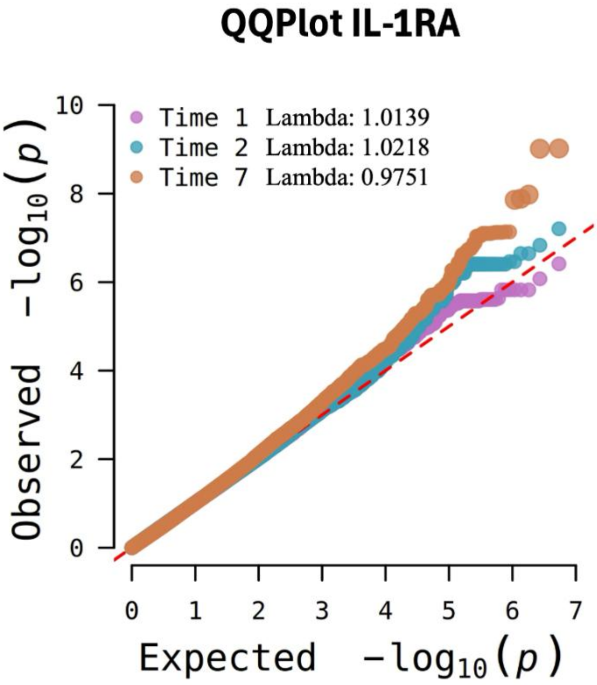
QQPlot IL-1RA
Time 1 Lambda: 1.0139
Time 2 Lambda: 1.0218
Time 7 Lambda: 0.9751
Observed $-\log_{10}(p)$
Expected $-\log_{10}(p)$

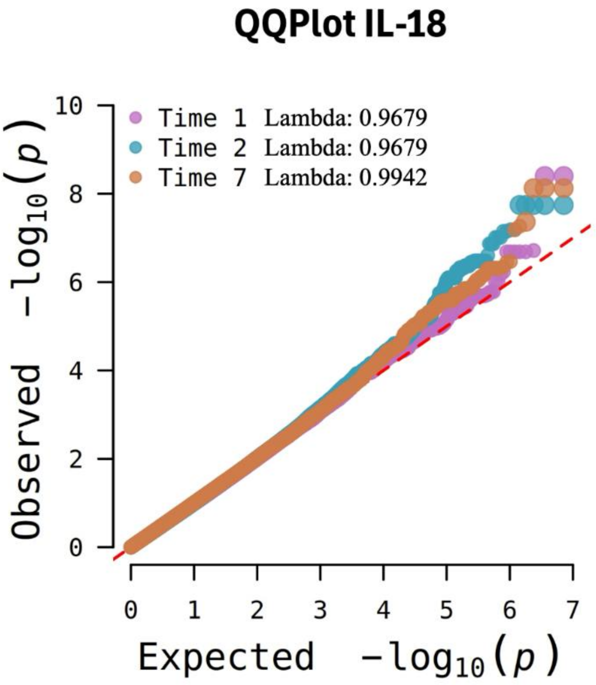
QQPlot IL-18
Time 1 Lambda: 0.9679
Time 2 Lambda: 0.9679
Time 7 Lambda: 0.9942
Observed $-\log_{10}(p)$
Expected $-\log_{10}(p)$

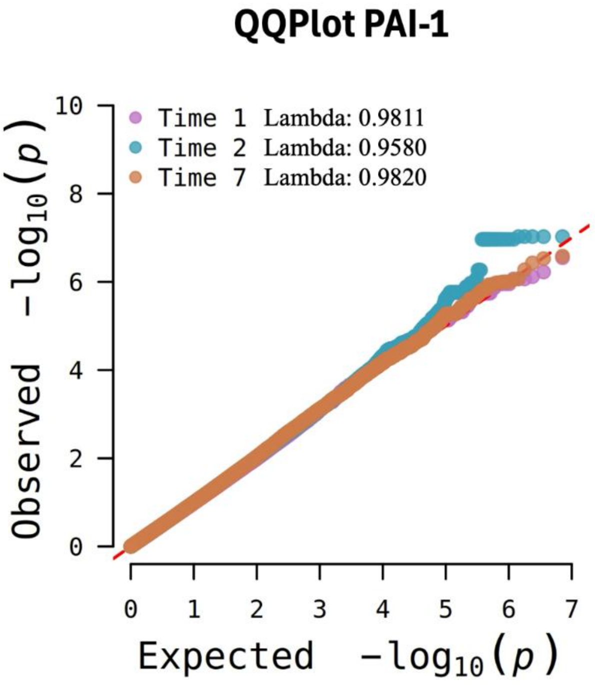
QQPlot PAI-1
Time 1 Lambda: 0.9811
Time 2 Lambda: 0.9580
Time 7 Lambda: 0.9820
Observed $-\log_{10}(p)$
Expected $-\log_{10}(p)$

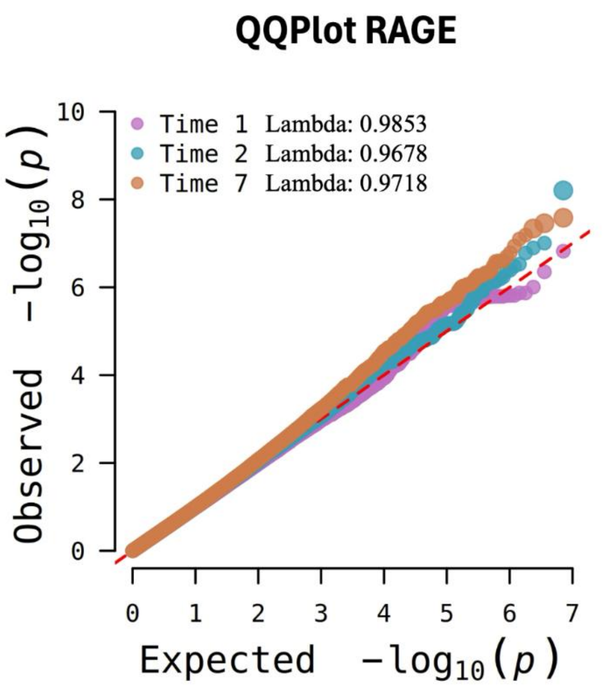
QQPlot RAGE
Time 1 Lambda: 0.9853
Time 2 Lambda: 0.9678
Time 7 Lambda: 0.9718
Observed −log10(p)
Expected −log10(p)

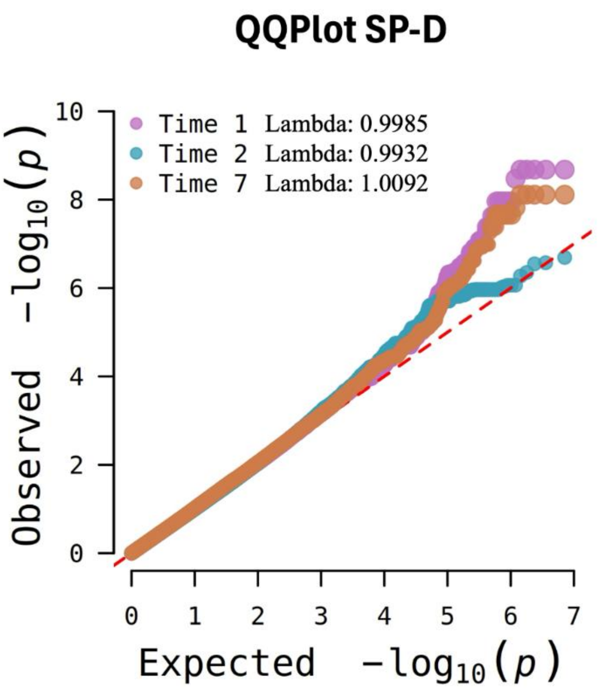
QQPlot SP-D
Time 1 Lambda: 0.9985
Time 2 Lambda: 0.9932
Time 7 Lambda: 1.0092
Observed −log10(p)
Expected −log10(p)

**Figure S8. Regional plots of the Bayesian fine mapping and credible set of significant loci.** The x-axis represents the chromosomal positions (GRCh37/hg19), and the y-axis shows the -log10($p$-value). The horizontal dashed line represents the genome-wide significance threshold ($p$-value=5.0x$10^{-8}$). The significant pQTLs are highlighted in orange and the pQTLs included in the credible set with 95% of confidence are in purple. T1: protein levels obtained within 24 hours after sepsis diagnosis; T2: protein levels obtained within 48-72 hours after sepsis diagnosis; T7: protein levels obtained 7 days after sepsis diagnosis.

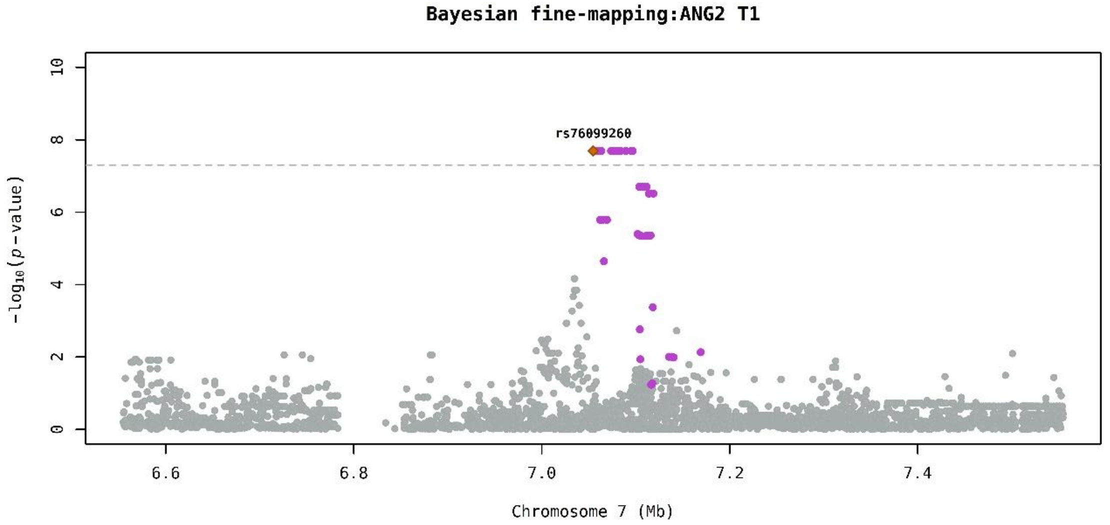

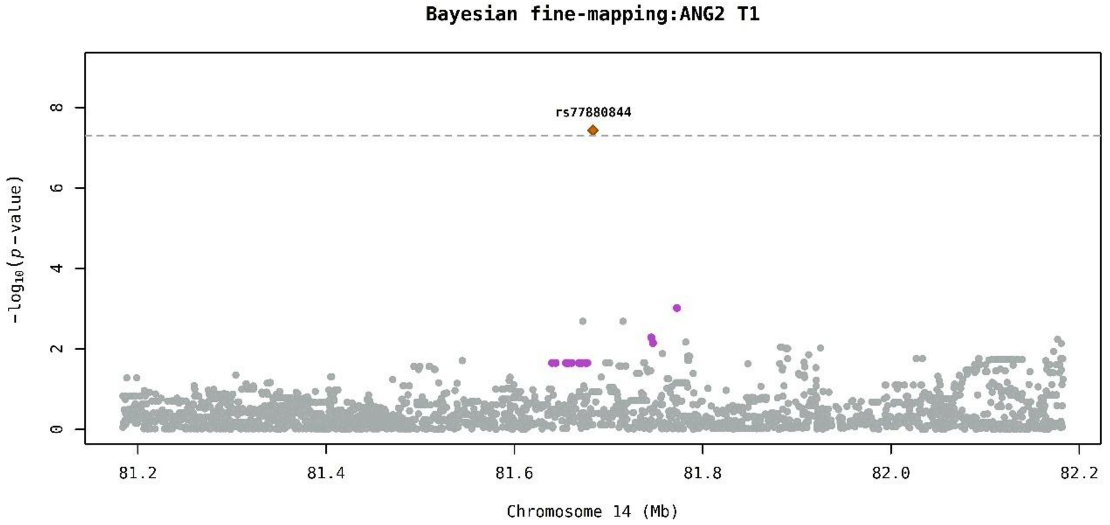

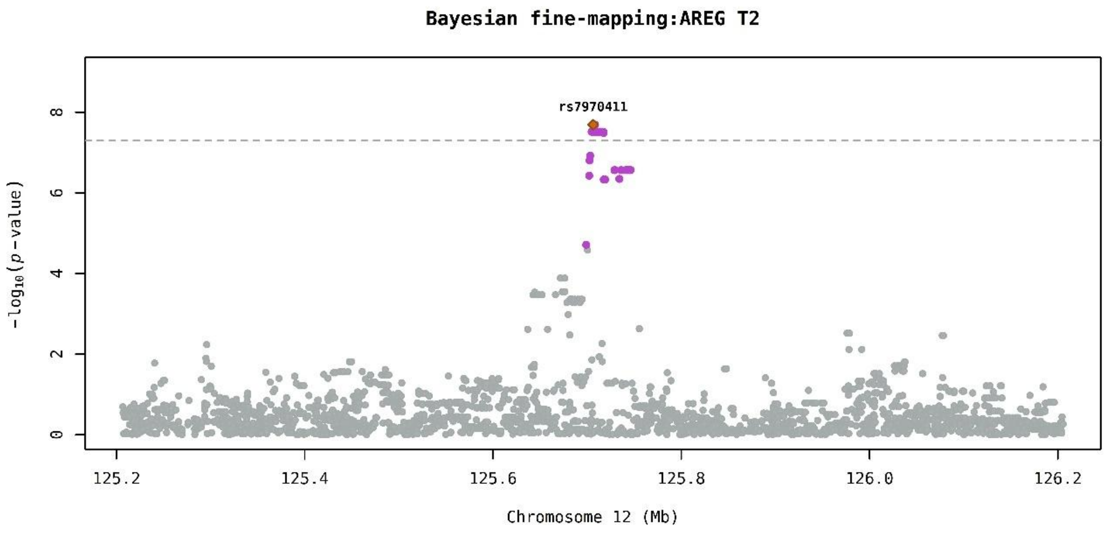
Bayesian fine-mapping:AREG T2
rs7970411
$-\log_{10}(p\text{-value})$
0
2
4
6
8
125.2
125.4
125.6
125.8
126.0
126.2
Chromosome 12 (Mb)

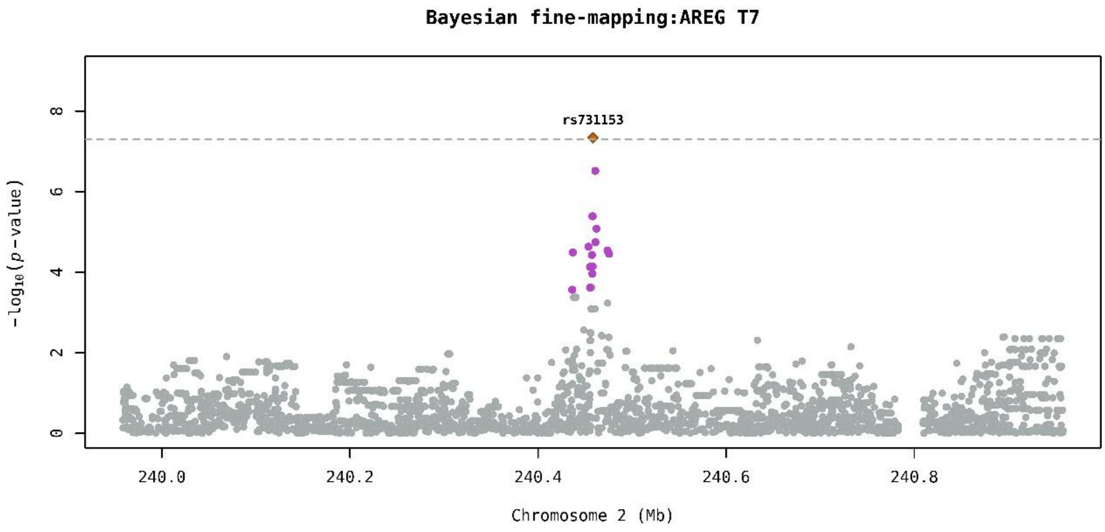
Bayesian fine-mapping:AREG T7
rs731153
$-\log_{10}(p\text{-value})$
0
2
4
6
8
240.0
240.2
240.4
240.6
240.8
Chromosome 2 (Mb)

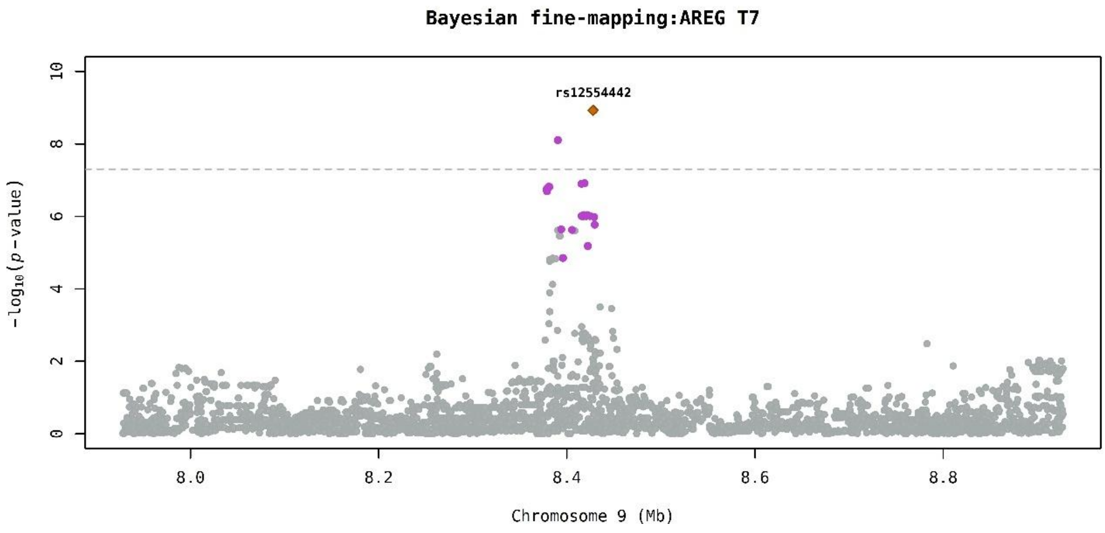
Bayesian fine-mapping:AREG T7
rs12554442
$-\log_{10}(p\text{-value})$
0
2
4
6
8
10
8.0
8.2
8.4
8.6
8.8
Chromosome 9 (Mb)

**Bayesian fine-mapping:AREG T7**

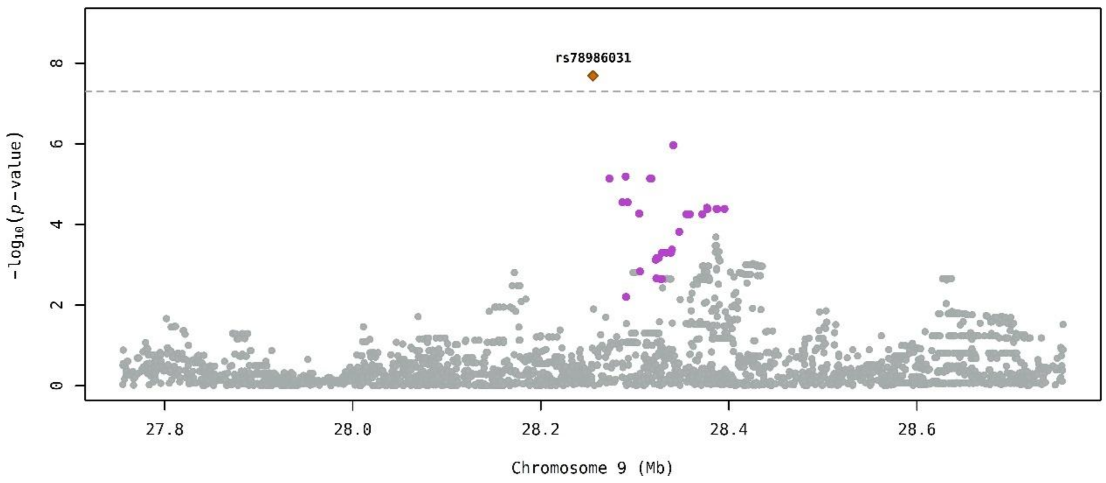

**Bayesian fine-mapping:AREG T7**

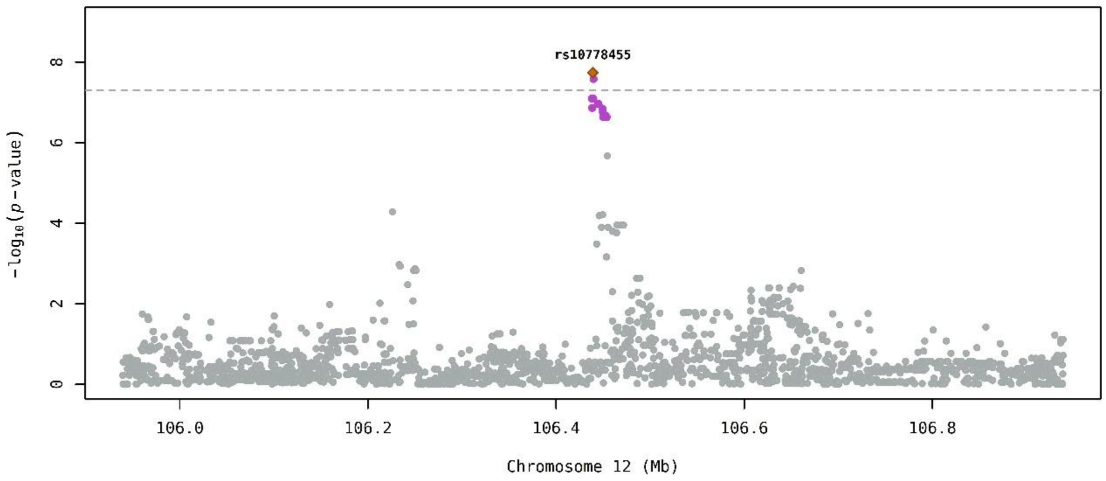

**Bayesian fine-mapping:AREG T7**

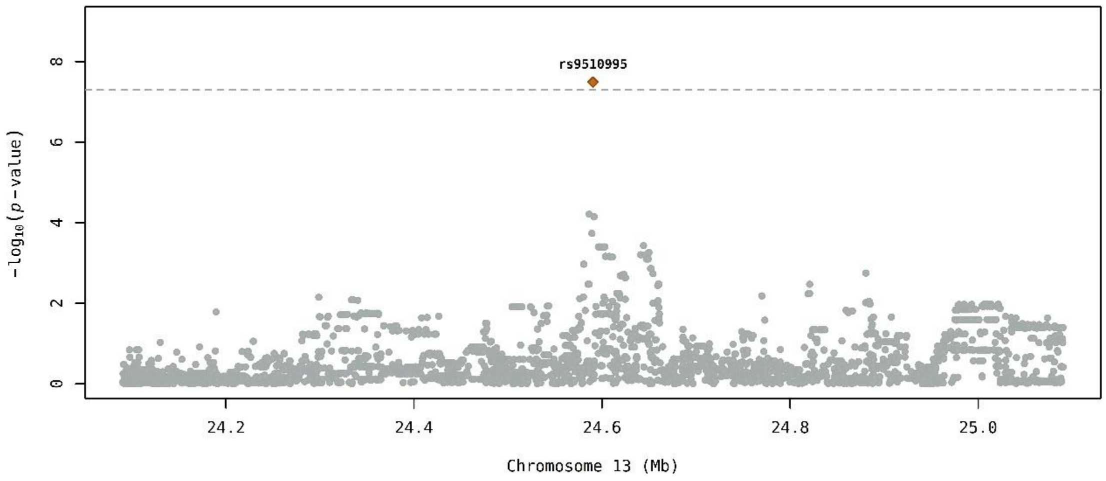

**Bayesian fine-mapping:AREG T7**

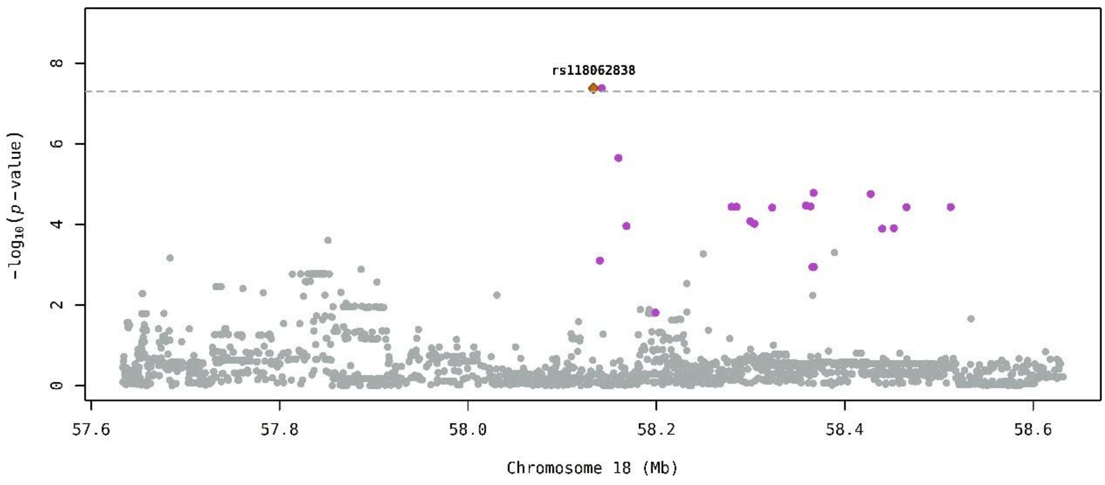

**Bayesian fine-mapping:CXCL16 T1**

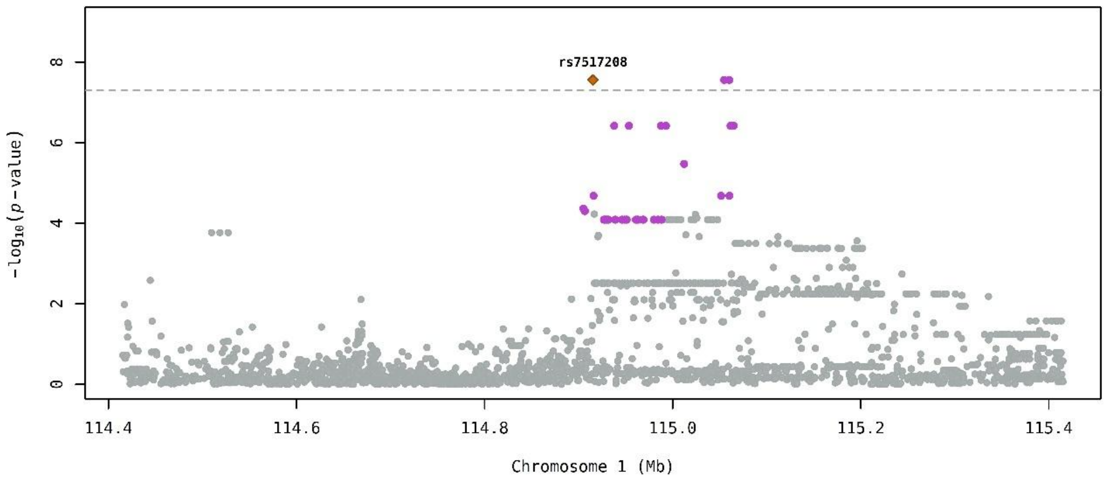

**Bayesian fine-mapping:CXCL16 T1**

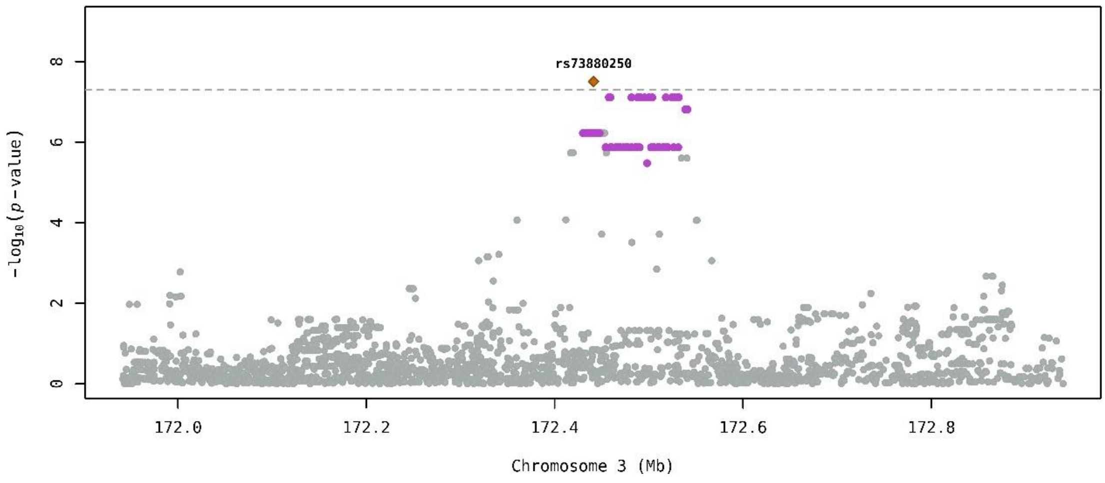

**Bayesian fine-mapping:ICAM1 T2**

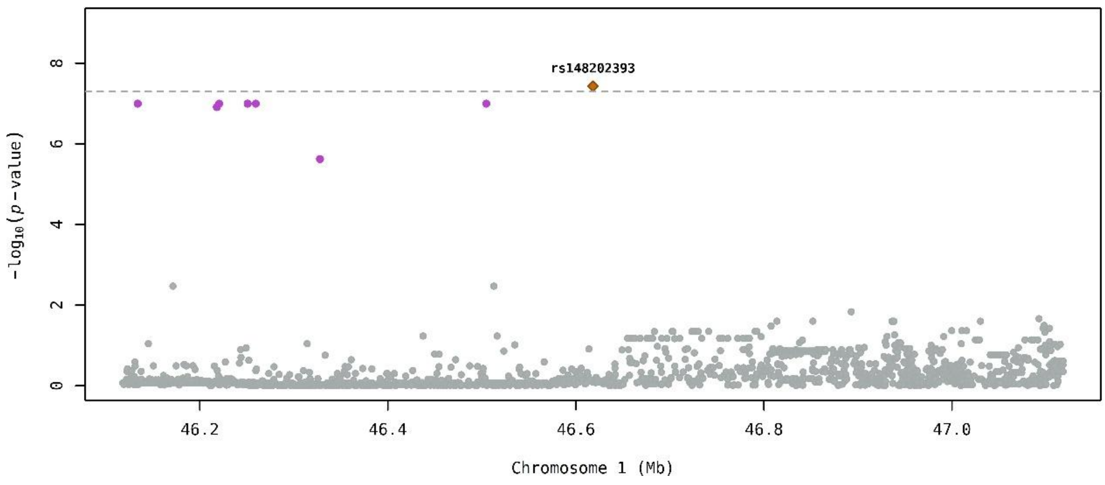

**Bayesian fine-mapping:ICAM1 T2**

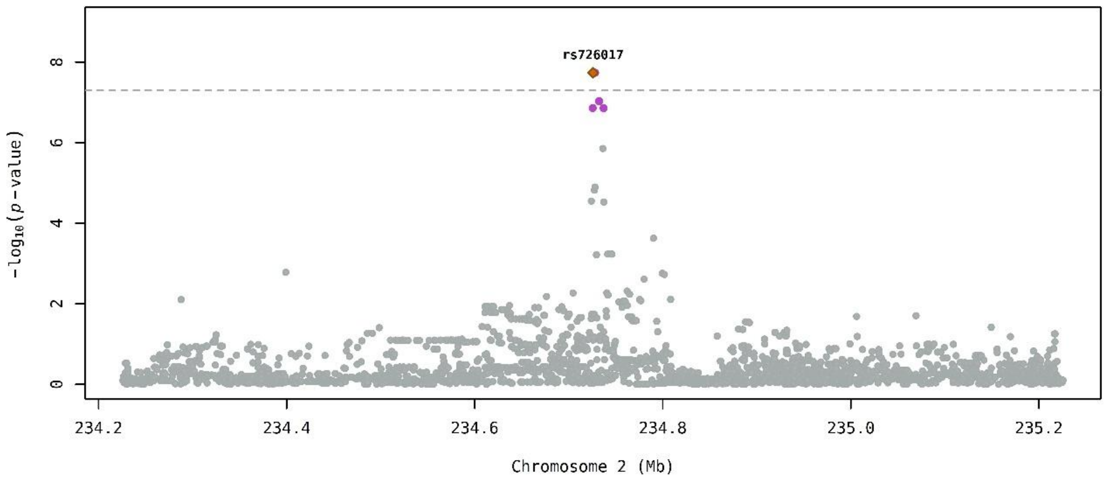

**Bayesian fine-mapping:ICAM1 T2**

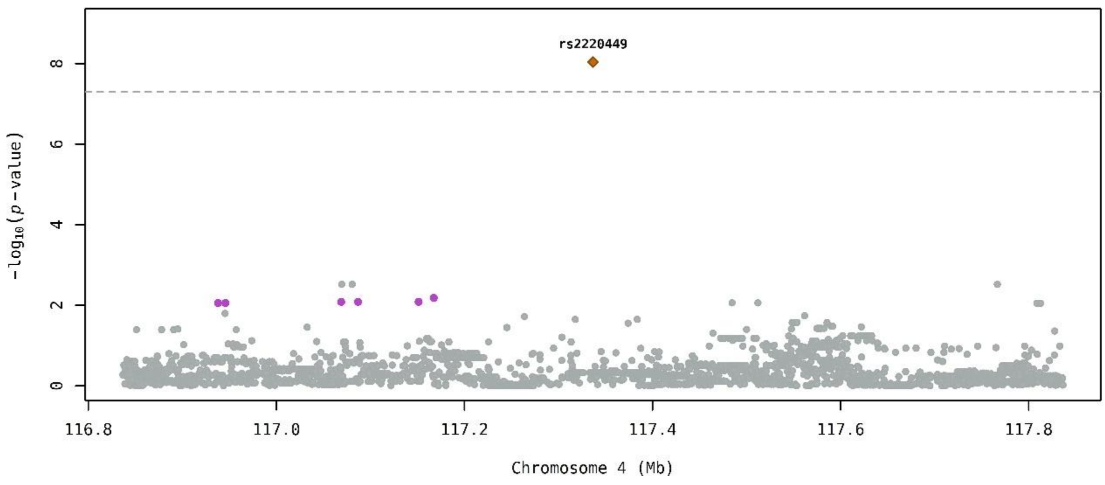

**Bayesian fine-mapping:IL18 T1**

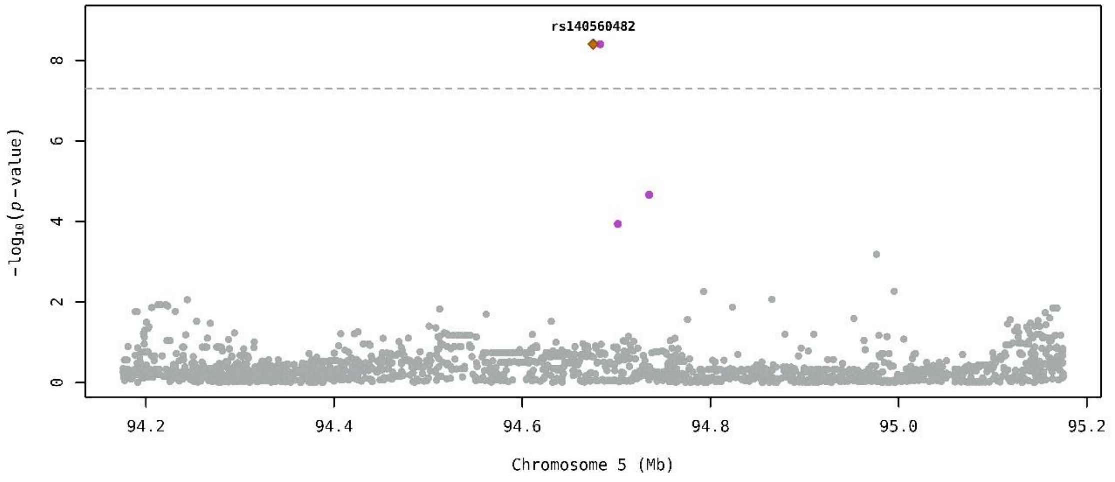

**Bayesian fine-mapping:IL18 T2**

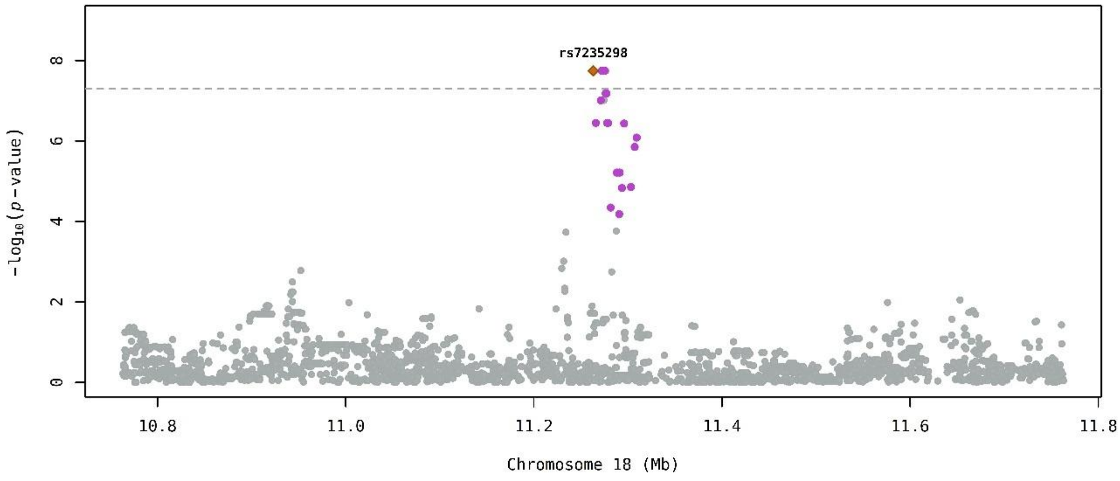

**Bayesian fine-mapping:IL18 T7**

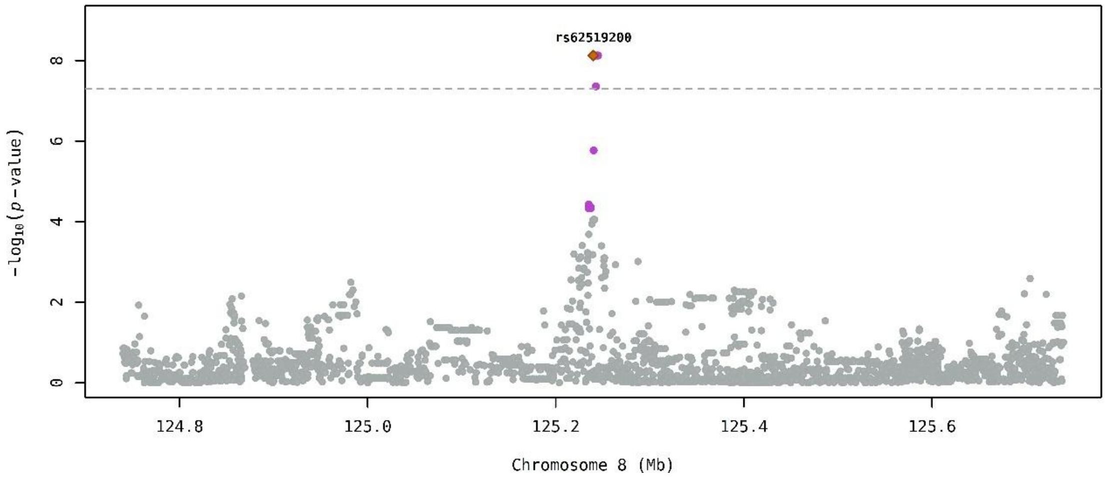

Bayesian fine-mapping:IL1RA T7

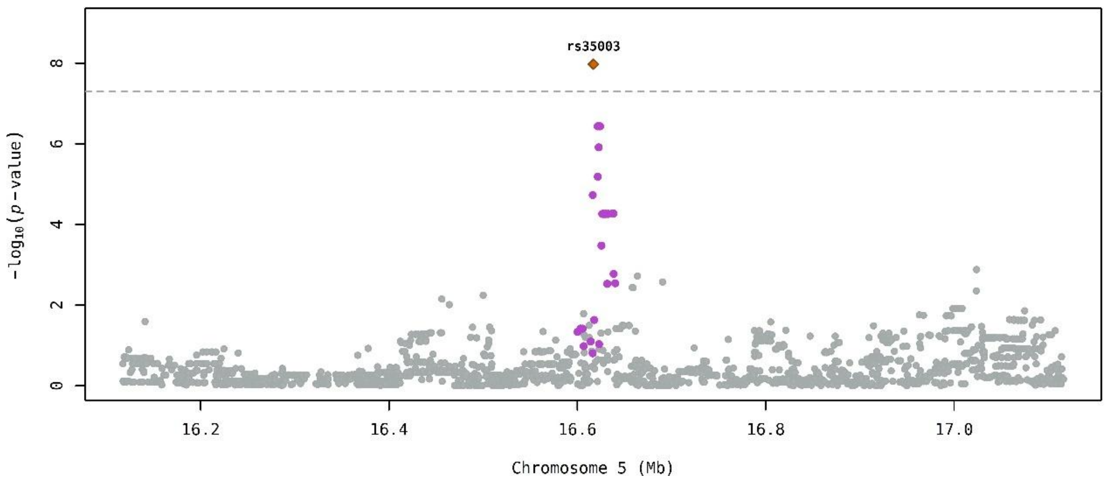

Bayesian fine-mapping:IL1RA T7

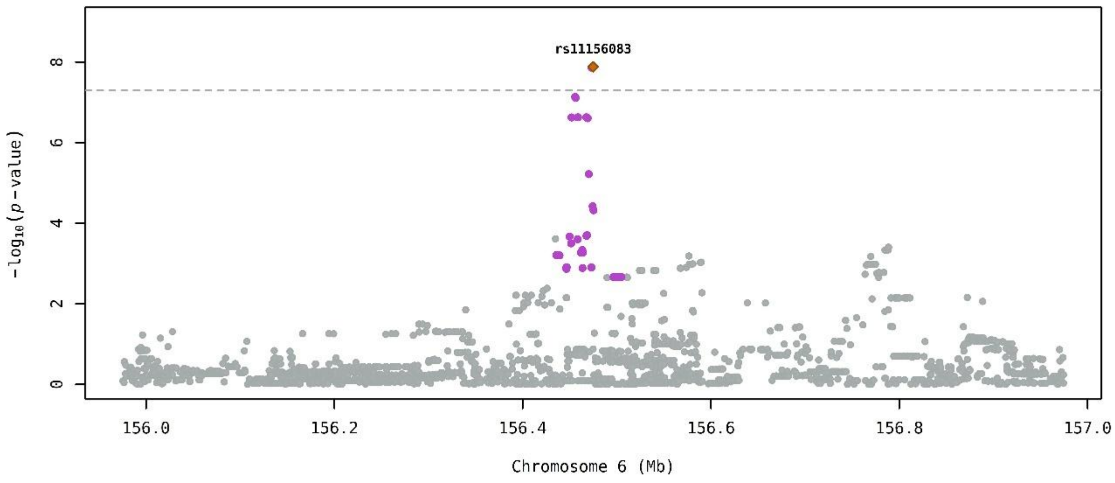

Bayesian fine-mapping:IL1RA T7

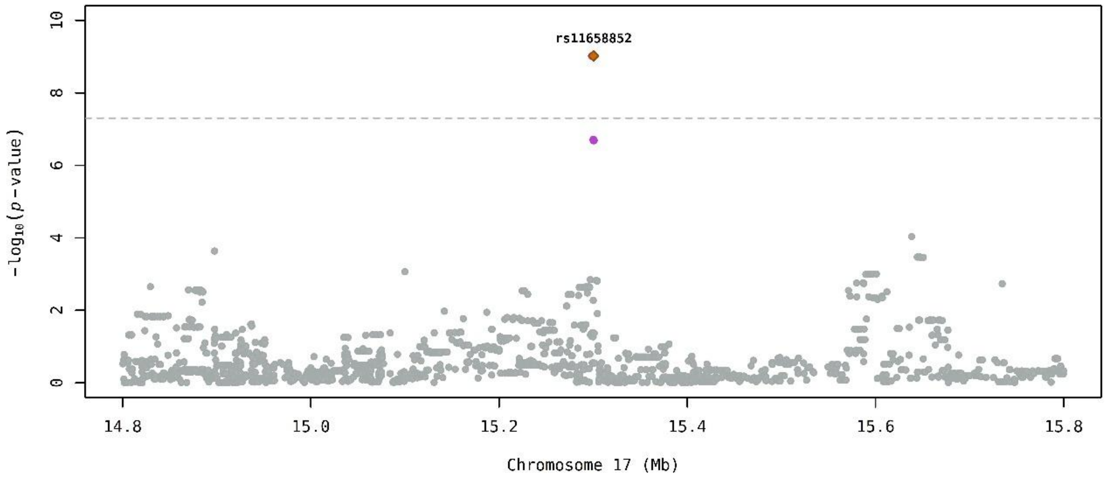

Bayesian fine-mapping:PAI-1 T1

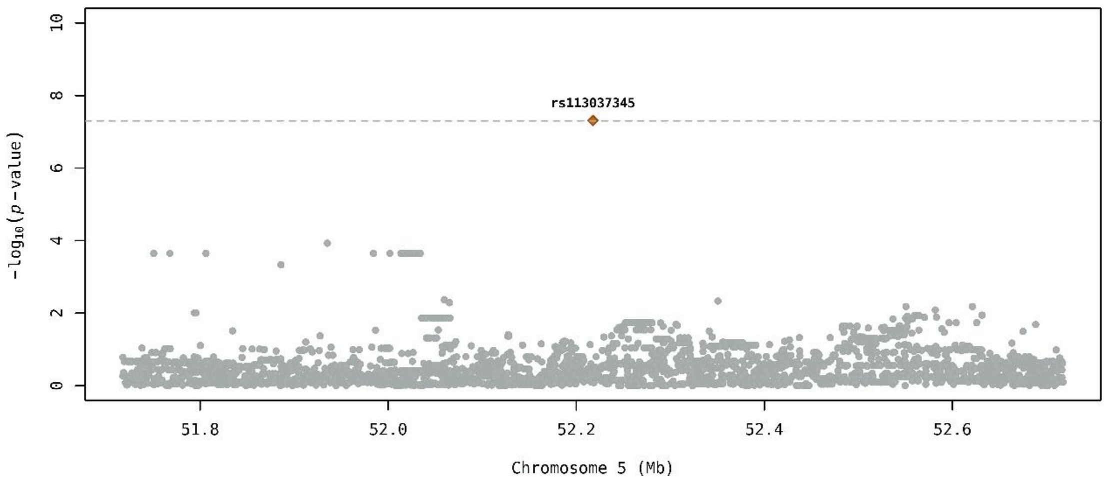

Bayesian fine-mapping:RAGE T2

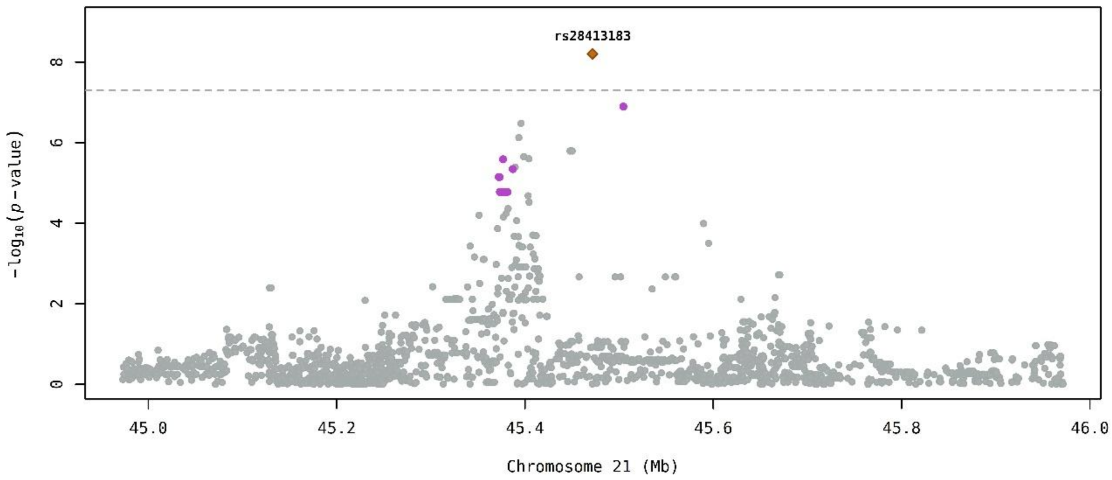

Bayesian fine-mapping:RAGE T7

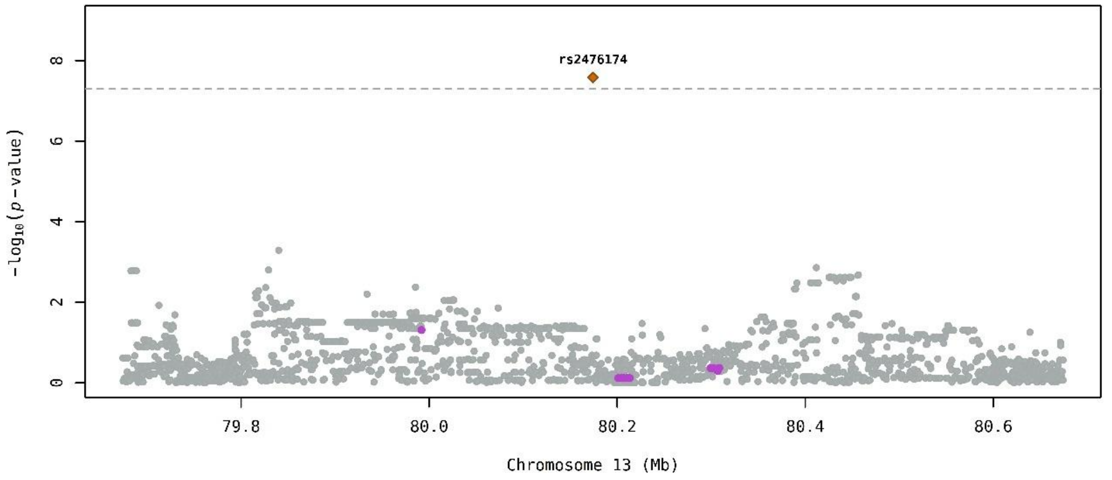

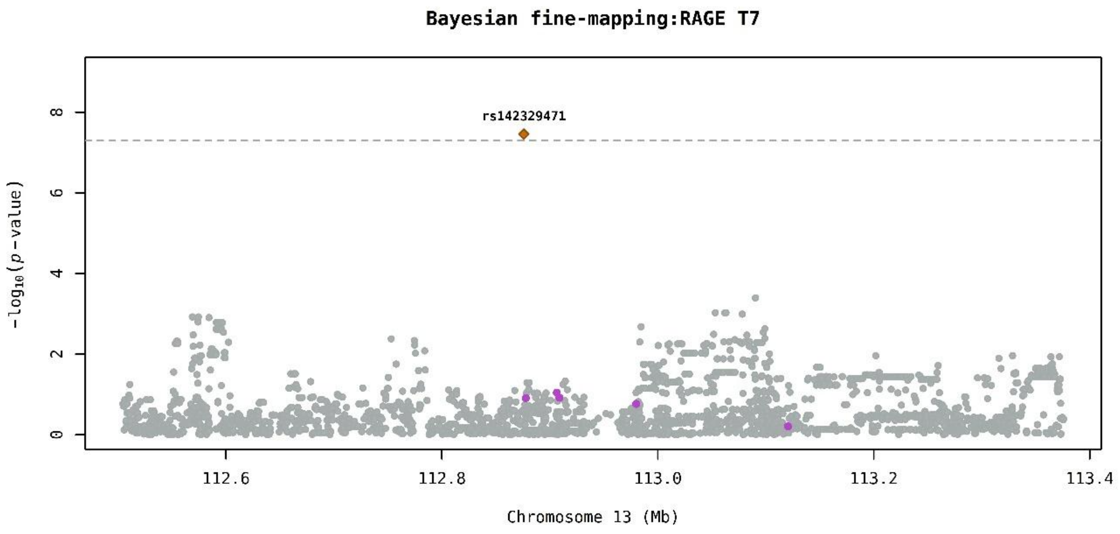
Bayesian fine-mapping:RAGE T7
rs142329471
-log10(p-value)
0
2
4
6
8
112.6
112.8
113.0
113.2
113.4
Chromosome 13 (Mb)

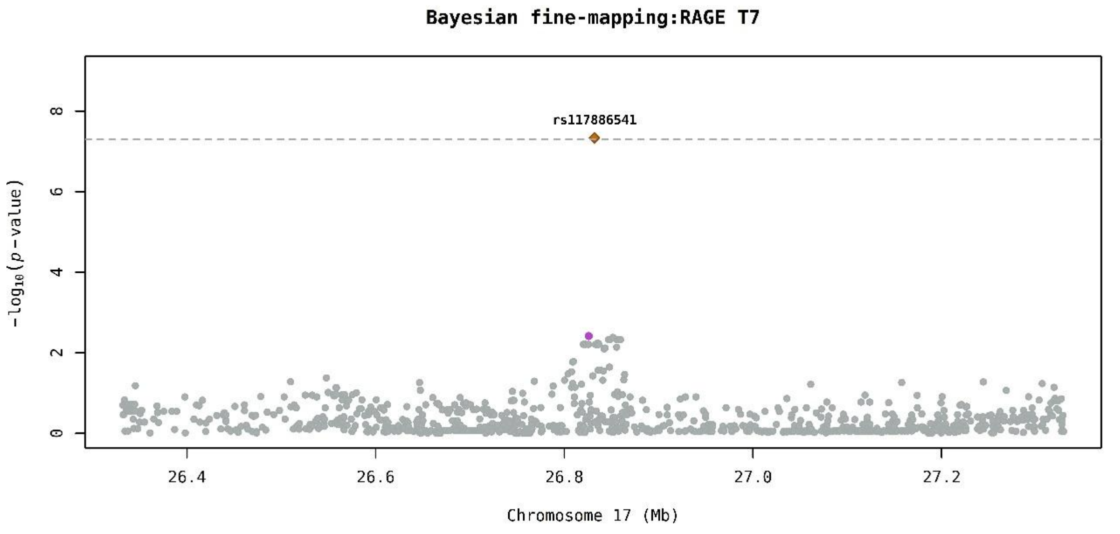
Bayesian fine-mapping:RAGE T7
rs117886541
-log10(p-value)
0
2
4
6
8
26.4
26.6
26.8
27.0
27.2
Chromosome 17 (Mb)

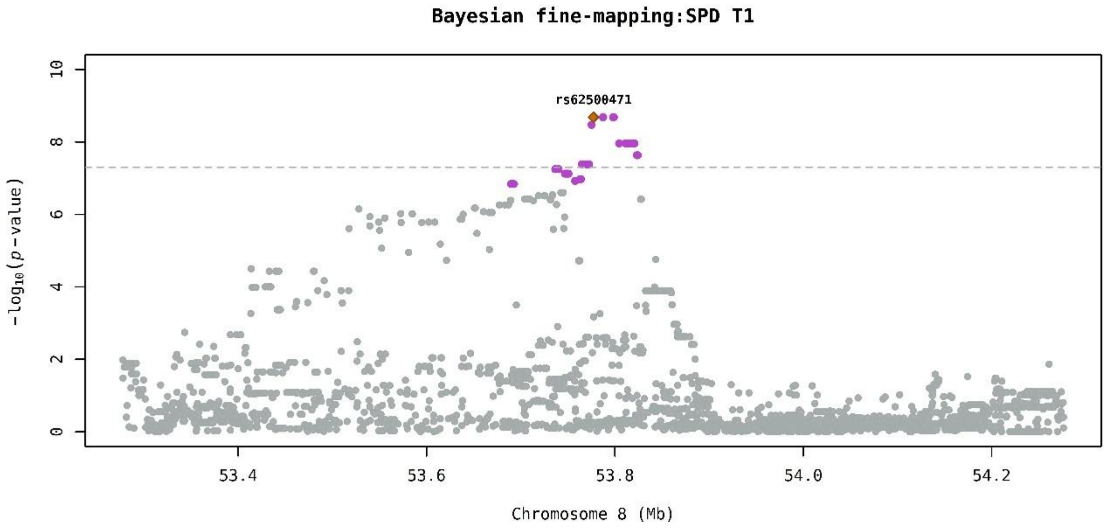
Bayesian fine-mapping:SPD T1
rs62500471
-log10(p-value)
0
2
4
6
8
10
53.4
53.6
53.8
54.0
54.2
Chromosome 8 (Mb)

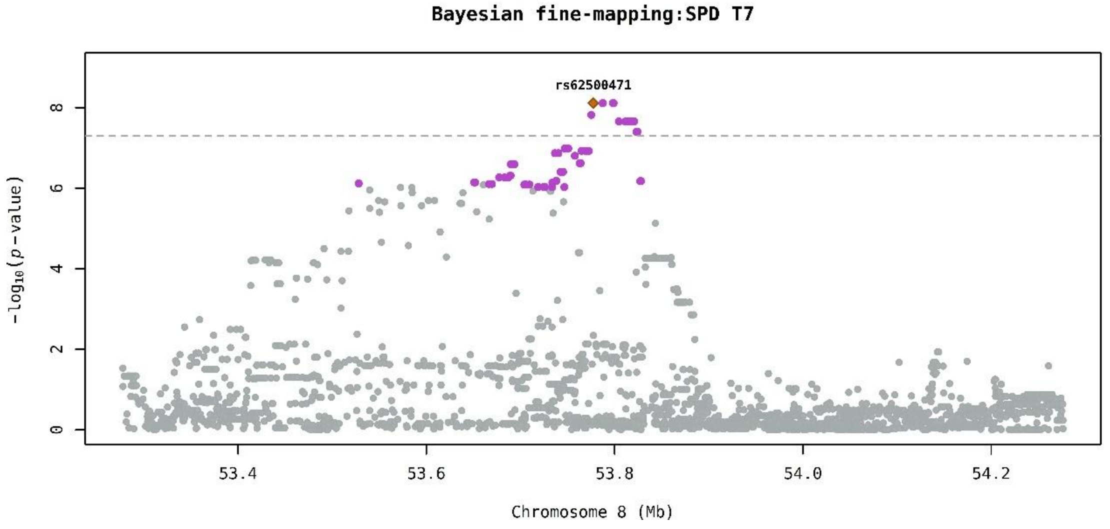

Bayesian fine-mapping:SPD T7
rs62500471
$-\log_{10}(p\text{-value})$
0
2
4
6
8
53.4
53.6
53.8
54.0
54.2
Chromosome 8 (Mb)

**Figure S9. Summary of previous genetic associations with traits (hematopoietic, infectious, respiratory, and Mendelian as provided by OMIM), colocalization, and rare variant analysis (WES) for prioritized genes.**

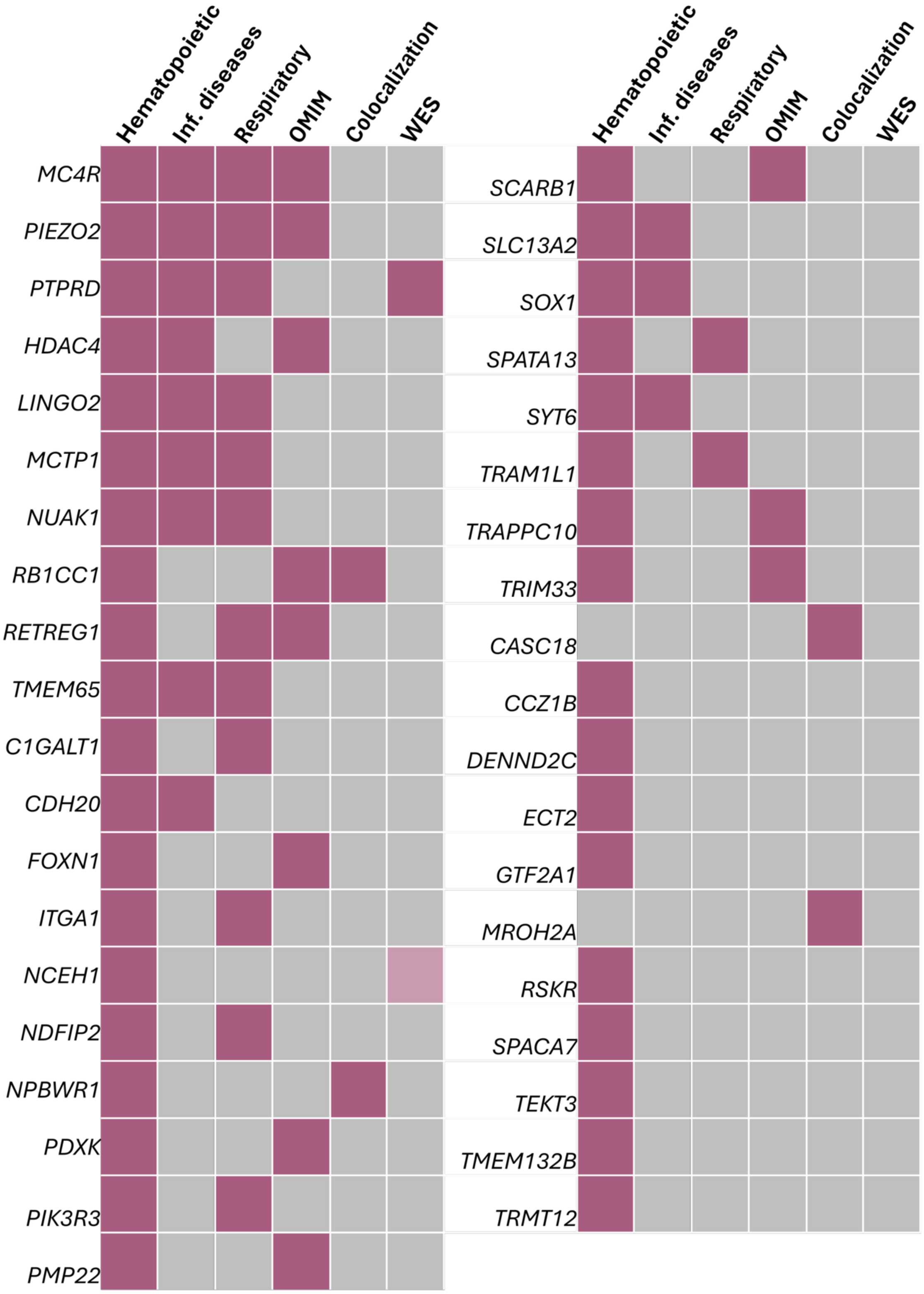
